# Risk and Protective Factors for Self-Harm and Suicide Behaviours among Serving and Ex-Serving Personnel of the UK Armed Forces, Canadian Armed Forces, Australian Defence Force and New Zealand Defence Force: A Systematic Review

**DOI:** 10.1101/2023.09.08.23295121

**Authors:** Charlotte Williamson, Bethany Croak, Amos Simms, Nicola T. Fear, Marie-Louise Sharp, Sharon A.M. Stevelink

## Abstract

**Purpose:** Self-harm and suicide behaviours are a major public health concern. Several factors are associated with these behaviours among military communities. Identifying these factors may have important implications for policy and clinical services. The aim of this review was to identify the risk and protective factors associated with self-harm and suicide behaviours among serving and ex-serving personnel of the United Kingdom Armed Forces, Canadian Armed Forces, Australian Defence Force and New Zealand Defence Force.

**Methods:** A systematic search of seven online databases (PubMed, Web of Science, Embase, Global Health, PsycINFO, PTSDpubs and CINAHL) was conducted alongside cross-referencing, in October 2022. Following an *a priori* PROSPERO approved protocol (CRD42022348867), papers were independently screened and assessed for quality. Data were synthesised using a narrative approach.

**Results:** Overall, 28 papers were included: 13 from Canada, 10 from the United Kingdom, five from Australia and none from New Zealand. Identified risk factors included being single/ex-relationship, early service leavers, shorter length of service, junior ranks, exposure to deployment-related traumatic events, physical and mental health diagnoses, and experience of childhood adversity. Protective included being married/in a relationship, higher educational attainment, employment, senior ranks, and higher levels of perceived social support.

**Conclusion:** Adequate care and support are a necessity for the military community. Prevention and intervention strategies for self-harm and suicide behaviours should be introduced early and should promote social networks as a key source of support. This review found a paucity of peer-reviewed research within some populations. More peer-reviewed research is needed, particularly among these populations where current work is limited, and regarding modifiable risk and protective factors.

## 1. INTRODUCTION

Self-harm and suicide behaviours are a major public health concern, with over 700,000 people dying by suicide and an estimated 14.6 million people engaging in self-harm each year globally [1, 2]. The aetiology and onset of these behaviours is complex and multifaceted; prevalence and risk are influenced by several factors including age, sex, ethnicity, geographical region and occupation [3–5]. Military populations are a potentially at-risk group as military service impacts on the health and wellbeing of personnel during and after service [6]. Self-harm and death by suicide appear to be relatively rare among the military community, although rates of both behaviours have increased in recent years but remain either lower or comparable to the general population [7–12]. Suicide behaviours can present as ideation (i.e. thoughts) and attempts which are also prevalent among serving and ex-serving personnel, with a reported 11% global prevalence of these behaviours in the entire military [13].

Previous research has reported that serving and ex-serving personnel with mental health diagnoses are at risk of self-harm and suicide behaviours, for example post-traumatic stress disorder (PTSD) [14–18]. Another specific at-risk group includes those separated from the military [19–21], especially if separation occurred involuntarily (e.g. medical discharge) [22]. Additionally, younger age is a risk factor [22–24]; a recent UK study reported suicide risk was two-to-three times higher in men and women under 25 years old who had left service (vs the general population) [24]. Ex-serving personnel with a shorter length of service (<10 years of service) were also at increased risk [24]. Protective factors have been less frequently explored, but include social support [25, 26], higher educational attainment [27], employment [28] and holding a more senior rank (e.g. officer) [15, 18].

There remains a paucity of research exploring the risk and protective factors for self-harm and suicide behaviours among several military populations, with available literature focussing on a limited number of countries. This review focused on military populations from four nations from the Five Eyes Alliance: namely the UK, Canada, Australia and New Zealand. The Five Eyes nations (which also includes the United States (US)) have a combined military population estimated at 2.6 million, are all developed countries which share a common language, have similarities in society and culture, broadly similar military involvement, and well-resourced military healthcare systems [29]. Of the limited systematic reviews in the field, studies on US military populations make up all or the majority of included papers [26, 30, 31]. To address this gap in the literature, the current review focuses on the remaining four Five Eyes nations where attention has been limited previously. Additionally, the US was excluded due to differences in firearms culture, including access, licensing and laws which affects access to means of suicide; a recent systematic review highlighted that US veterans are at substantially increased risk of firearm suicide and have higher rates of firearm ownership than the US general population [32].

The current systematic review aimed to identify risk and protective factors associated with self-harm and suicide behaviours among serving and ex-serving personnel of the UK Armed Forces, Canadian Armed Forces, Australian Defence Force and New Zealand Defence Force.

## 2. METHOD

### 2.1. Design

This systematic review was conducted following Cochrane methodology and PRISMA guidelines **(Supplementary 1)**. Prior to commencing the review, the protocol was registered with PROSPERO (CRD42022348867).

### 2.2. Search Strategy

Seven electronic databases were searched in October 2022: PubMed (including MEDLINE and PubMed Central), Web of Science, Embase, Global Health, PsycINFO, PTSDpubs and CINAHL. All databases were searched using pre-defined terms related to: (1) self-harm and suicide behaviours, (2) the military, and (3) geographical locations. See **Supplementary 2** for full search strategy.

The search included all original, peer-reviewed work that identified risk and/or protective factors associated with self-harm and/or suicide behaviours both during (serving personnel) and after (ex-serving personnel/veterans) military service. Restrictions were placed on publication dates from 1^st^ January 2001 to 30^th^ September 2022 to allow for the coverage of the start of the Iraq and Afghanistan conflicts. Additionally, these limits were chosen due to better availability and quality of literature in the field of military medicine since 2001 [33].

PROSPERO was searched to identify systematic reviews and meta-analyses of relevance. The reference lists of included studies and other relevant studies were searched. At least one expert from each nation was contacted to ensure key papers had been identified. If the full-text of a paper was not available online, authors were contacted for access.

### 2.3. Study Selection Criteria

Eligibility was determined using the following criteria:

- Published in English
- Original, peer-reviewed work
- Published 1^st^ January 2001 to 30^th^ September 2022
- Studies that reported risk and/or protective factors for outcomes of self-harm, suicidal ideation, suicide attempts and/or completed suicide
- Population comprising of serving and/or ex-serving personnel (Regulars/Reservists; Navy/Army/Air Force/Marines) from the UK Armed Forces, Canadian Armed Forces, Australian Defence Force or New Zealand Defence Force
- If military only make up part of the reported sample, only papers that reported on outcomes for serving/ex-serving personnel separate from the other population (e.g., general population) were included

Exclusion criteria included:

- Study design: qualitative, systematic reviews/meta-analyses, pilot studies, case-control studies, case reports/series, study protocols, grey literature, conference abstracts/papers, dissertations/theses, and editorials
- Outcomes of assisted suicide (i.e. the act of deliberately assisting another person to kill themselves [34]) and suicide bombing (i.e. a terrorist bomb attack in which the perpetrator expects to die whilst killing a number of other people)
- Studies that only included a population sample of conscripts (i.e., people compulsorily enlisted into the military), cadets or officer students

### 2.4. Screening and Data Extraction

Following the initial search, all identified studies were imported into Endnote 20 and duplicates were removed. CW independently reviewed the titles/abstracts of all papers. Subsequently, the full papers for the remaining studies identified as relevant were then reviewed. The reference lists of all included papers were manually searched for any additional papers of relevance (i.e., cross-referencing). BC independently performed second reviewer screening on a sample of studies (10% at title/abstract screening stage and 20% at full text screening stage). The reviewers (CW and BC) independently decided which studies met the eligibility criteria to be included in the review and, at full text screening stage, noted any reasons for exclusion. Any discrepancies were resolved through discussion. Interrater reliability was calculated at each screening stage; agreement was 99% at title/abstract and 100% at full-text stage.

The following data were extracted independently by CW for all included papers:

- **General information:** title, lead author, publication date, journal title, location/country of study
- **Study characteristics:** study aim, study design and methods, response rate (where relevant), sample size, data collection date
- **Sample characteristics**: age (in years), sex distribution, ethnicity, population type (e.g., clinical/non-clinical)
- **Military characteristics:** serving status (serving/ex-serving), engagement type (regular/reserve), service branch, rank, deployment, era of service
- **Outcomes:** type (self-harm, suicidal ideation, suicide attempts, completed suicide), definition and measures
- **Associated factors**
- **Study findings:** conclusions, limitations, future research

Statistical findings are reported in **Supplementary 3**. Where available, adjusted odds ratios or effect estimates have been presented. Otherwise unadjusted results have been reported and clearly identified.

### 2.5. Quality Assessment

The quality of each included paper was assessed independently by CW using the National Heart, Blood and Lung Institute (NHBLI) tailored quality assessment tools [35]. BC independently performed second reviewer quality assessment on a sample of papers (20%). Any discrepancies were resolved through discussion. Papers were not excluded based on their quality, instead, results from the assessment provided additional insight into the quality of research in the field.

## 3. RESULTS

Overall, 4,576 papers were identified, of which 497 duplicates were removed **(Supplementary 4)**. The title/abstracts of 4,079 papers were screened, leaving 94 papers at full-text stage. Experts in the field identified no additional peer-reviewed papers and cross-referencing identified one additional paper of relevance. Overall, 28 papers met the inclusion criteria **(Supplementary 5)**.

### 3.1. Study Characteristics

Included studies used a range of study designs; 23 cohort studies [15, 18, 27, 28, 36–54] and five retrospective cohort studies [55–59] **(Table 1)**. The majority of papers were from Canada (n=13) [36, 38–40, 43–45, 49–51, 58, 59] and the UK (n=10) [15, 18, 46, 47, 52–57]. The remaining papers were from Australia (n=5) [27, 28, 41, 42, 48], but none were from New Zealand. Papers explored military samples of ex-serving personnel only (n=12) [38–41, 43, 49–51], serving personnel only (n=8) [27, 28, 42, 44–47, 52, 55–57, 59] and mixed samples of serving and ex-serving personnel (n=8) [15, 18, 36, 37, 48, 53, 58, 60]. Of the 28 included papers, 24 included mixed male and female samples, however the percentage of male participants typically ranged from around 85% to 99% [15, 18, 28, 36, 38–40, 43–59]. Of the remaining four papers, three used all male samples [27, 41, 42] and one did not explicitly report on the sex of the sample directly within the paper [37].

**Table 1.**
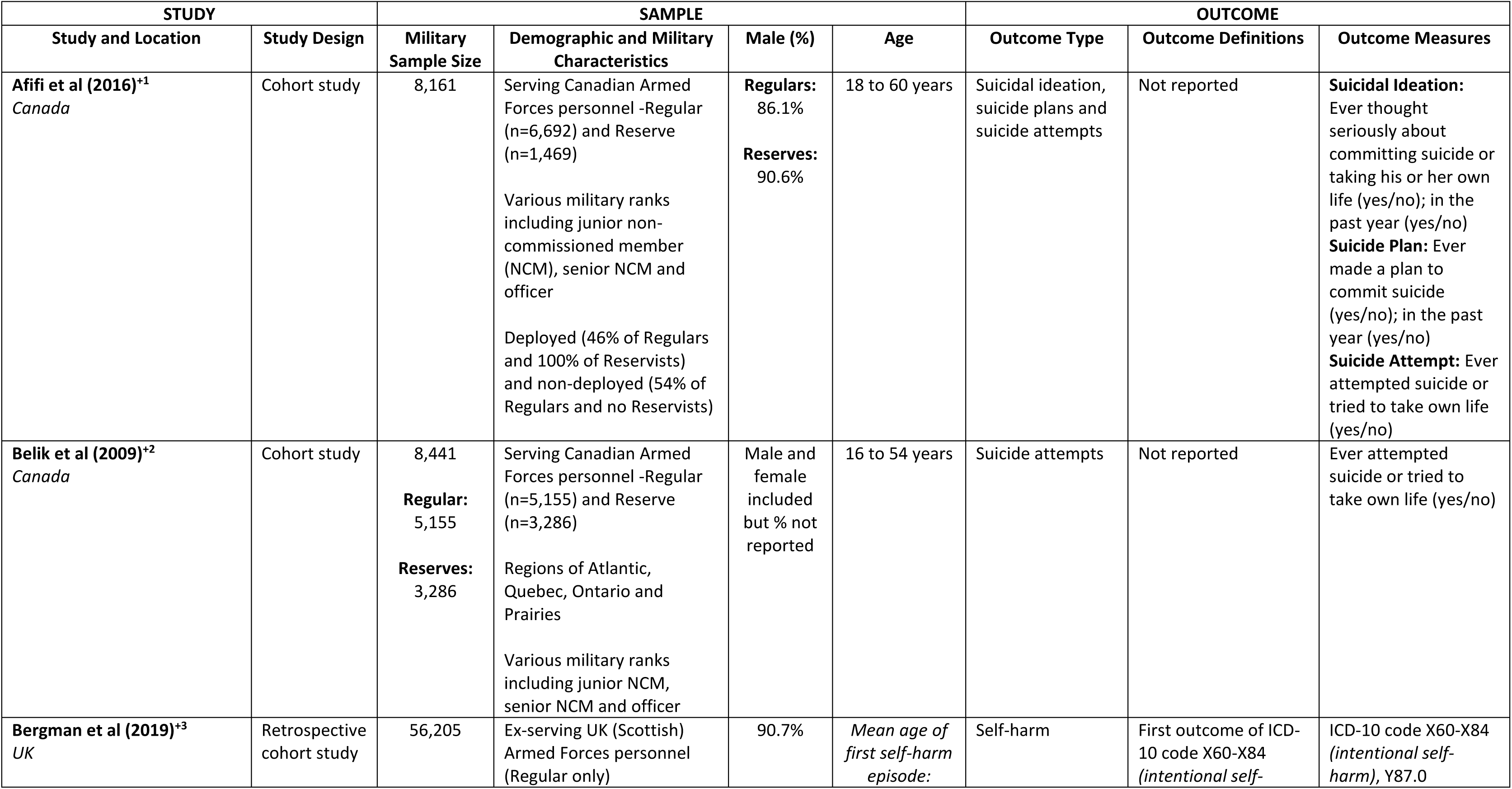
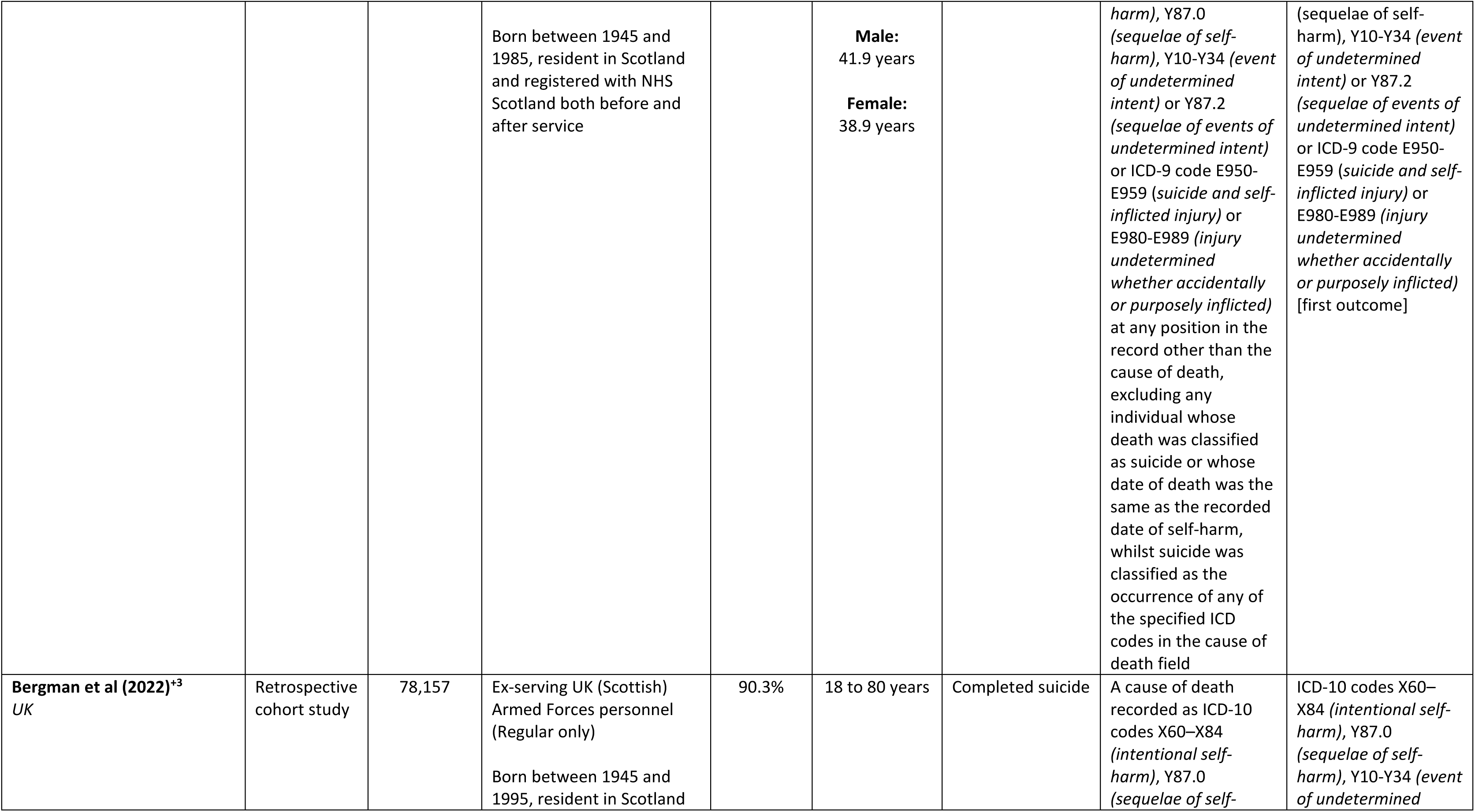
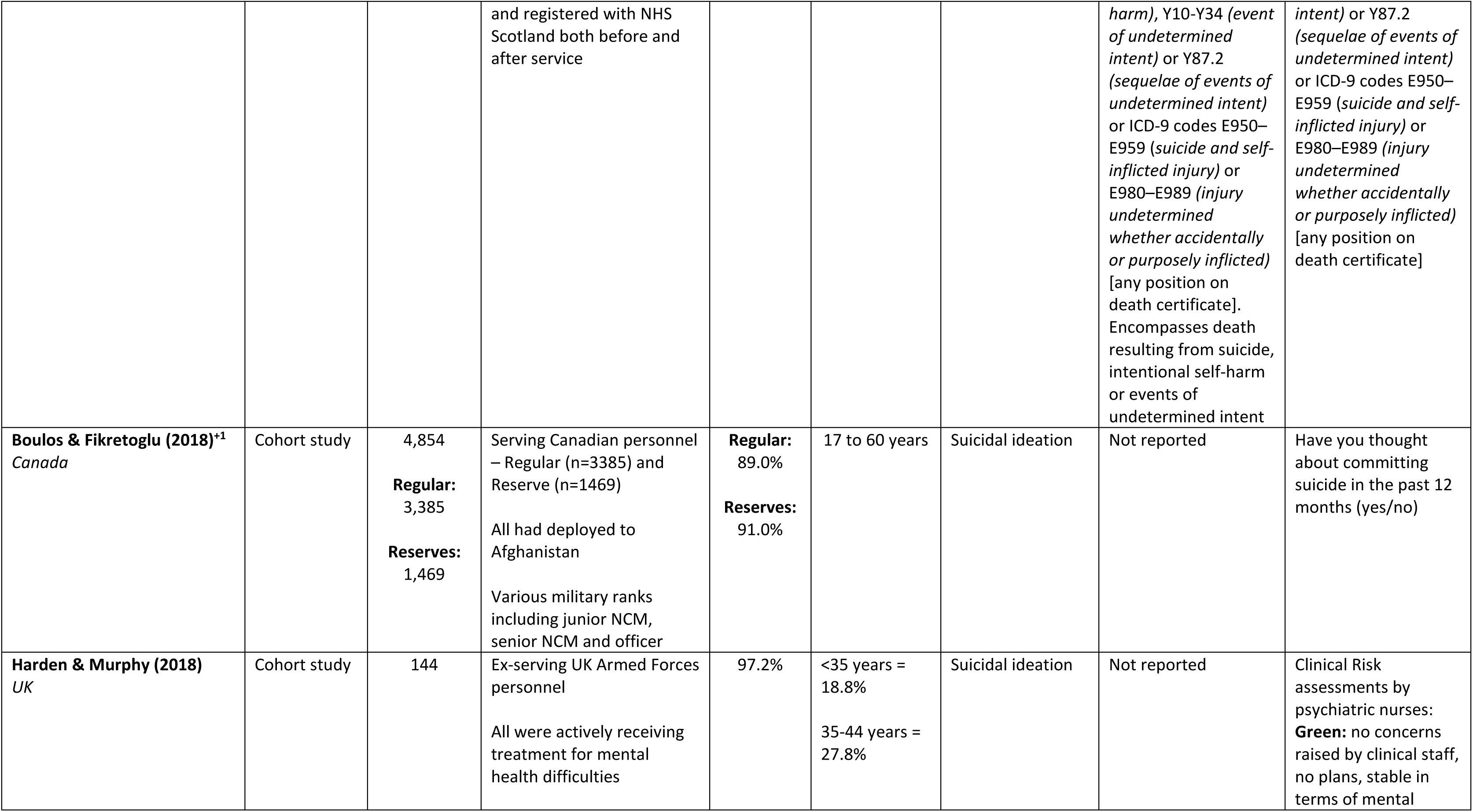
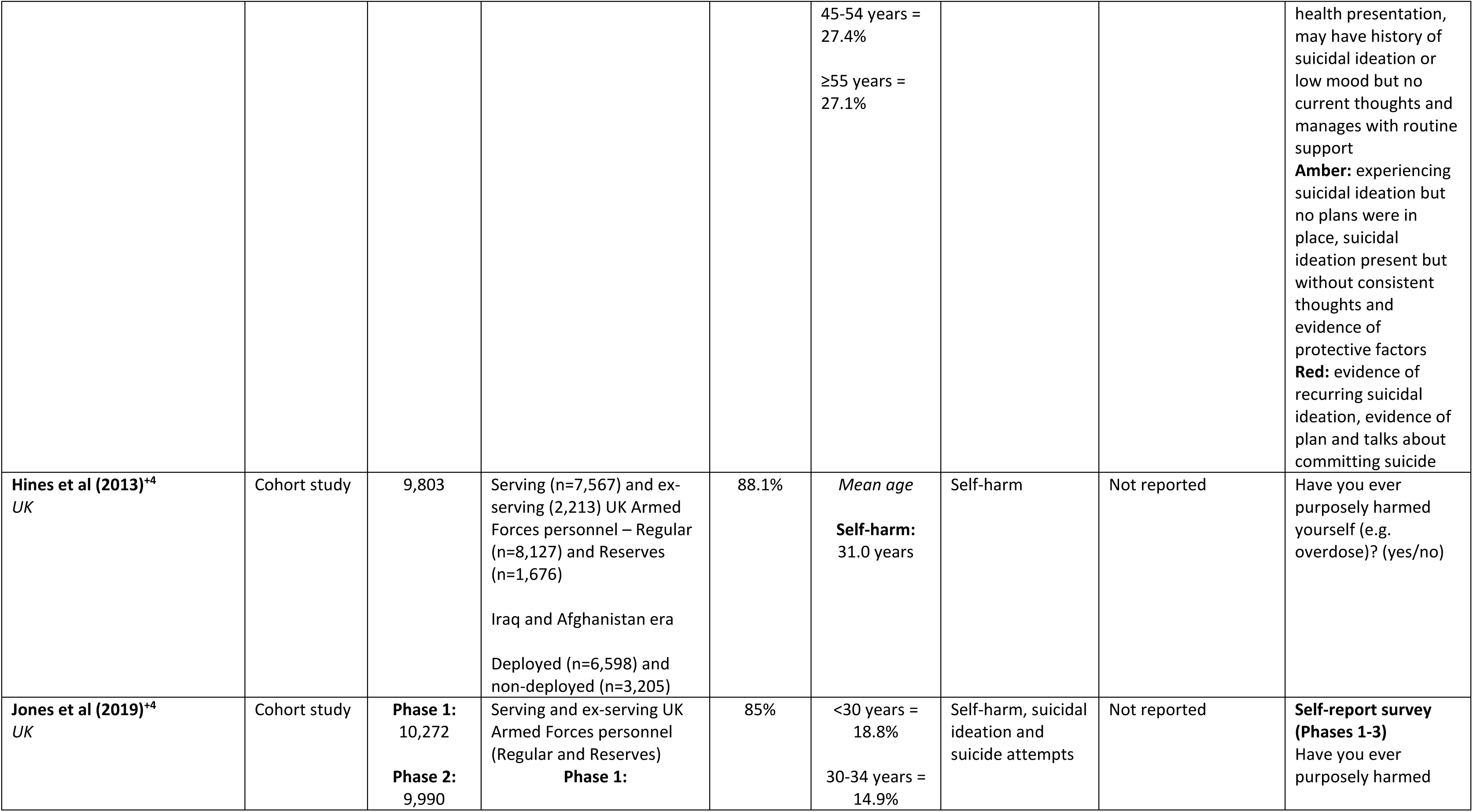
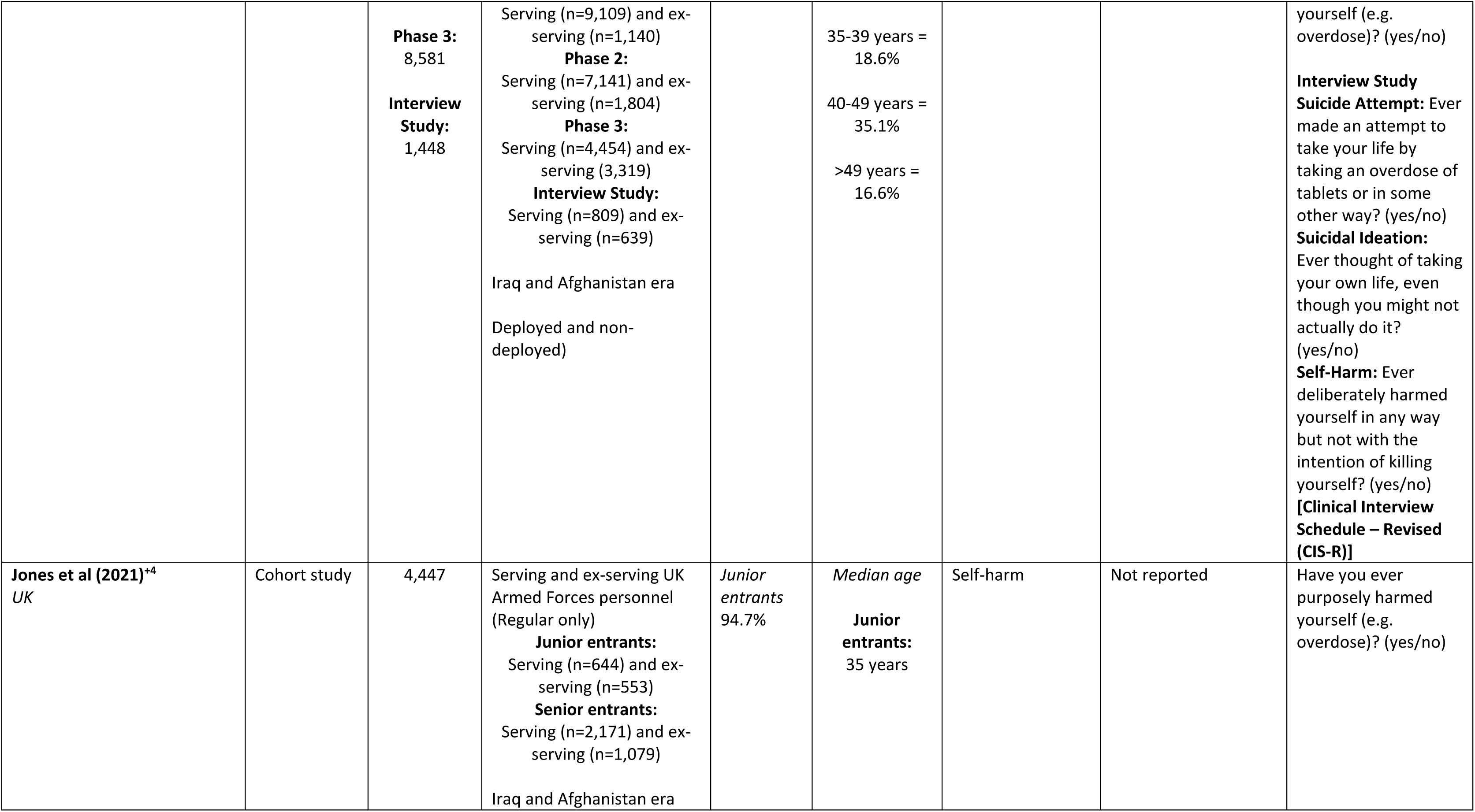
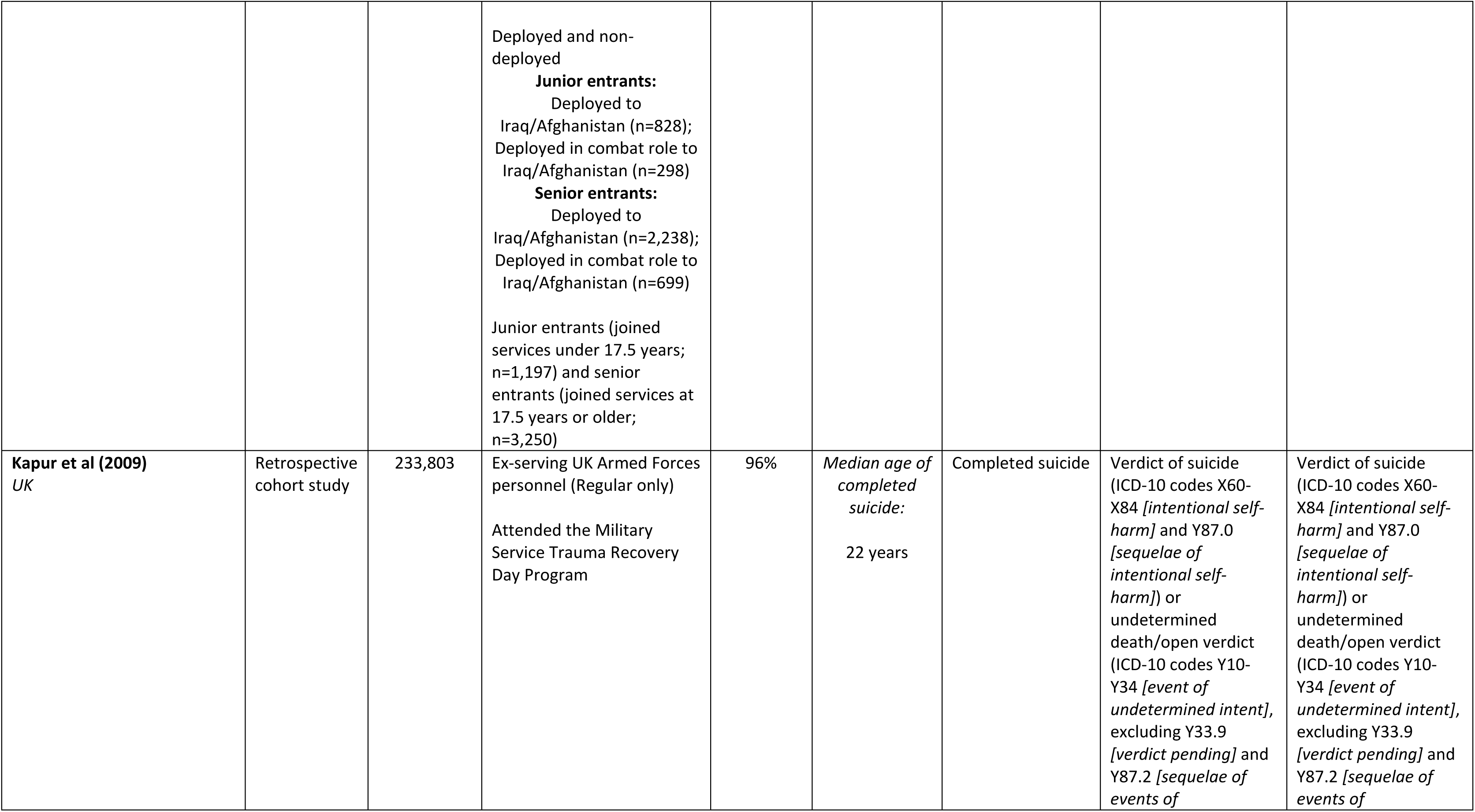
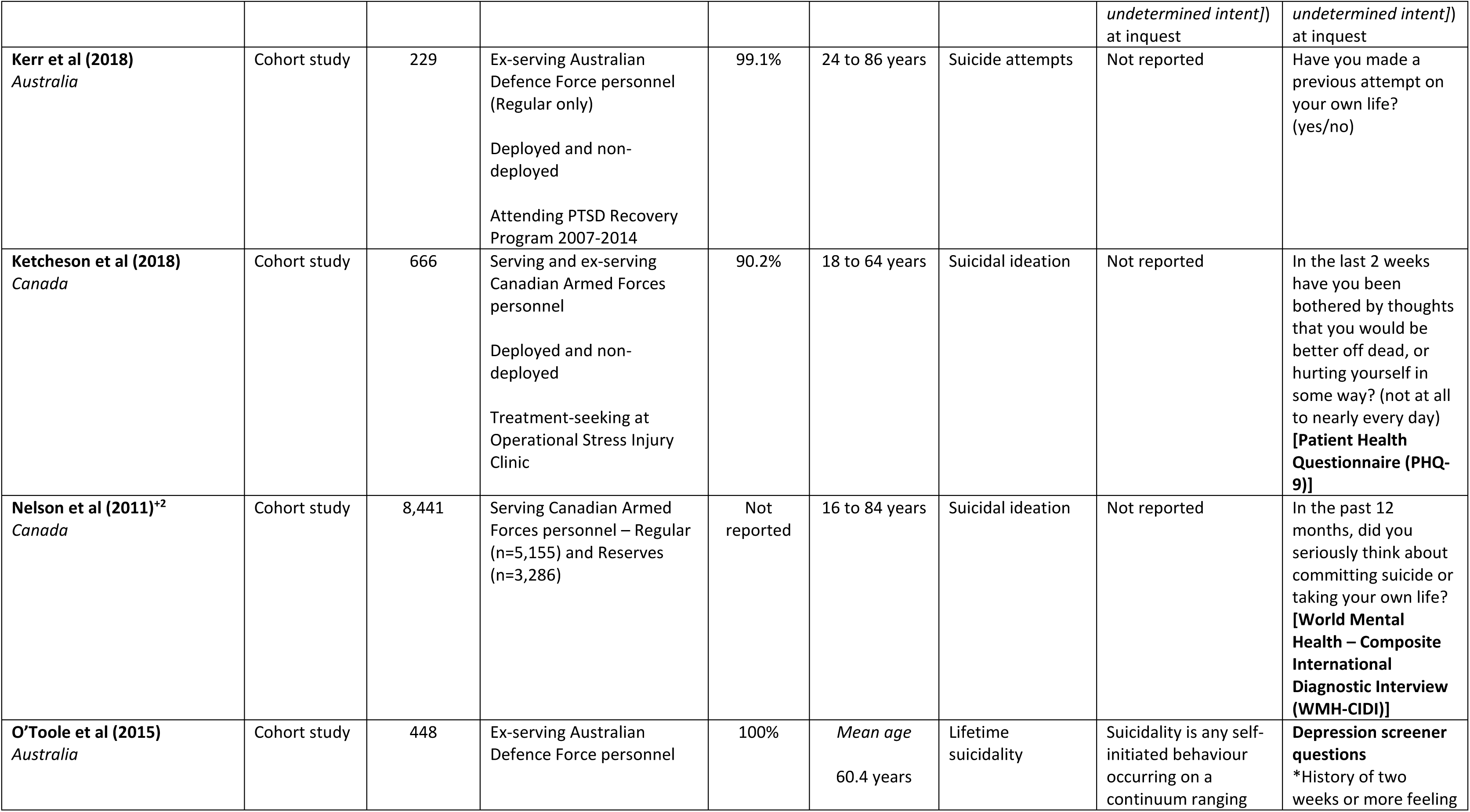
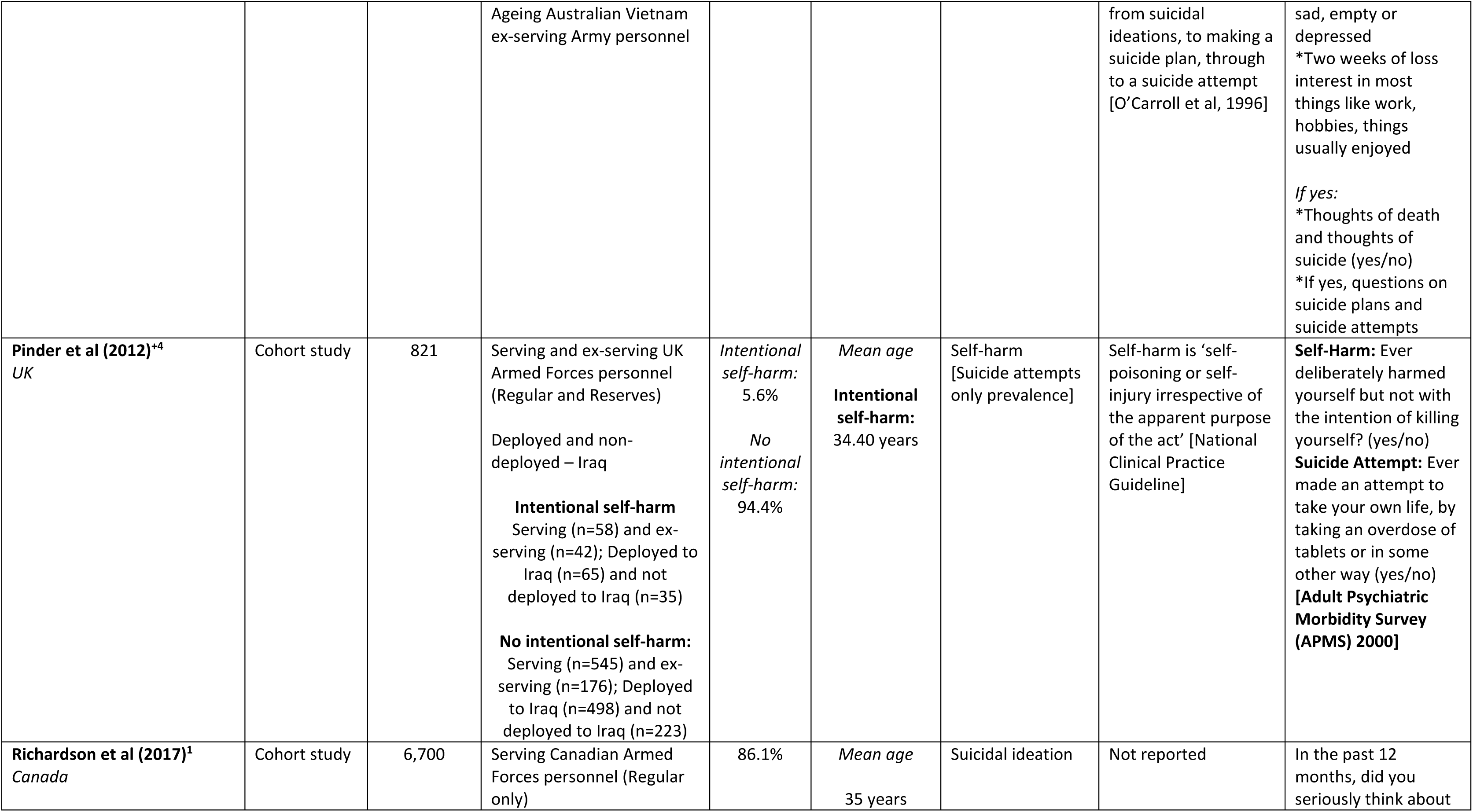
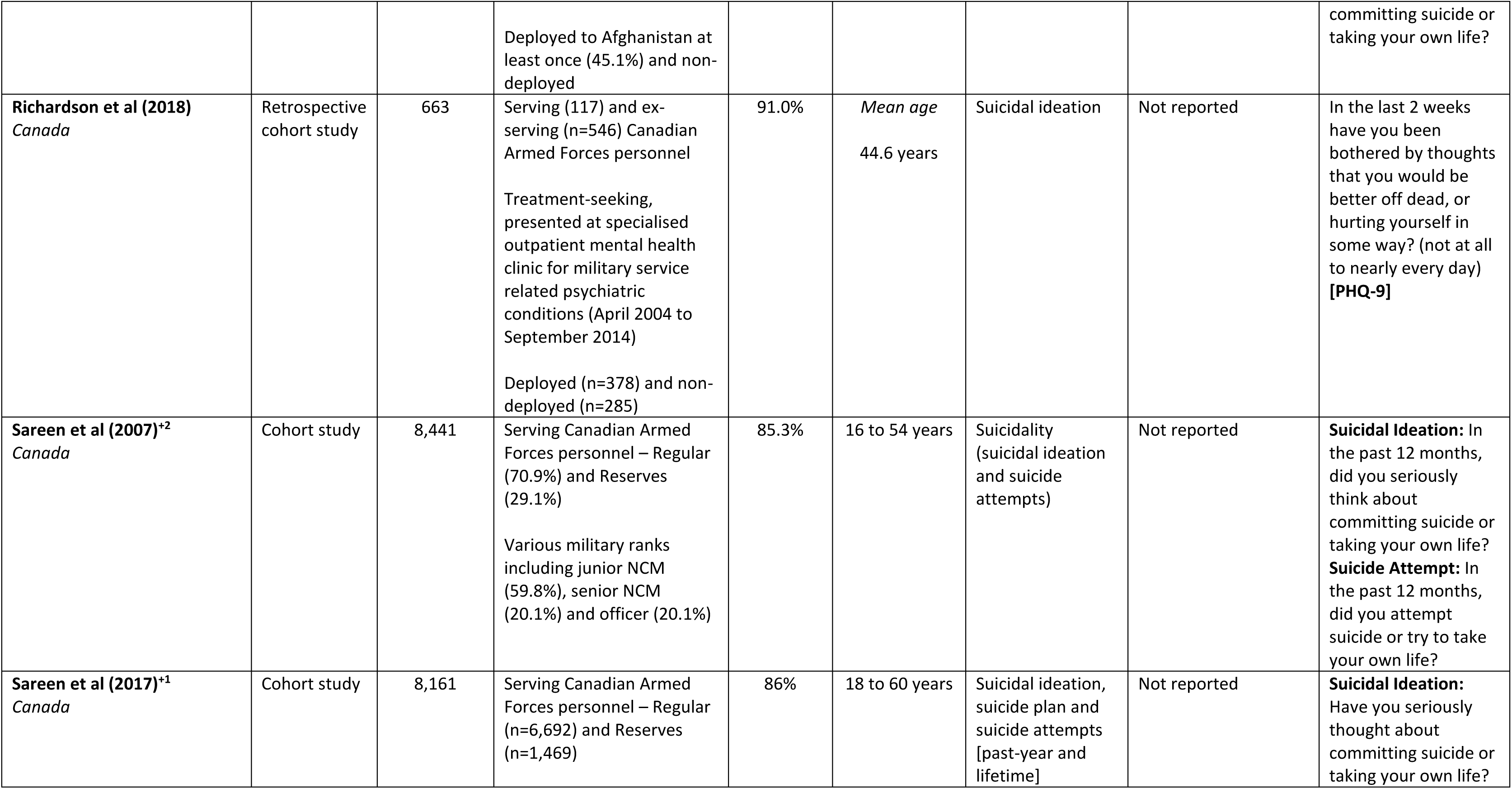
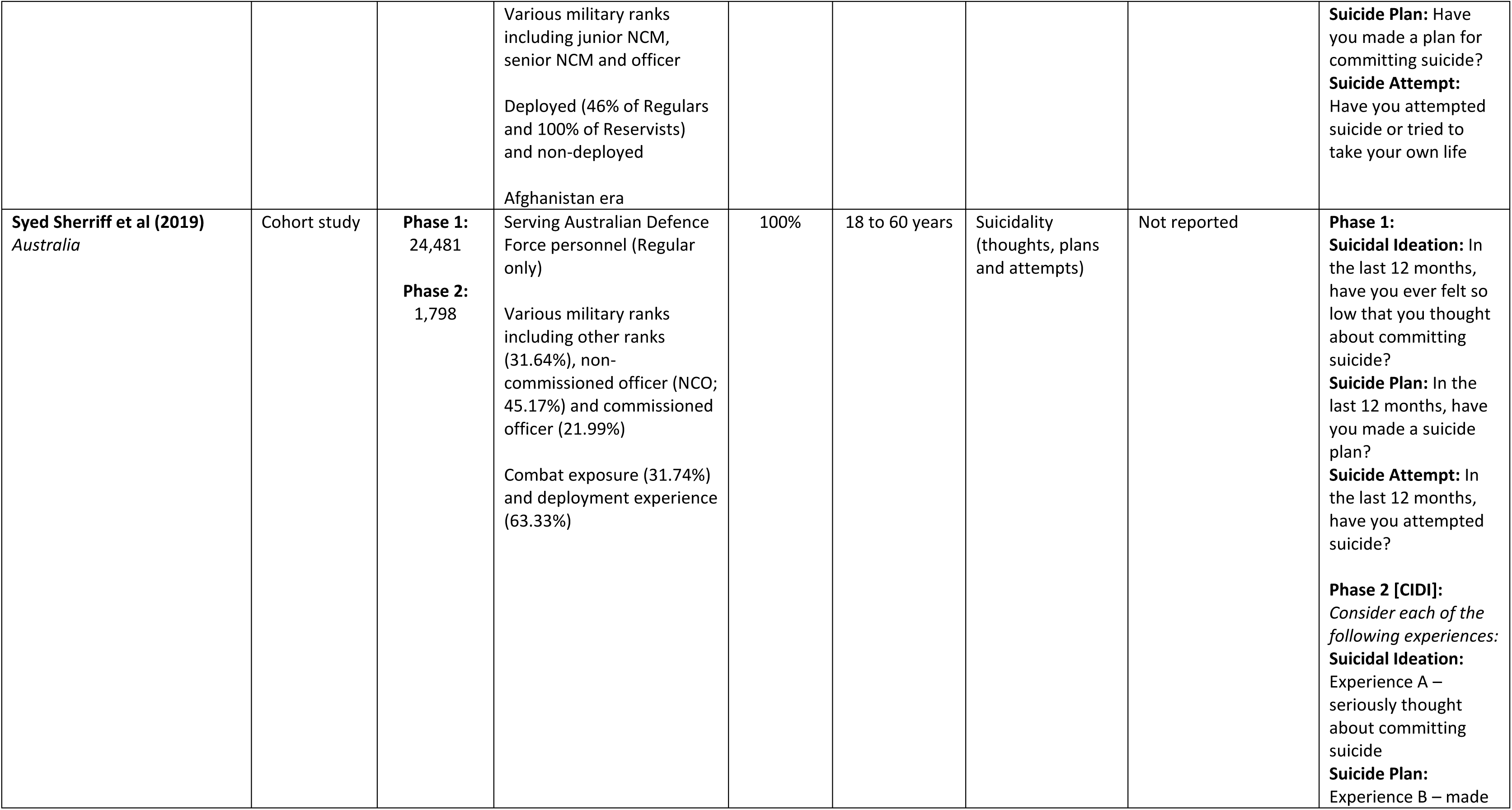
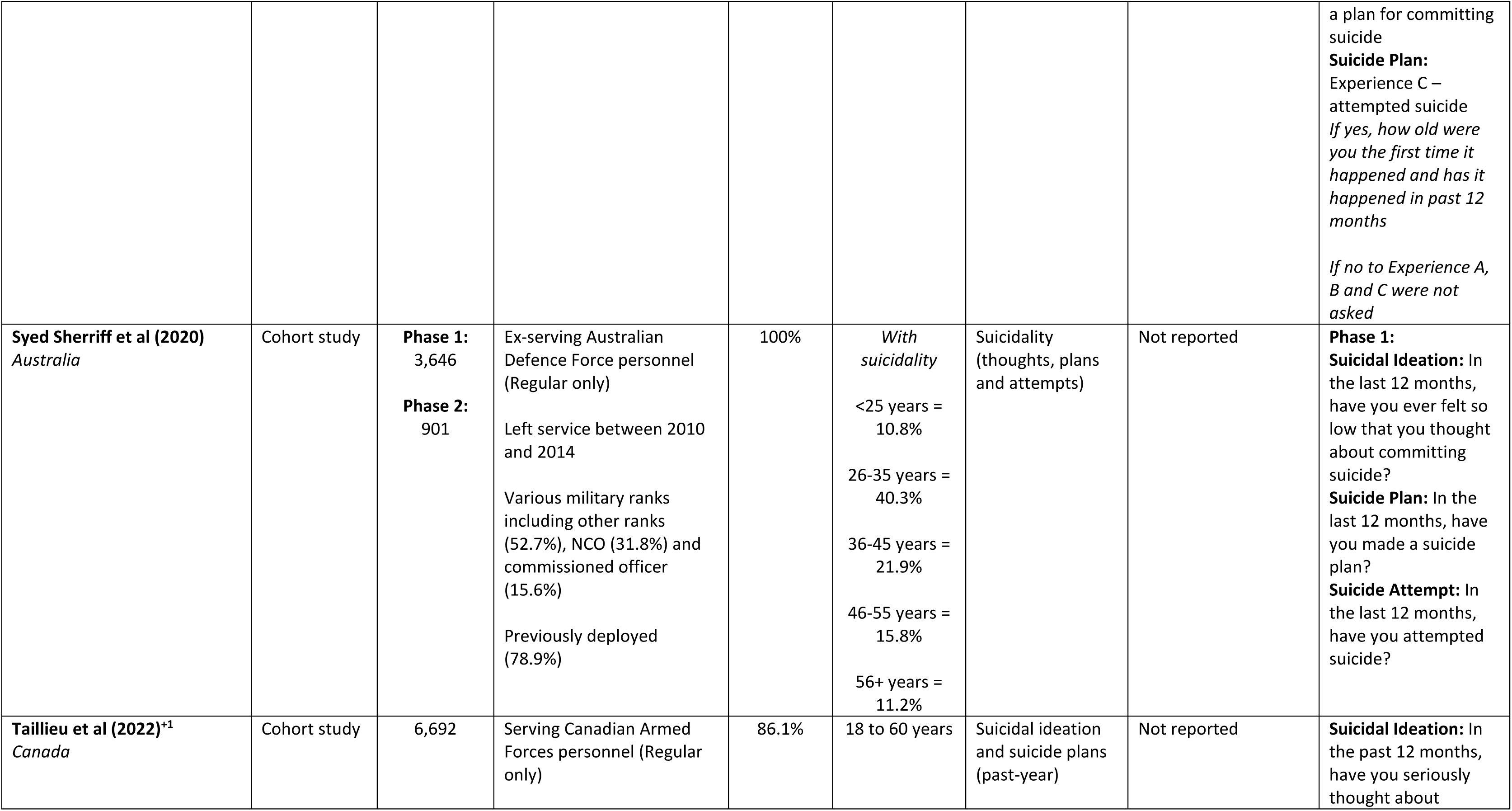
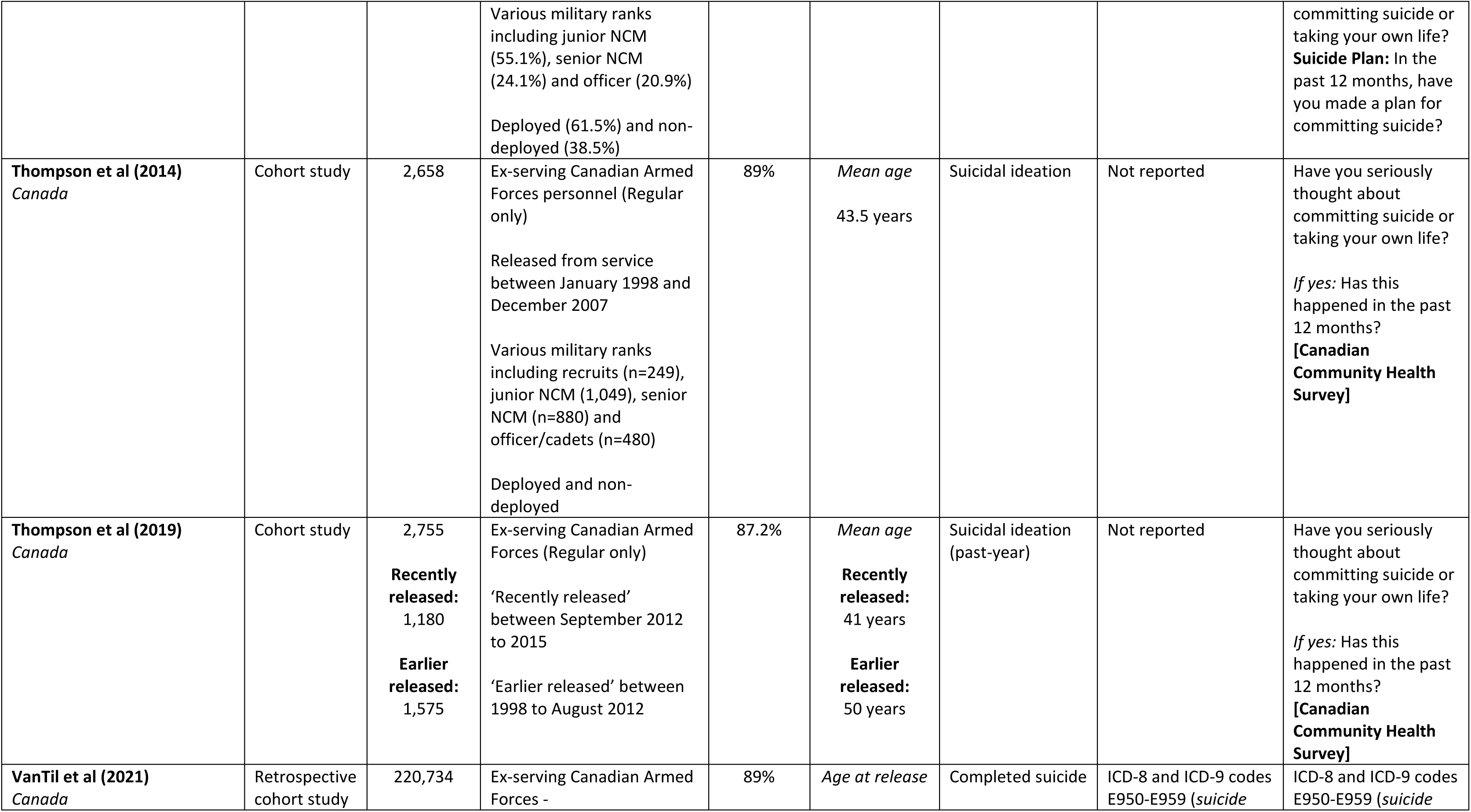
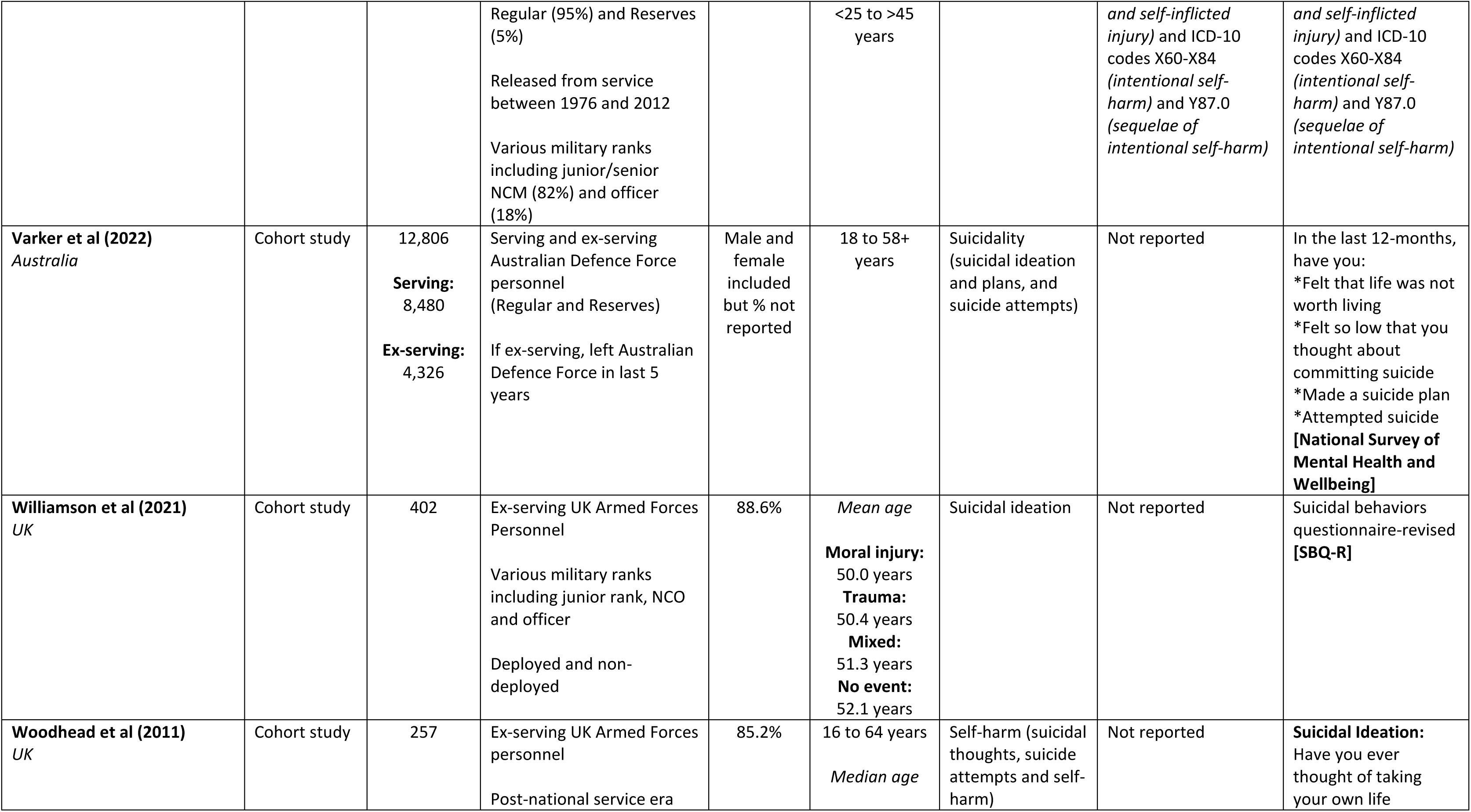
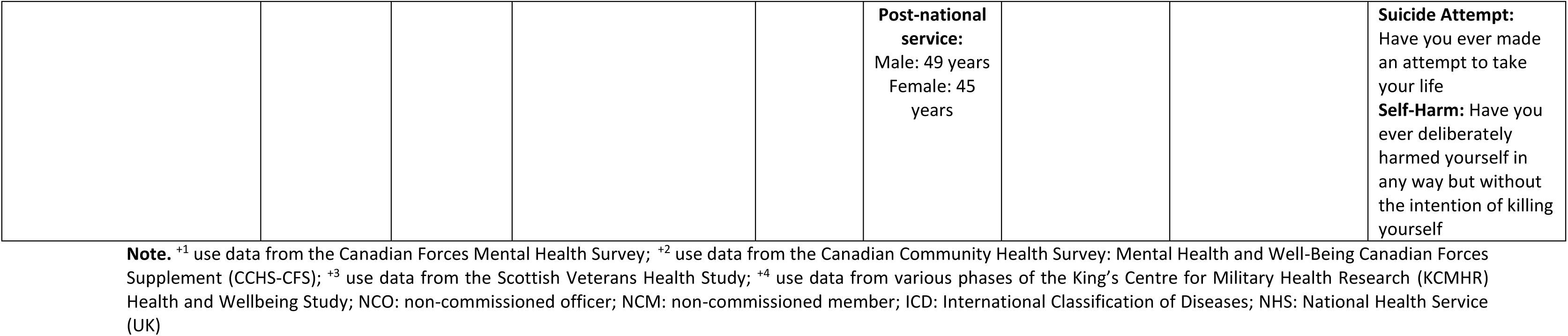
Characteristics of Identified in Included Studies (N=28)

### 3.2. Quality Assessment

Most papers received a quality assessment score of ‘fair’ (n=21), and the rest scored ‘good’ (n=7) (**Supplementary 3).** Common reasons for lower scores included lacking a sample size justification/power calculation, not assessing the exposure variables more than once, and not using outcome measures which were clearly defined, valid and reliable.

### 3.3. Reported Self-Harm and Suicide Behaviour Outcomes

Included papers reported on a range of outcomes outlined in **Table 1**. Four papers reported on self-harm only [15, 53–55], nine reported on suicidal ideation only [36–38, 44–46, 51, 52, 58], two on suicide attempts only [28, 50] and three on completed suicides only [56, 57, 59]. The remaining 10 papers reported on a mix of these outcomes, typically referred to as ‘suicidality’ (often including outcomes of suicidal ideation, suicide plans and suicide attempts) [18, 27, 39–43, 47–49].

#### 3.3.1. Definition and Measurement of Self-Harm and Suicide Behaviour Outcomes

Only six papers provided a definition for the outcomes of interest **(Table 1)**, with the remaining papers failing to report why these terms were not defined.

The majority of papers used self-report surveys (n=17) [15, 28, 36–40, 43, 46, 48–51, 53, 54, 58]. Two papers collected data via structured clinical interviews [44, 45] and four utilised a mix of self-report surveys and structured clinical interviews [18, 27, 41, 42]. One study employed clinical assessment [52]. The remaining papers used ICD codes to define and measure the outcome of interest [55–57, 59].

A variety of outcome measures were used to collect data on the outcomes of self-harm and suicidal behaviours **(Table 1).** For instance, ICD codes [55–57, 59], Patient Health Questionnaire-9 [36, 58], Clinical Interview Schedule-Revised [15, 18] and Suicidal Behaviors Questionnaire-Revised [46].

### 3.4. Risk Factors

Numerous risk factors were identified in the review **[Table 2].**

**Table 2.**
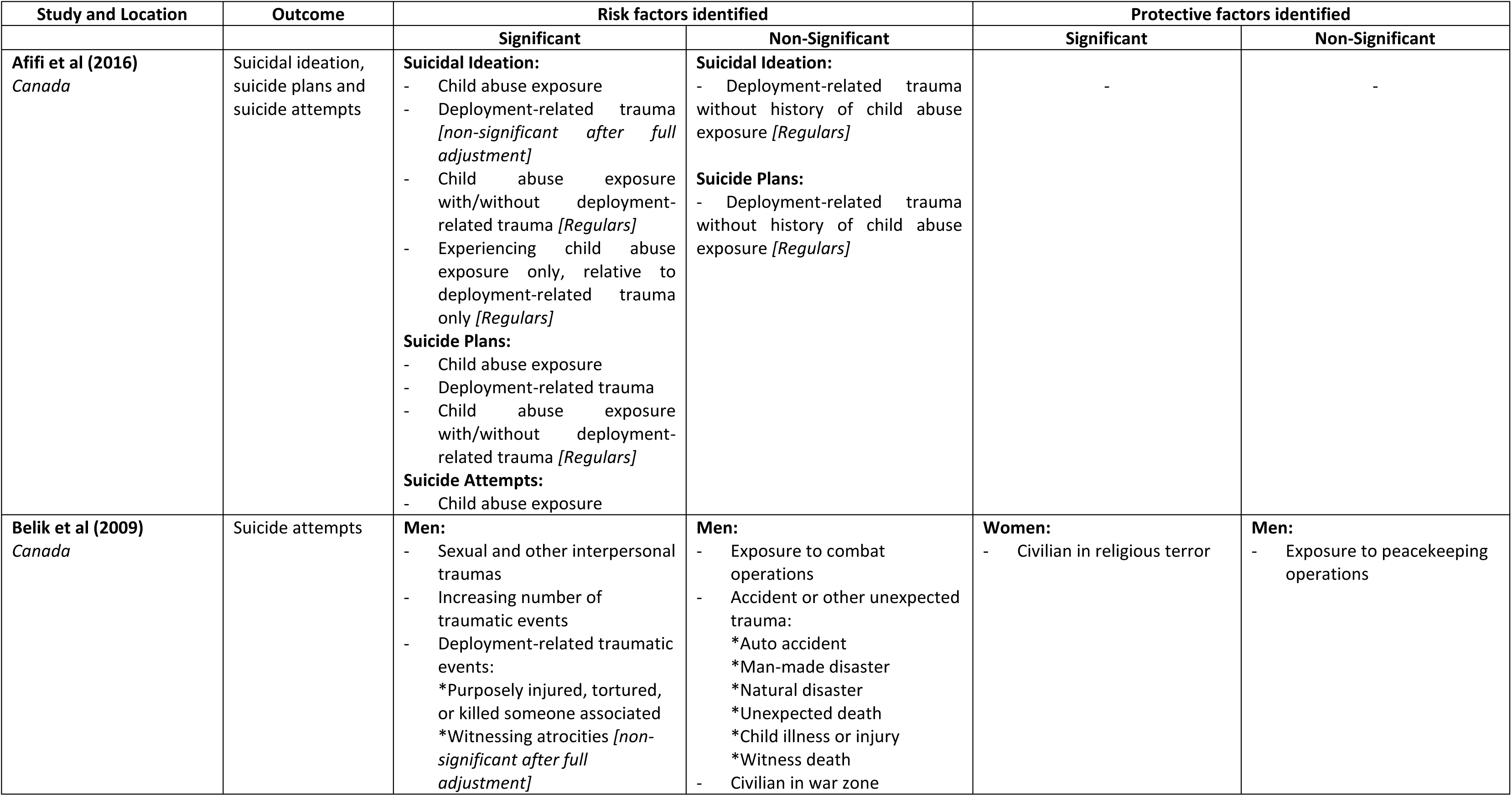
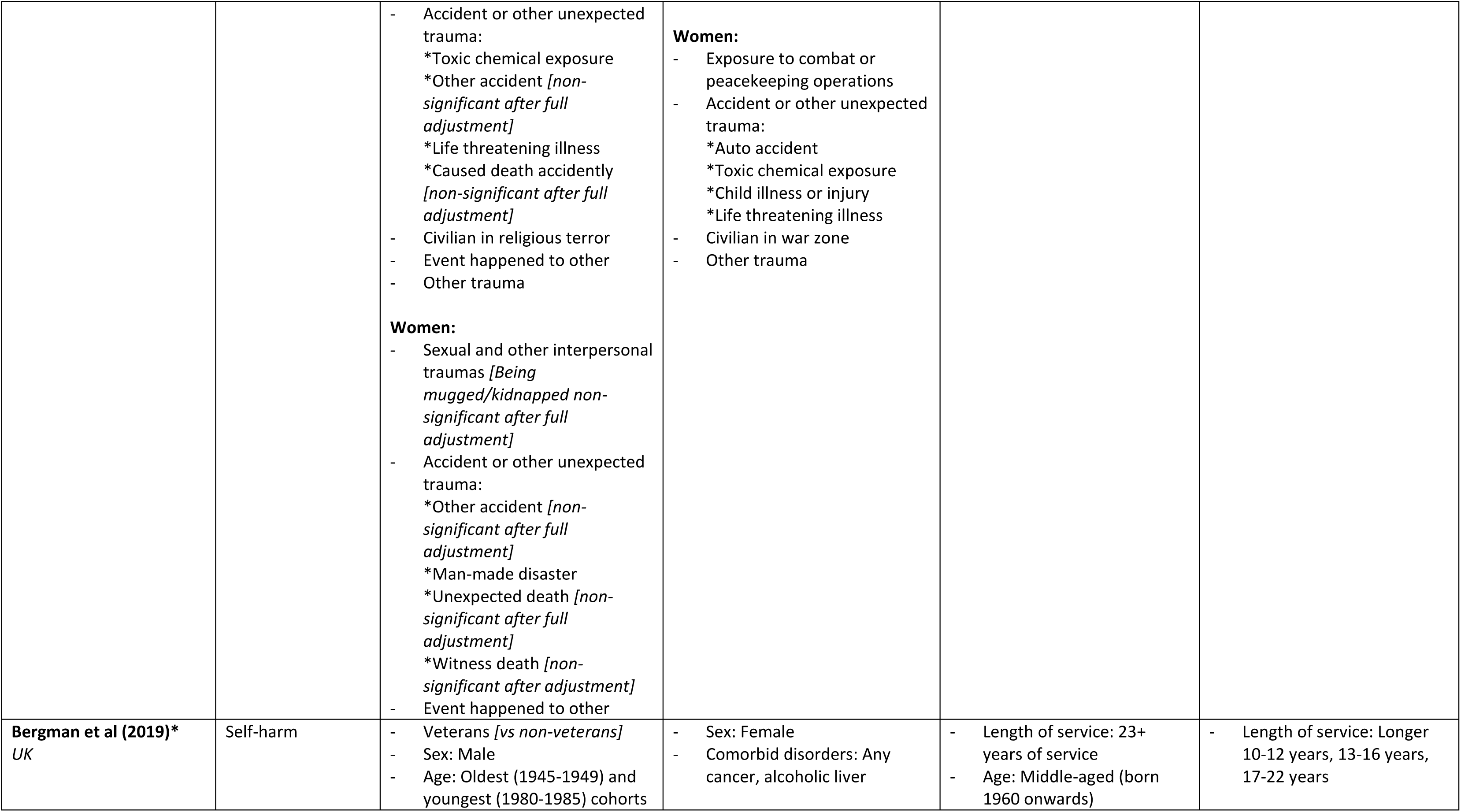
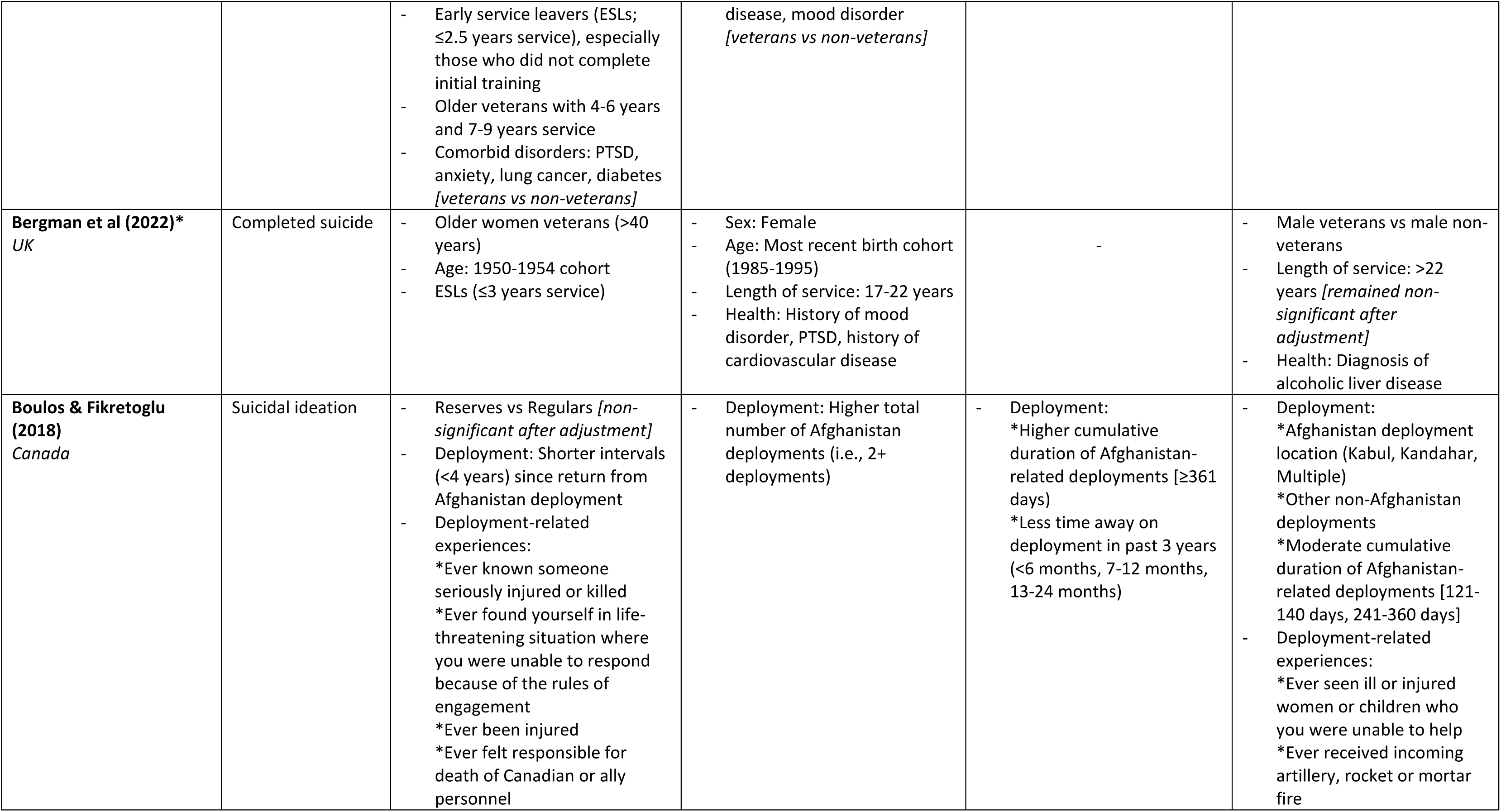
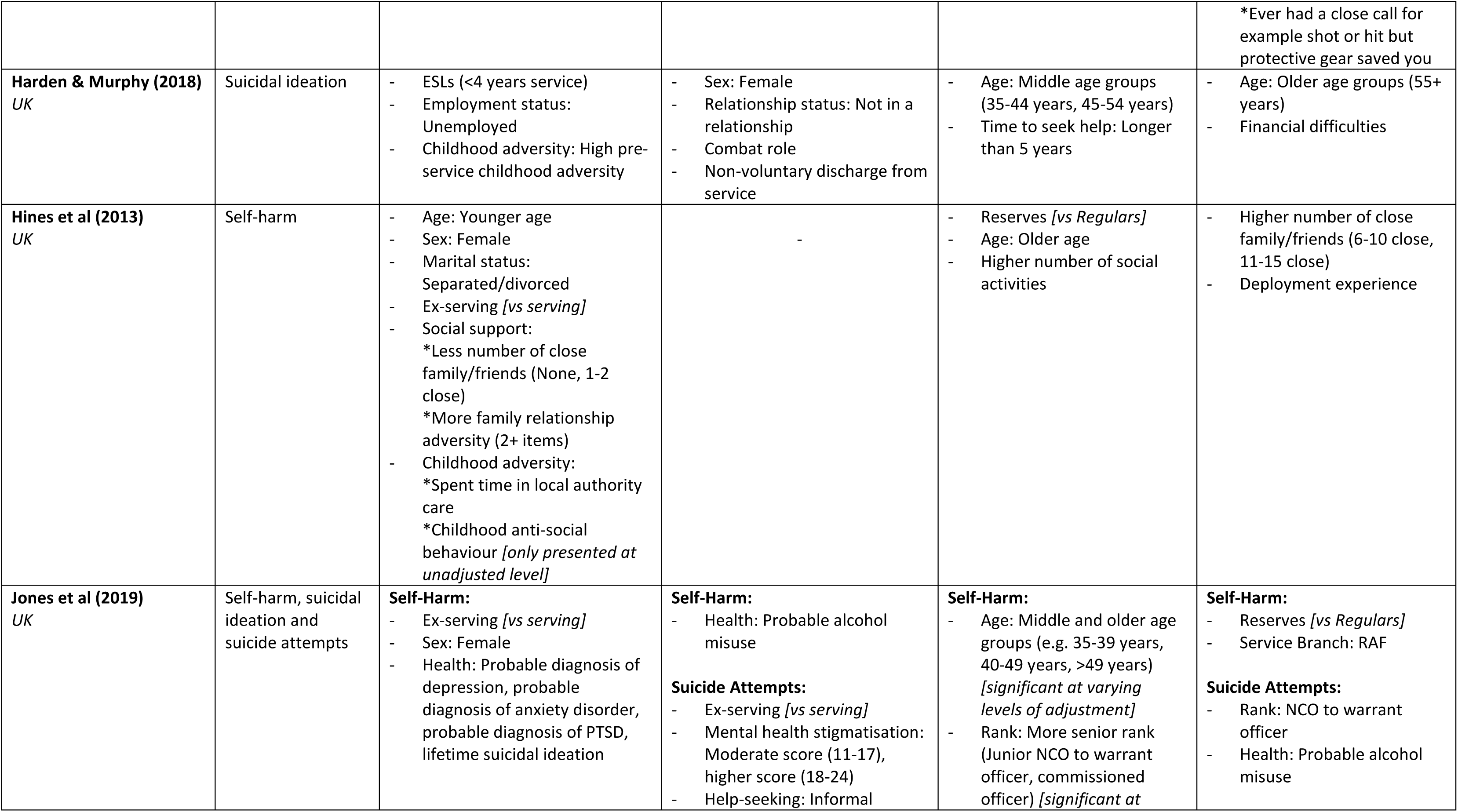
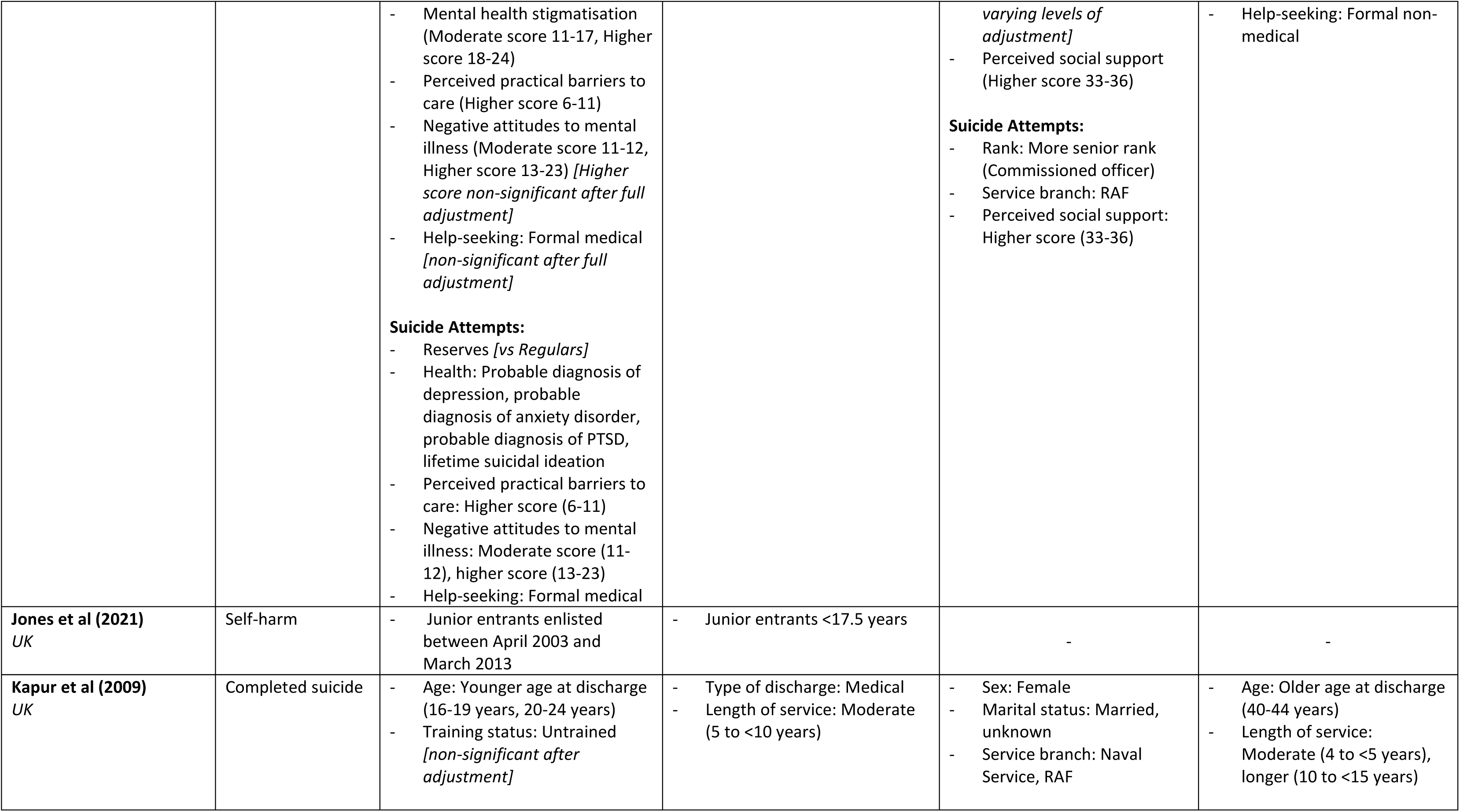
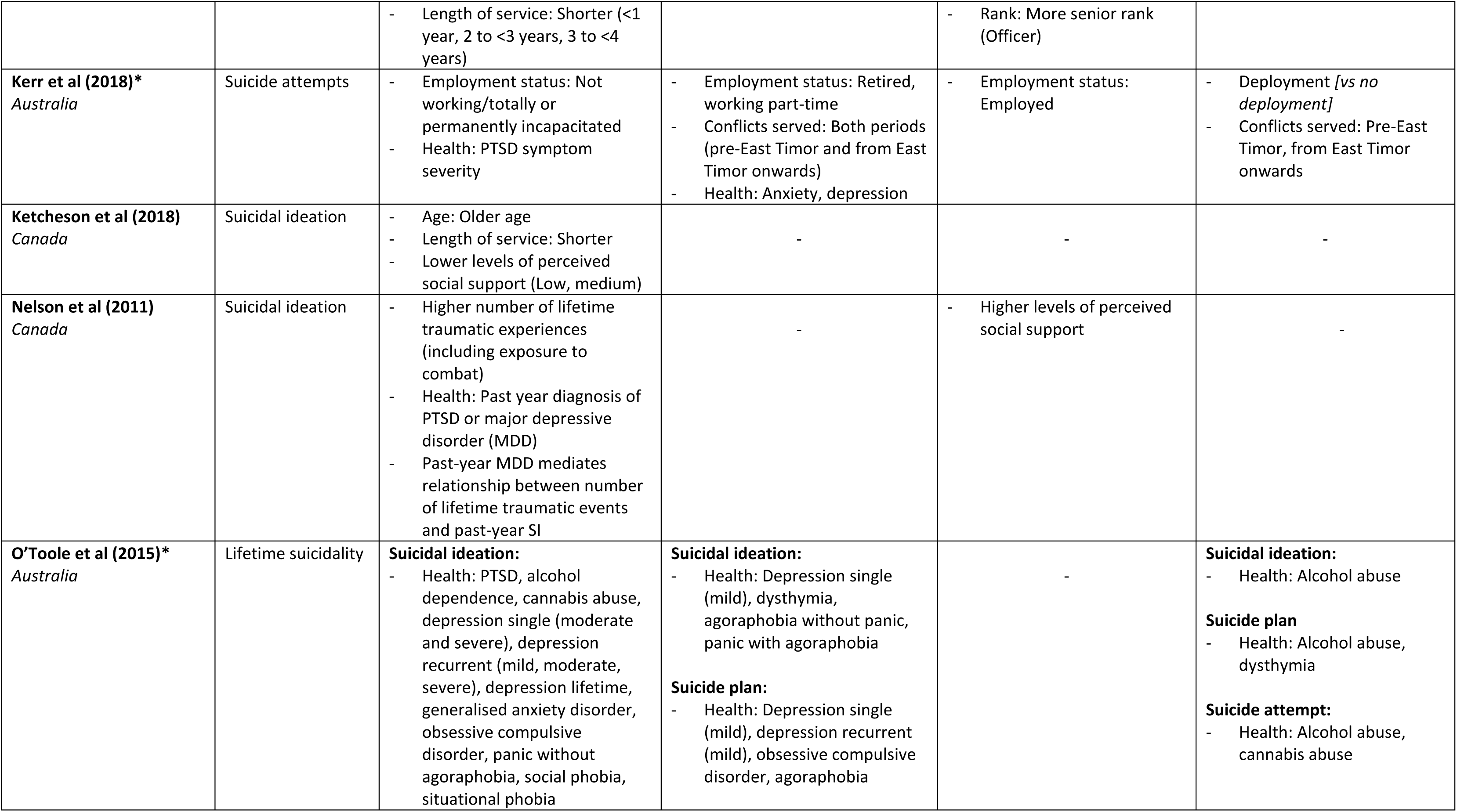
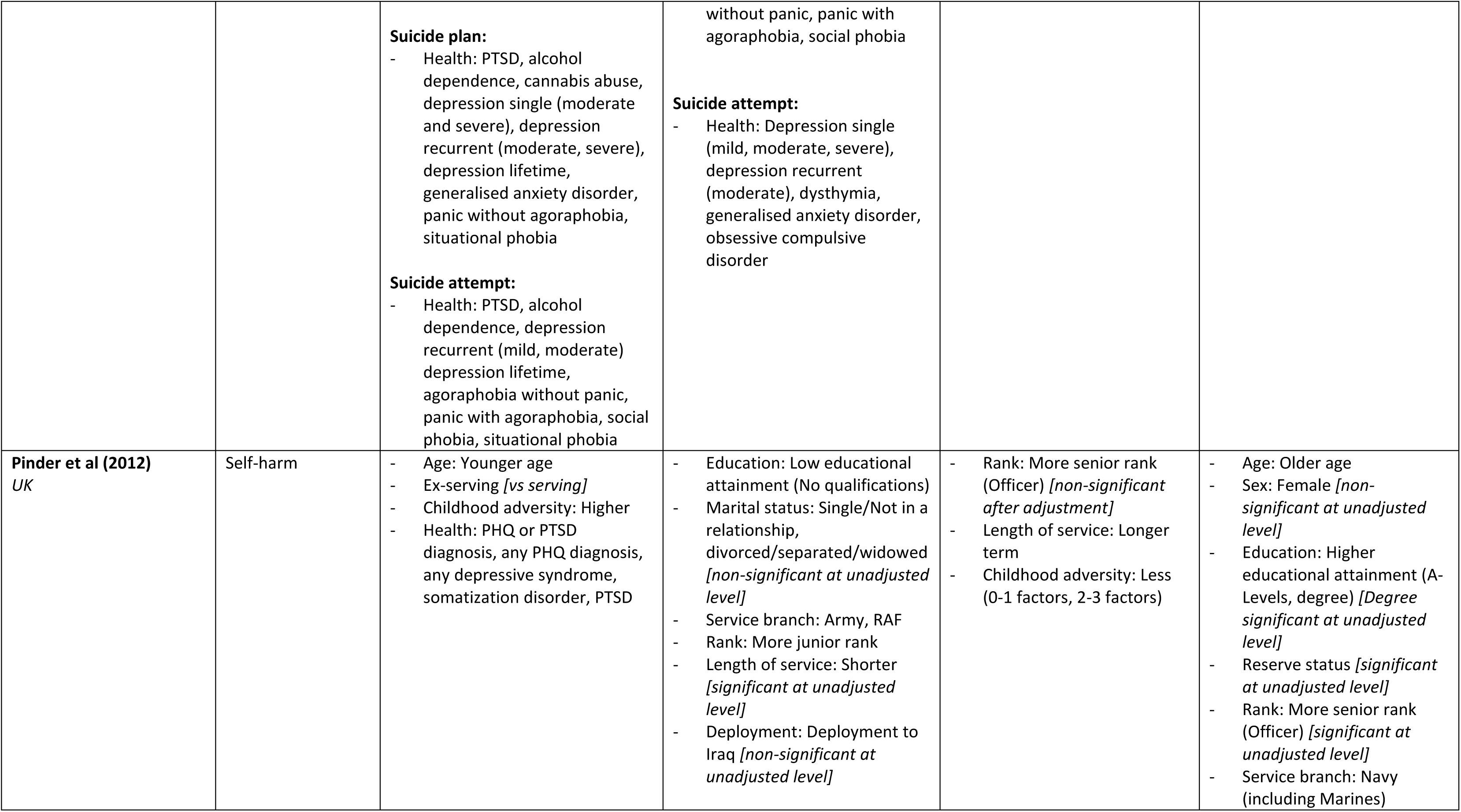
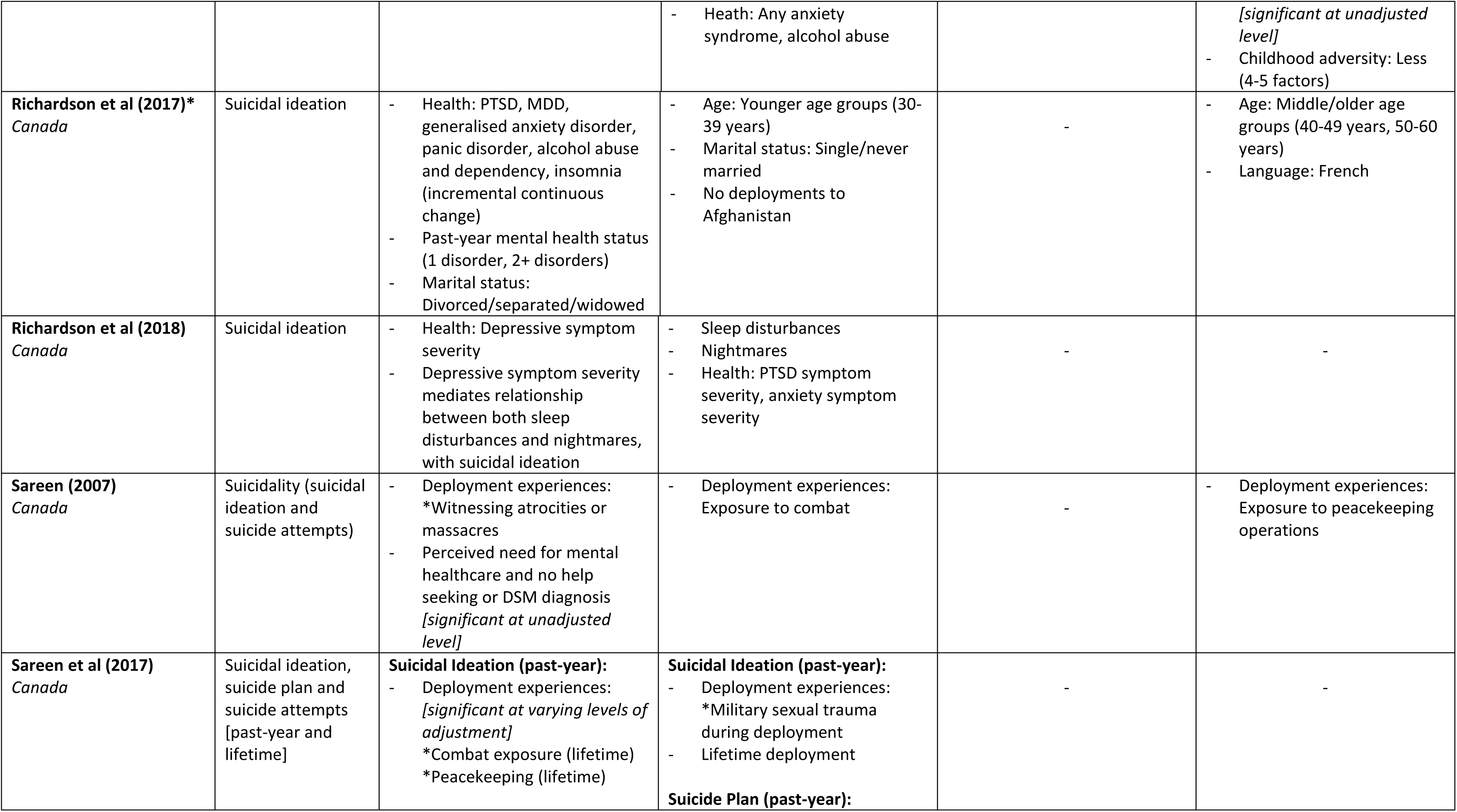
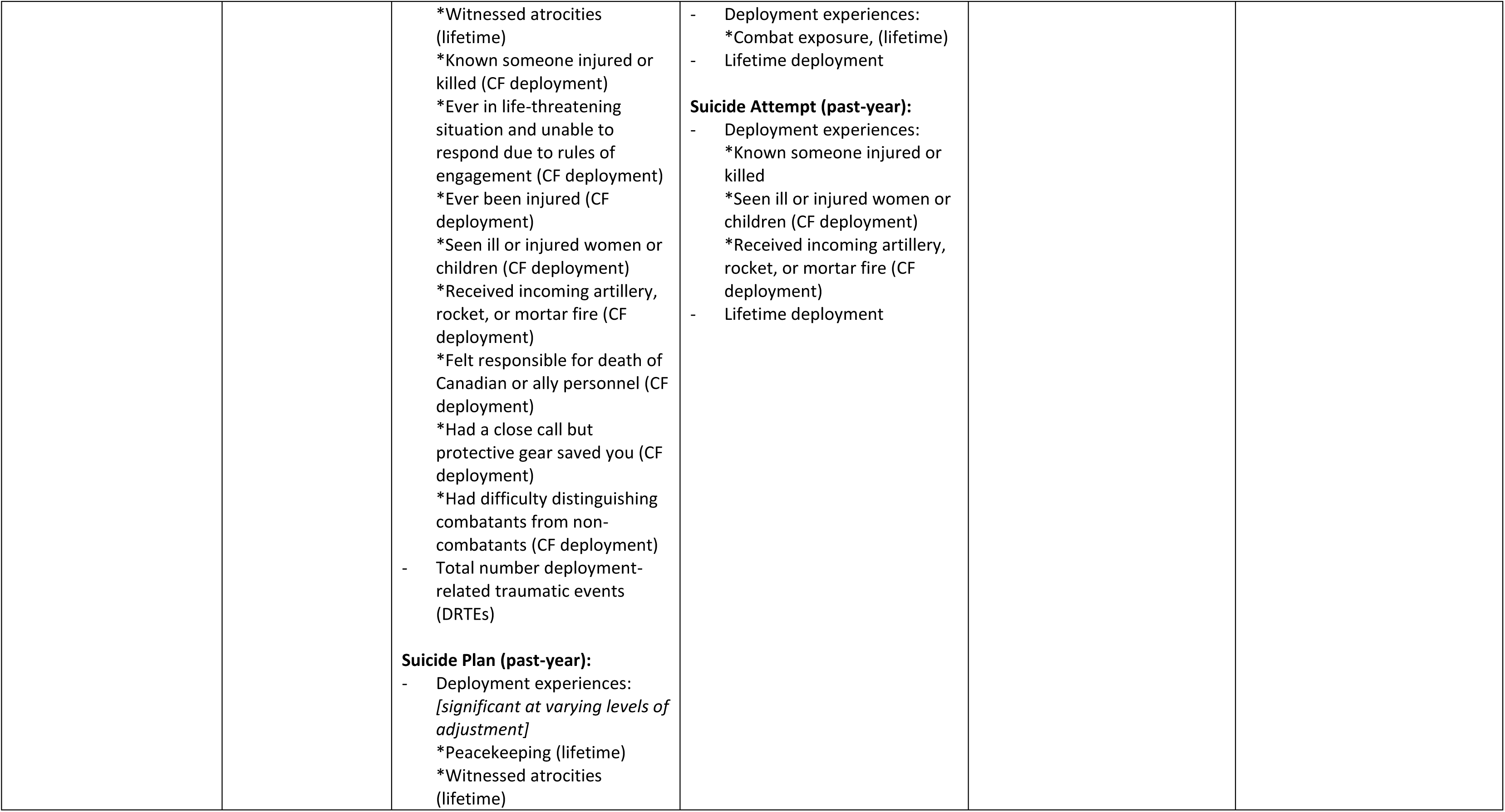
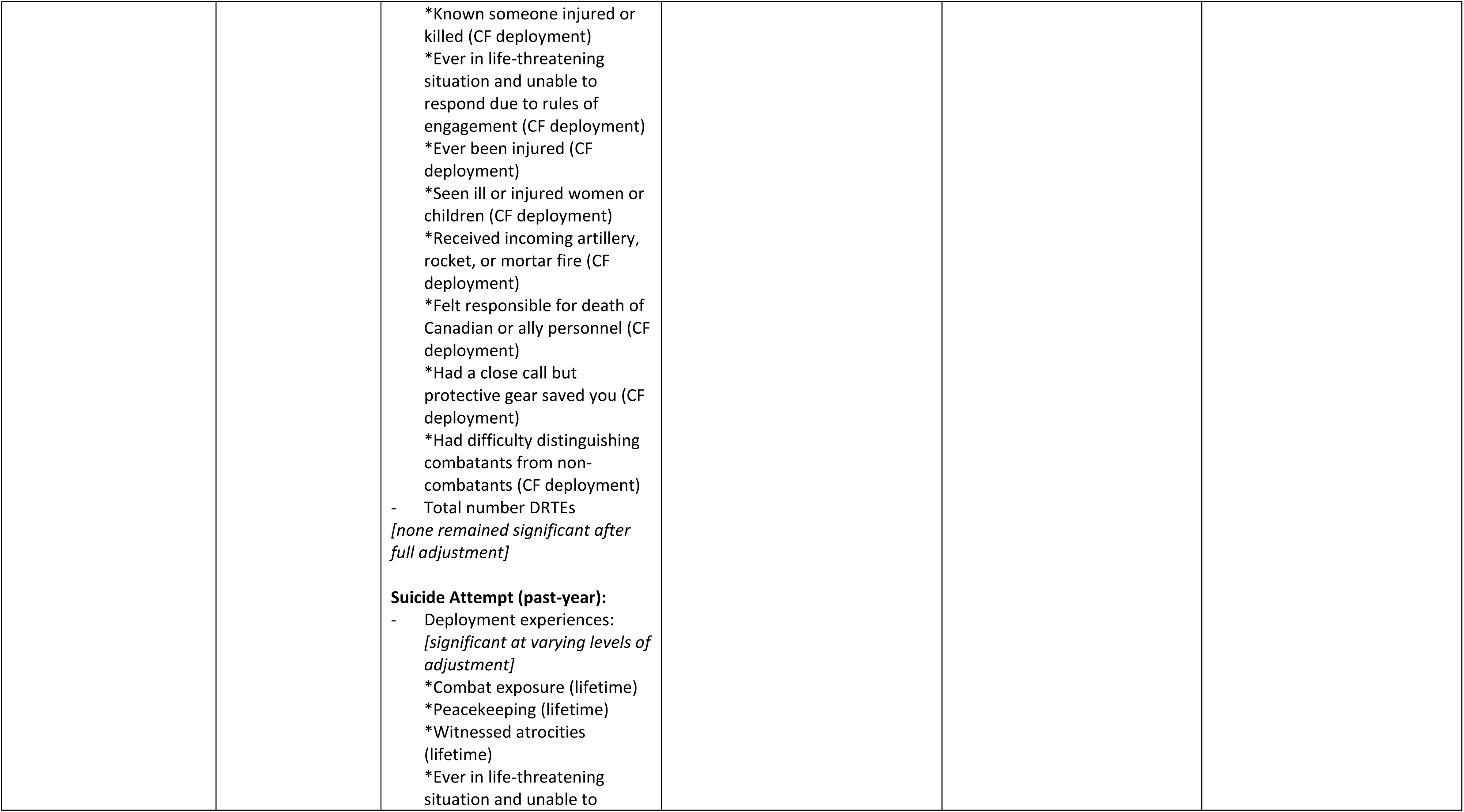
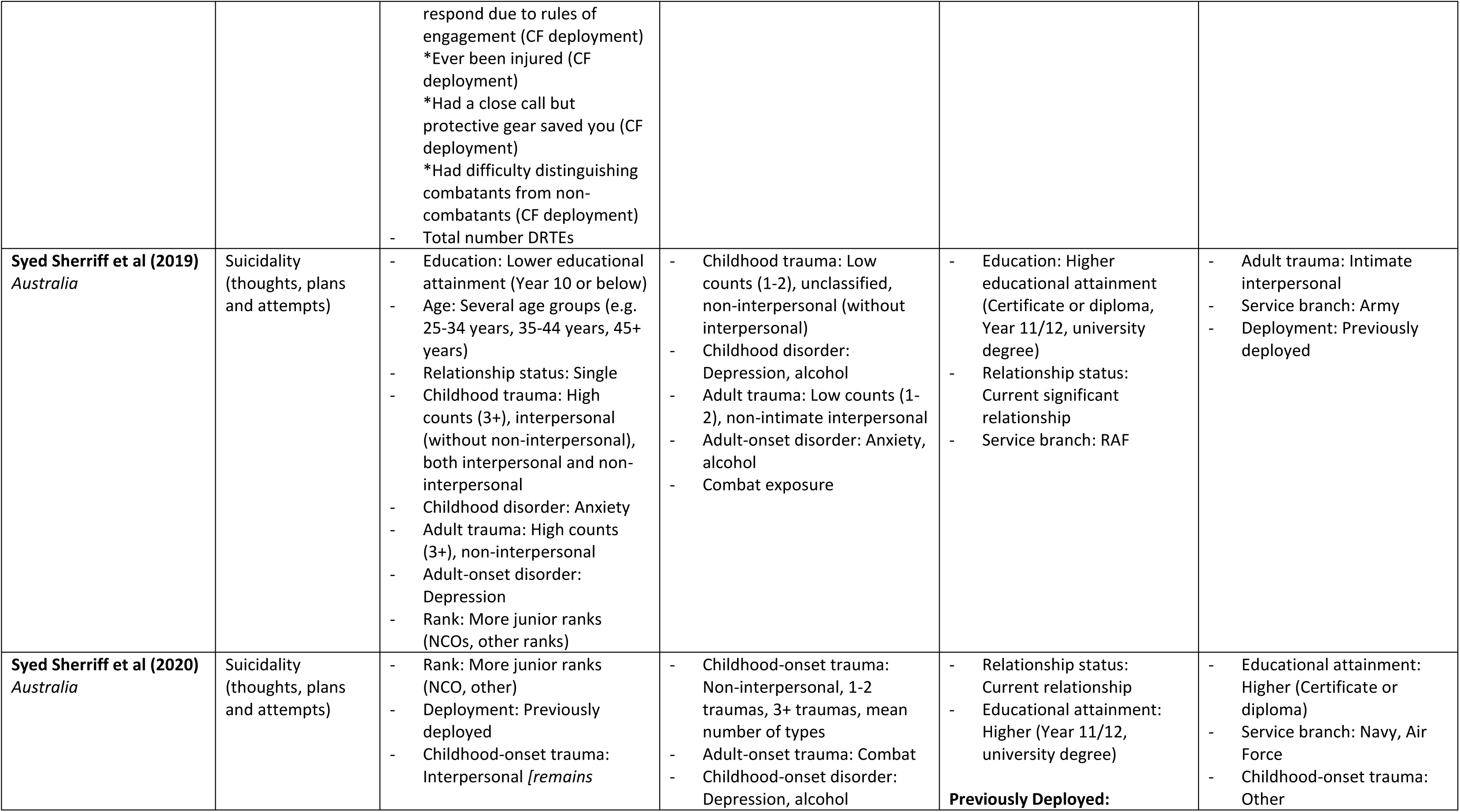
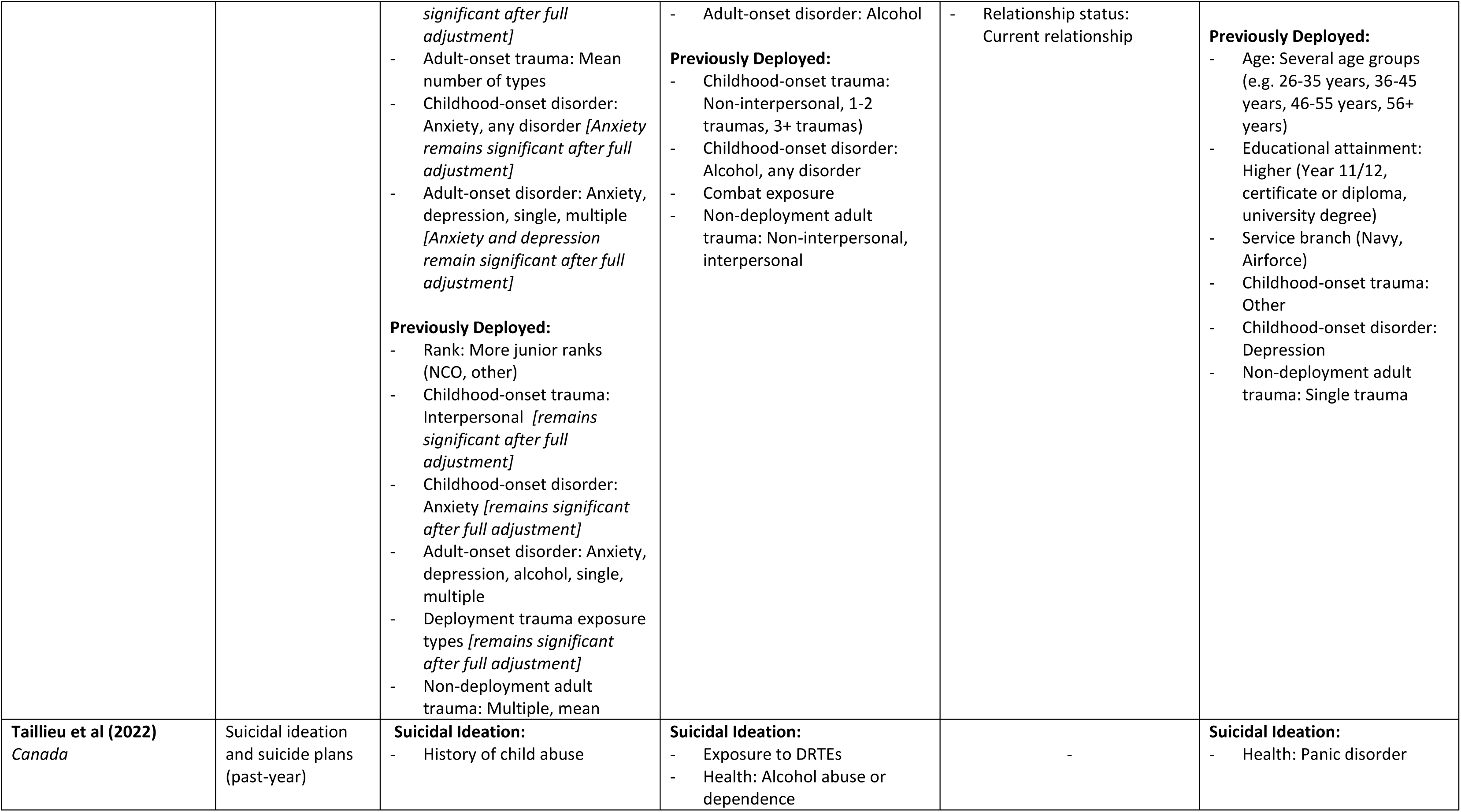
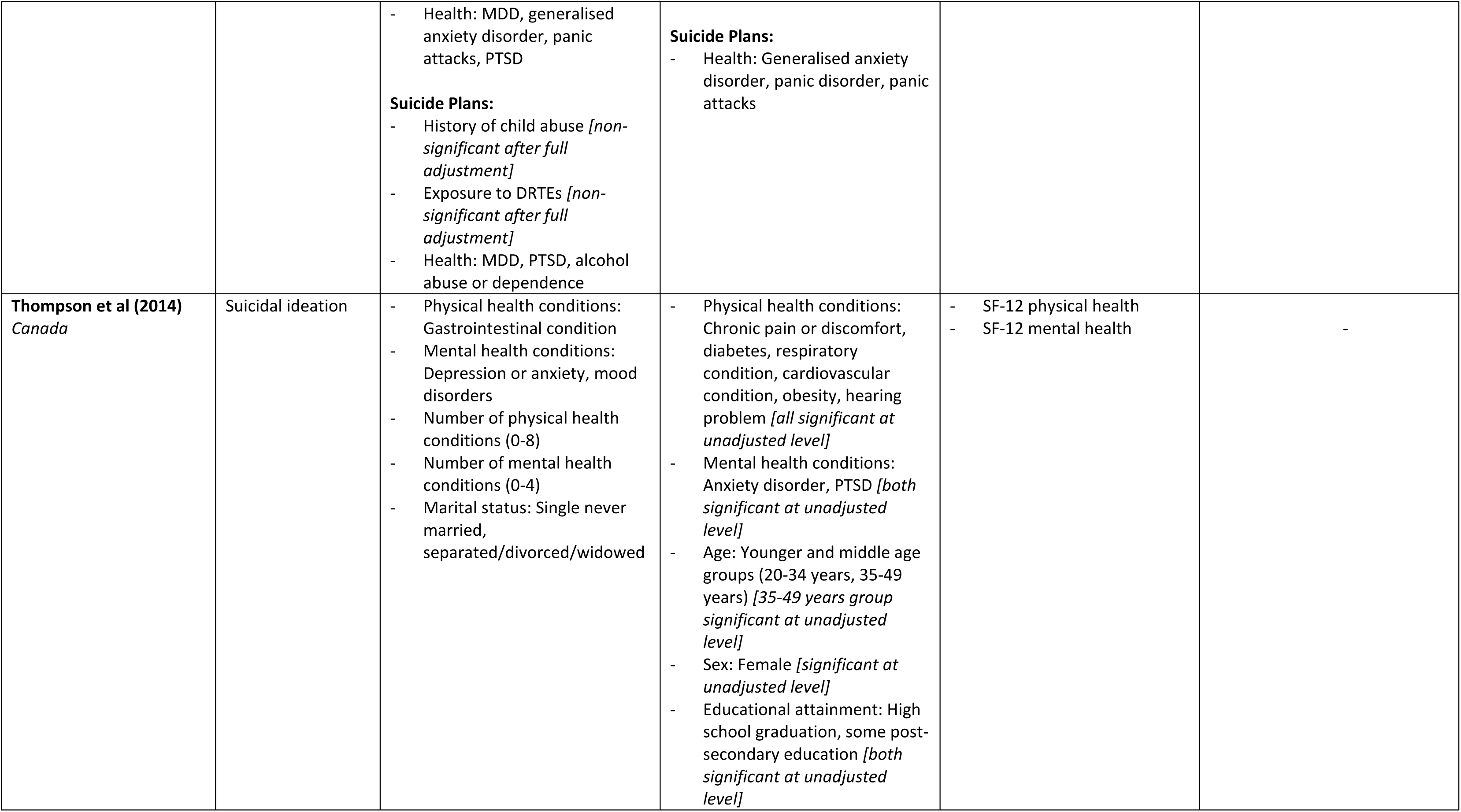
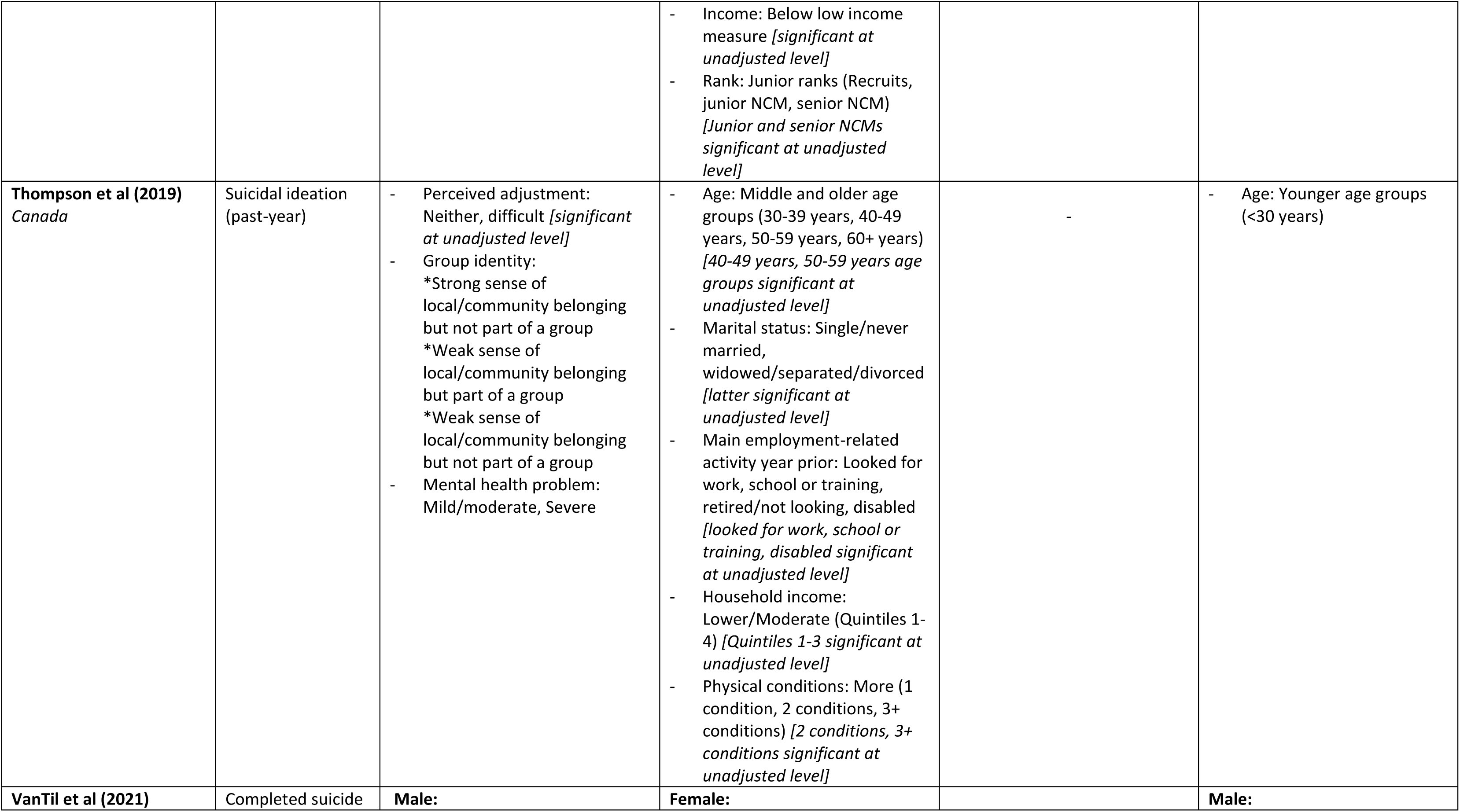
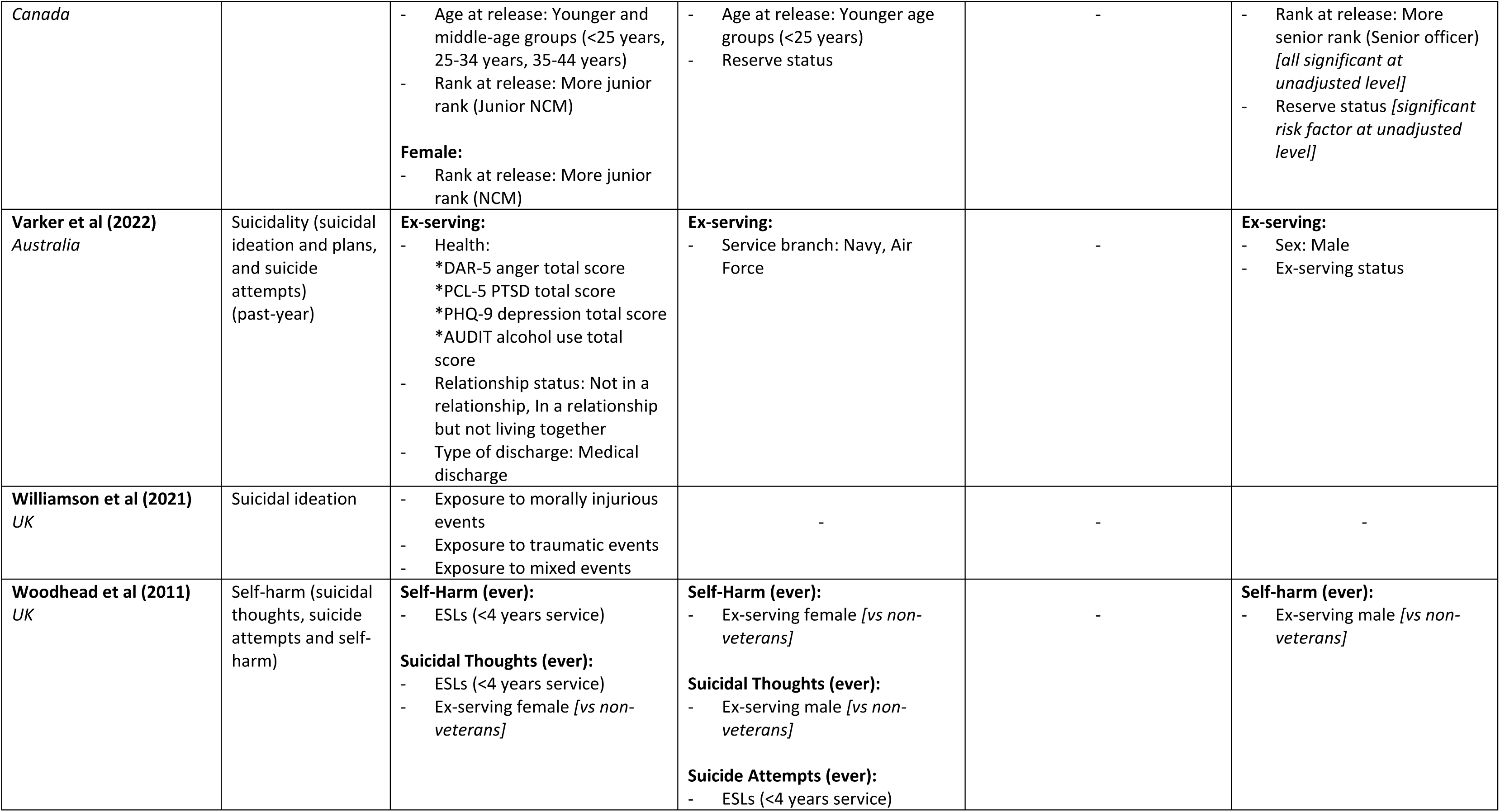
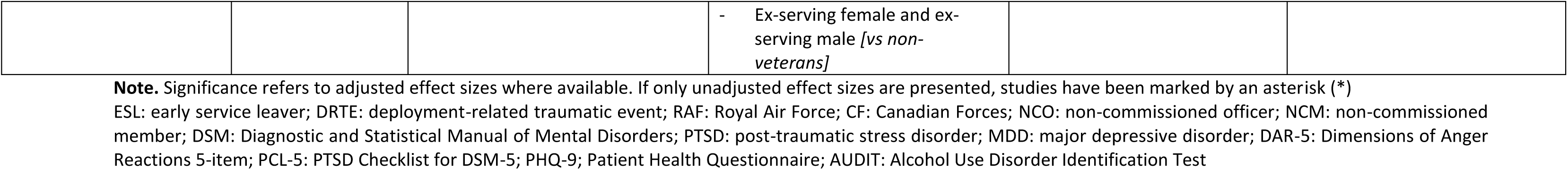
Risk and Protective Factors Identified in Included Studies (N=28)

#### 3.4.1. Risk Factors for Self-Harm

Pre-enlistment factors increased the risk of self-harm, including experiencing childhood adversity and abuse than those without these experiences (approximately doubling the risk) [15, 53]. Certain demographic groups were also at heightened risk including younger age groups [15, 53, 54] compared to middle/older age groups and those no longer in a relationship (i.e., separated/divorced) [53] compared to in a relationship. There were discrepancies between studies regarding the impact of sex, with two studies reporting the risk of self-harm was increased for females [18, 53] but one study stated increased risk for males [55]. Military characteristics associated with increased risk of self-harm included having left service, with ex-serving personnel reported to be as much as three times more at risk of self-harm than serving personnel or non-military population depending on sample explored [15, 18, 53, 55]. Other military characteristics that increased the risk of self-harm included being an early service leaver, defined within the relevant papers as <4 years [47] and ≤2.5 years [55] of service.

Several health-related factors also increased risk of self-harm. The factors with some of the largest effect sizes (ranging from approximately two to eight times more likely) included clinical or probable diagnosis of PTSD [15, 18, 55], depression [15, 18], and anxiety [18, 55], as well as history of suicidal ideation [18]. Other risk factors for self-harm included a lack of social support [53], mental health-related stigmatisation [18], perceived practical barriers to care [18], negative attitudes to mental illness [18], and formal medical help-seeking [18] (all approximately tripling risk).

#### 3.4.2. Risk Factors for Suicide Behaviours

Similar risk factors were identified for suicide behaviours (i.e., suicidal ideation, suicide attempts and completed suicide). Pre-enlistment factors, such as experience of childhood adversity and abuse were found to increase the risk of suicide compared to those without these experiences (around two to seven times more likely) [41–43, 49, 52]. Certain demographic characteristics increased the risk of suicide behaviours, including being single or no longer in a relationship (i.e., separated, divorced, widowed) [38, 41, 44, 48], lower educational attainment (less than university degree level) [41], and unemployment [28, 52]. Another identified risk factor for suicide behaviours was a lack of social support [36]. The influence of age remains unclear, as studies reported a variety of age groups to be at increased risk of suicide behaviours [36, 41, 56, 57, 59]; for instance younger age groups (e.g., 16 to 24 years [57]), younger/middle age groups (e.g., <25 to 44 years [59]), and a mixture of age groups (i.e., 25 to 45+ years [41]).

Numerous military characteristics increased the risk of suicide behaviours, including being medically discharged from service [48], having a shorter length of service (i.e., one to four years [57]) [36, 57], holding a more junior rank during service [41, 42, 59], serving as a Reservist (vs Regular) [18], and shorter intervals since return from last Afghanistan deployment (i.e., <4 years) [51]. Being an early service leaver was another factor that increased risk of suicide behaviours, defined within the relevant papers as ≤3 years [56] and <4 years [47, 52] of service (e.g., increased risk by as much as eight-fold [52]). Experience of deployment-related traumatic events [37, 39, 40, 43, 49–51] and exposure to trauma [37, 41, 42, 46, 50] were other factors associated with increased risk of suicide behaviours.

Several health-related factors were positively associated with suicide behaviours among serving and ex-serving personnel. These included number of physical health disorders (small effect size of just over one) [44] and number of mental health disorders (odds ranging from around two to 20 times more likely) [38, 44, 45]. More specifically, clinical or probable diagnosis of PTSD [18, 27, 28, 37, 38, 43, 48], depression [18, 27, 37, 38, 41–44, 48, 58], anxiety [18, 27, 38, 42, 43, 48], alcohol use disorder [27, 38, 43, 48], cannabis use [27], mood disorder [44], insomnia [38], or panic attacks [43] (ranging from just over one to 15 times more likely). Other risk factors included a higher number of perceived practical barriers to care, negative attitudes to mental illness, and formal medical help-seeking [18]. Further, lifetime suicidal ideation was reported as a risk factor for suicide attempts (up to 12 times more likely) [18].

### 3.5. Protective Factors

A number of protective factors were also identified **[Table 2]**.

#### 3.5.1. Protective Factors for Self-Harm

Several factors were identified that decreased the risk of self-harm, including middle/older age groups [18, 53, 55] (e.g. 35 to 49+ years [18]) compared to other age groups, and experience of less adversity during childhood (i.e., less than three factors) [15]. Certain military characteristics reduced the likelihood of self-harm, for example holding a more senior rank (vs junior ranks) [15, 18], being a Reservist (vs Regular) [53], and having a longer length of service [15, 55]. Further, a higher level of perceived social support was associated with a lower likelihood of self-harm [18, 53].

#### 3.5.2. Protective Factors for Suicide Behaviours

When considering suicide behaviours (i.e., suicidal ideation, suicide attempts and completed suicide), several identified factors reduced risk. Certain demographics were significantly associated with decreased risk of suicide behaviours including being female [57], middle age groups (e.g. 35 to 54 years compared to those <35 years) [52], having a current significant relationship (i.e., being married or in a relationship approximately half as likely than those no longer in relationships) [41, 42, 57], higher educational attainment [41, 42], and being employed [28].

Several military characteristics decreased the risk of suicide behaviours, including holding a more senior rank (vs junior ranks) [18, 57], service in the Royal Air Force [18, 57] or Naval Services [57] (vs Army), and certain deployment-related factors such as higher cumulative duration of Afghanistan-related deployments (i.e., ≥361 days) [51], and less time away on deployment in the past three years (i.e., up to two years) [51] (all approximately half as likely). Higher levels of perceived social support [18, 37] and taking longer than five years to seek support (vs less than five years) [52] were also identified factors that reduced the likelihood of suicide behaviours.

## 4. DISCUSSION

This review identified 28 papers reporting on a range of factors associated with self-harm and suicide behaviours among serving and ex-serving personnel of the UK Armed Forces, Canadian Armed Forces, Australian Defence Force and New Zealand Defence Force.

A variety of definitions and measurements were used for the outcomes reported in the included papers. Definition is an important aspect of academic research and clinical practice, yet precise definitions have been contested, and current terminology varies across nations [61]. For instance, there are several terms for self-harm, including non-suicidal self-injury, deliberate self-harm and self-inflicted violence. Definitions of mental health conditions and the use of consistent language are an important starting point within good quality research papers and are also important for reducing stigma and encouraging individuals to seek help, particularly relating to self-harm and suicide behaviours [62].

Several of the identified risk factors reflect those among the general population and similar occupational groups (such as emergency responders) [3, 63, 64], and were generally consistent across the included geographical regions. Although no relevant peer-reviewed papers were identified from New Zealand, suicide prevention work is being conducted [65]. Risk factors for suicide among the New Zealand Defence Force include current mental health concerns, acute life stressors and negative attitudes towards help-seeking, and protective factors include positive mental health and social support [65]. The paucity of peer-reviewed work from New Zealand may be due to the smaller military population of the New Zealand Defence Force compared to the military populations of other included nations, making large quantitative studies more difficult.

It is important to recognise that some risk and protective factors were not always consistent across included studies in this review, and the influence of age, sex, certain military service characteristics, and certain health diagnoses remains unclear. For instance, having served as a Reservist increased the likelihood of suicide behaviours [18] but decreased the likelihood of self-harm [53], highlighting the need for a holistic approach when supporting military communities. It is important to develop an enhanced understanding about motivations for engaging in self-harm and suicide behaviours, as for some self-harm acts as a coping mechanism to regulate internal emotions, whereas suicide behaviours become more apparent when the individual can no longer cope [66].

One key risk factor for self-harm was having left service. Potentially the transition from serving to ex-serving is a period of risk and support should be in place during this time. It might be that after leaving service, ex-serving personnel no longer feel like part of the military ‘family’, experience a shift in their sense of self and have difficulty connecting to civilian life [67]. Alternatively, it could reflect the influence of time as ex-serving personnel are typically older, and therefore, if considering lifetime prevalence and risk, this would capture a longer period of time. Additionally, there may be an underrepresentation among serving personnel as they are potentially more hesitant to report these behaviours due to stigma and fear of negative consequences to their career [68]; personnel with ill health (physical or mental) may be forced to leave service (i.e. medical discharge) and those who involuntarily separate from the military are at high risk of self-harm and suicide behaviours [22, 69, 70].

Poor health was another key identified risk factor, in particular clinical or probable diagnosis of PTSD, depression and anxiety. This highlights the importance of early detection of mental health problems, providing adequate care and support to military personnel throughout their military career, and providing continuity of care as they transition out of service. Interestingly, seeking help from formal medical sources was positively associated with lifetime suicide attempts [18] and taking more than five years to seek help was negatively associated with suicidal ideation [52]. It is unlikely that seeking help from formal medical sources has a causal relationship with suicide behaviours. Instead, UK research has found that ex-serving personnel are known to present to clinical services at times of crisis, which might involve an active episode of self-harm or suicide behaviours, therefore, placing them more at risk [71]. Additionally, those who delay seeking help for longer may have discovered ways to cope with their difficulties on their own, whereas those who seek help sooner may have been in crisis and at higher risk of suicidal ideation [52]. Clearly, a key challenge relating to self-harm and suicide behaviour risk is prevention. Most developed nations already have suicide prevention strategies in place, including for serving and ex-serving personnel, or as part of the wider mental health strategy [72–75]. The findings of this review suggest that prevention and intervention strategies should focus on the broader context of improving health and wellbeing (physical, mental, and social health).

Less research focussed on protective factors for self-harm and suicide behaviours among the included military populations. Despite this, one key association was with higher levels of perceived social support [18, 37], indicating that this was likely a salient factor that could help prevent or mitigate risk, particularly due to its modifiable nature. This aligns with other international military populations, including the US [76], as well as in the general population where social support has been recommended for use in suicide prevention strategies [77]. One way to bolster social support among military populations is the use of peer support as a preventative strategy [78, 79].

### Strengths and Limitations

A strength of this review was the search of seven literature databases using a broad search strategy outlined in an a priori PROSPERO approved review protocol. The protocol was generally followed but any changes were reflected by updating the protocol (i.e., removing the limit around the age of the sample). Additionally, a proportion of the eligibility assessment and critical appraisal of papers was conducted by a second, independent reviewer with high inter-rater reliability. Including a wider range of geographical regions may have led to more included papers (e.g., Na et al (2021) [80]), however focus this review aimed to address the literature gap by collating evidence where attention has previously been limited.

There were some limitations to note. As with all systematic reviews, the findings of this review were subject to publication bias. Additionally, some potential associated factors received less attention than others, however this review tried to provide the best synthesis of the evidence currently available.

There were also several limitations relating to the included papers. First, papers used different definitions and measurements of self-harm and suicide behaviours, limiting the possibility of comparing findings across studies. However, combining these papers in a review contributes to understanding. Second, there is no universally agreed definition of veterans (i.e., ex-serving personnel) which makes cross-cultural comparison difficult. Third, studies did not always report on some military characteristics (such as service branch, rank, deployment experience, era of service and time since leaving service) and demographic characteristics (such as ethnicity and sexuality) which would have been useful for contextualisation and interpretation of the findings. There remains limited understanding of the impact of sex, experiences of ethnic minority and LGBTQ+ personnel. Fourth, included studies used a variety of sample sizes (range n= 144 to 233,803), study procedures, and sample characteristics. For example, the majority of papers relied on self-reported data which may have been subject to recall bias and social desirability bias.

### Implications

This review has several important implications for policy, practice, and research. The identification of risk and protective factors can be useful to inform military health services and policies including where to target suicide prevention policy to reduce the incidence and impact of suicide. In the UK, this is one of nine key health priority themes laid out in the Defence People Health and Wellbeing Strategy 2022-2027 [81].

Identifying the risk and protective factors for self-harm and suicide behaviour outcomes is an important aspect of the development and implementation of effective prevention and intervention strategies to protect the mental health and wellbeing of military populations. Evidence on associated factors can inform healthcare practice and service provision for the Armed Forces community by highlighting several at-risk groups which may require additional support. This review suggests that additional support is required during the period of transition from military to civilian life but also highlights the importance of prevention early on in military service to reduce the impact on personnel as they transition out of service. The identified protective factors suggest that prevention and intervention strategies should focus on encouraging help-seeking for mental health problems early on before crisis events occur, as well as promoting social support networks and strengthening connections with family, friends and the community as a whole.

Additionally, focus should be placed on modifiable factors (i.e., factors that could reasonably be altered, such as psychiatric symptoms and social support). Although non-modifiable factors can help to identify level of risk, they are of less use during prevention and intervention as they cannot be changed to alter the level of risk. Therefore, particular focus on modifiable risk and protective factors is warranted as they are amenable to therapeutic intervention and can be key to addressing long-term risk due to their adaptive nature. However, it is still important to understand the role of non-modifiable factors as there may be an indirect effect, for example when considering military rank, it might be that officers are at lower risk of self-harm and suicide behaviours because they typically hold higher socio-economic status and higher educational attainment [82] which are known protective factors.

Future studies should also focus on conducting longitudinal investigations which distinguish between pre-, peri- and post-service factors in order to identify pathways of self-harm and suicide behaviours, and to ensure support is in place at the right point in the military lifecycle.

### Conclusions

This review highlighted several risk and protective factors for self-harm and suicide behaviours which warrant attention. Adequate care and support are a necessity for serving and ex-serving personnel who may be at risk of experiencing self-harm or suicide behaviours. Particular focus should be placed on implementing prevention strategies early on in military service to reduce the impact on personnel as they transition out of service. The identified protective factors suggest that prevention and intervention strategies should promote social networks as a key source of support for military personnel. Whilst this review was limited due to the paucity of peer-reviewed research within some populations, current work, such as that being undertaken in New Zealand will add to the understanding. Research should continue to progress towards understanding and preventing self-harm and suicide behaviours among military populations.

## Data Availability

Data availability is not applicable to this article as no new data were created or analysed in this study.

## STATEMENTS AND DECLARATIONS

### Ethical approval

N/A

### Authors’ Contributions

CW proposed the review. NTF and SAMS secured the funding for Phase 4 of the King’s Centre for Military Health Research Health and Wellbeing Cohort Study. CW, MLS and SAMS contributed to the design of the review. CW completed all stages of screening and data extraction. BC was the second rater for 10% of results at title/abstract screening and 20% at full text screening. CW completed the quality assessment for all included papers and BC performed second rater assessment for 20% of included papers. CW wrote the first draft, and all other authors (BC, AS, NTF, MLS, SAMS) contributed to each revision and approved the final manuscript.

## Acknowledgements

We would like to thank Colonel Clare Bennett, Chief Mental Health Officer New Zealand Defence Force, for sharing relevant work being conducted in New Zealand.

