## Supplementary 1 for "Risk and Protective Factors for Self-Harm and Suicide Behaviours among Serving and Ex-Serving Personnel of the UK Armed Forces, Canadian Armed Forces, Australian Defence Force and New Zealand Defence Force: A Systematic Review"

**Supplementary 1. Concepts and Keywords Used in Database Searches**

| Concept | Keywords |
|---|---|
| **Concept A:**<br><br>**Self-harm and suicide behaviours**<br>*(Ti/Abs)*<br><br>**AND** | self-harm* **OR** "self harm*" **OR** self-injur* **OR** "self injur*" **OR** "self-injurious behavi?r" **OR** "deliberate self-harm" **OR** "non-suicidal self-injur*" **OR** "nonsuicidal self-injur*" **OR** "non suicidal self injur*" **OR** "self-inflicted injur*" **OR** "self-inflicted violence" **OR** "self-destructive behavi?r" **OR** self-violen* **OR** self-poison***OR** self-cut* **OR** suicid* **OR** "suicid* attempt*" **OR** "attempted suicid*" **OR** "suicid* ideation" **OR** "suicid* thoughts" **OR** parasuicid* **OR** "completed suicid*" **OR** "fatal suicid*" |
| **Concept B:**<br><br>**Military**<br>*(Ti/Abs)*<br><br>**AND** | military **OR** "military personnel" **OR** "armed force*" **OR** servicemember* **OR** soldier* **OR** "military service" **OR** servicem?n **OR** servicewom?n **OR** veteran* **OR** ex-servi* **OR** navy **OR** naval servic* **OR** army **OR** **"**air force" **OR** airforce **OR** marine* **OR** "marine corp*" **OR** reserve* **OR** reservist **OR** national guard **OR** infantry* **OR** "military psychiatry" **OR** "military health" **OR** "military deployment*" **OR** deploy* war **OR** warfare **OR** warfighter* **OR** combat **OR** "armed conflict*" **OR** "defen?e force" |
| **Concept C:**<br><br>**Location**<br>*(Anywhere/Full Text)* | "United Kingdom" **OR** UK **OR** "Great Britain" **OR** Britain **OR** British **OR** England **OR** English **OR** Scotland **OR** Scottish **OR** Wales **OR** Welsh **OR** "Northern Ireland" **OR** "North Ireland" **OR** "N Ireland" **OR** Irish **OR** Canada **OR** Canadian **OR** "First Nations" **OR** Métis **OR** Inuit **OR** Australia **OR** Australian **OR** "Aboriginal Australians" **OR** "Torres Straight Islanders" **OR** "New Zealand" **OR** "New Zealanders" **OR** Māori **OR** "Pacific Islanders" |

***Note.*** The Boolean operators 'OR' and 'AND' were used where appropriate. The asterisk (*) denotes truncation and the question mark (?) denotes wildcards.
