## Supplementary 2 for "Risk and Protective Factors for Self-Harm and Suicide Behaviours among Serving and Ex-Serving Personnel of the UK Armed Forces, Canadian Armed Forces, Australian Defence Force and New Zealand Defence Force: A Systematic Review"

**Supplementary 2. Key Findings and Quality Assessment (N=28)**

| Study and Location | Key Findings | Statistical Findings | Quality Rating (Good, Fair, Poor) |
|---|---|---|---|
| **Afifi et al (2016)**<br>*Canada* | - Child abuse exposure is more likely among military personnel compared with the general population in Canada<br>- Child abuse exposure is associated with increased odds of suicidal ideation, suicide plans and suicide attempts among the CGP and CAF (Regular and Reserve Forces). However, the effect of child abuse exposure on suicide-related outcomes is generally stronger among the CGP compared with military personnel<br>- Deployment-related trauma was associated with past-year suicidal ideation and suicide plans, however, in comparison, child abuse exposure was more strongly and more consistently associated with suicide-related outcomes<br>- Prevention efforts targeting child abuse may reduce suicide-related outcomes | - Child abuse exposure associated with increased odds of suicidal ideation, suicide plans, suicide attempts (range AORs 1.7 [95% CI 1.0-2.9] to 6.3 [95% CI 4.2-9.5], *p*<.05)<br>- Deployment related trauma was associated with past-year suicidal ideation (AOR 1.4 [95% CI 1.0-1.8], *p*<.05) and suicide plans (AOR 1.7 [95% CI 1.1-2.7], *p*<.05) *[suicidal ideation model non-significant after further adjustment]*<br>- Child abuse exposure without deployment-related trauma and child abuse exposure with deployment-related trauma *[Regulars]* were associated with increased odds of past-year suicidal ideation (AORs 2.0 [95% CI 1.3-3.0] and 2.7 [95% CI 1.8-4.2], *p*<.05) and suicide plans (AOR 2.9 [95% CI 1.4-6.0] and 4.6 [95% CI 2.3-9.2], *p*<.05)<br>- Experiencing child abuse exposure only, relative to deployment-related trauma only *[Regulars]* was associated with increased odds of past-year suicidal ideation but not suicide plans<br>- Experience of childhood abuse exposure and deployment-related trauma significantly increased the offs of past-year suicidal ideation and suicide plans compared with exposure to deployment-related trauma only<br>- Deployment-related trauma without history of child abuse exposure *[Regulars]* was not associated with past-year suicidal ideation (AOR 1.2 [95% CI 0.7-1.8]) or suicide plans (AOR 2.9 [95% CI 0.8-4.3]) | Fair |
| **Belik et al (2009)**<br>*Canada* | - Exposure to combat or peacekeeping operations was not significantly associated with an increased risk of suicide attempts<br>- Sexual and other interpersonal traumas are highly associated with suicide attempts in both men and women currently active in the Canadian military<br>- Increasing levels of traumatic exposure are associated with further increased likelihood of suicide attempts in this population | **Males:**<br>- Deployment-related traumatic events:<br>- *Combat (range AORs 1.25 [95% CI 0.70-2.21] and 1.15 [95% CI 0.62-2.12])<br>*Peacekeeping (range AORs 0.84 [95% CI 0.51-1.38] and 0.82 [95% CI 0.47-1.43])<br>*Purposely injured, tortured, or killed (range AORs 3.59 [95% CI 1.60-8.06] and 2.69 [95% CI 1.09-6.61])<br>*Witnessing atrocities (range AORs 1.97 [95% CI 1.17-3.31] and 1.68 [95% CI 0.94-3.01])<br>- Accident or other unexpected trauma:<br>*Auto accident (range AORs 1.07 [95% CI 0.64-1.80] and 0.91 [95% CI 0.50-1.68])<br>*Other accident (range AORs 1.78 [95% CI 1.06-2.99] and 1.59 [95% CI 0.90-2.81]) | Fair |

| | | |
|---|---|---|
| | | *Toxic chemical exposure (range AORs 1.87 [95% CI 1.14-3.06] and 1.86 [95% CI 1.09-3.18])<br>*Man-made disaster (range AOR 1.41 [95% CI 0.79-2.53] and 1.35 [95% CI 0.73-2.50])<br>*Natural disaster (range AORs 1.29 [95% CI 0.73-2.26] and 1.23 [95% CI 0.65-2.31])<br>*Unexpected death (range AORs 1.41 [95% CI 0.93-2.16] and 1.17 [95% CI 0.71-1.95])<br>*Child illness or injury (range AORs 1.15 [95% CI 0.42-3.18] and 0.64 [95% CI 0.18-2.22])<br>*Life threatening illness (range AORs 2.20 [95% CI 1.05-4.62] and 2.25 [95% CI 1.04-4.89])<br>*Witness death (range AORs 1.34 [95% CI 0.87-2.08] and 1.25 [95% CI 0.75-2.08])<br>*Caused death accidently (AOR 2.92 [95% CI 1.10-7.72] and 2.16 [95% CI 0.73-6.37])<br>- Sexual trauma<br>*Raped [range AORs 8.56 [95% CI 3.66-19.99] and 4.29 [95% CI 1.47-12.53])<br>*Sexual assault (range AORs 6.57 [95% CI 3.77-11.45] and 3.73 [95% CI 1.89-7.34])<br>- Other interpersonal traumas<br>*Child abuse (range AORs 6.51 [95% CI 3.70-11.49] and 4.43 [95% CI 2.35-8.36])<br>*Other person abused you (range AORs 3.24 [95% CI 2.04-5.15] and 2.31 [95% CI 1.34-3.96])<br>*Witness to domestic violence (range AORs 4.01 [95% CI 2.40-6.72] and 3.43 [95% CI 1.94-6.04])<br>*Stalked (range AORs 4.83 [95% CI 2.36-9.89] and 3.71 [95% CI 1.56-8.83])<br>*Mugged (range AORs 2.39 [95% CI 1.53-3.72] and 2.45 [95% CI 1.53-3.91])<br>*Kidnapped (range AORs 3.80 [95% CI 1.49-9.69] and 3.44 [95% CI 1.23-9.62])<br>- Civilian in war zone or refugee<br>*Civilian in war zone (range AORs 1.48 [95% CI 0.58-3.79] and 1.54 [95% CI 0.56-4.19])<br>*Civilian in religious terror (range AORs 2.46 [95% CI 1.12-5.38] and 2.38 [95% CI 1.00-5.72])<br>- Event happened to other (range AORs 3.08 [95% CI 1.87-5.07] and 2.07 [95% CI 1.17-3.64])<br>- Other trauma (range AORs 3.26 [95% CI 1.81-5.87] and 2.57 [95% CI 1.38-4.77])<br><br>**Women:**<br>- Deployment-related traumatic events:<br>- *Combat (range AORs 1.46 [95% CI 0.73-2.94] and 1.30 [95% CI 0.63-2.68])<br>*Peacekeeping (range AORs 1.26 [95% CI 0.73-2.18] and 1.15 [95% CI 0.62-2.16]) |

| | | | |
|---|---|---|---|
| | | *Witnessing atrocities (range AORs 1.43 [95% CI 0.57-3.58] and 0.96 [95% CI 0.37-2.50])<br>- Accident or other unexpected trauma:<br>*Auto accident (range AORs 1.50 [95% CI 0.88-2.57] and 1.28 [95% CI 0.66-2.48])<br>*Other accident (range AORs 2.43 [95% CI 1.26-4.70] and 1.91 [95% CI 0.97-3.77])<br>*Toxic chemical exposure (range AORs 1.21 [95% CI 0.62-2.36] and 1.25 [95% CI 0.62-2.50])<br>*Man-made disaster (range AOR 2.67 [95% CI 1.40-5.10] and 2.16 [95% CI 1.02-4.55])<br>*Natural disaster (range AORs 1.42 [95% CI 0.81-2.49] and 1.11 [95% CI 0.55-2.26])<br>*Unexpected death (range AORs 1.47 [95% CI 1.00-2.16] and 1.03 [95% CI 0.65-1.63])<br>*Child illness or injury (range AORs 1.42 [95% CI 0.48-4.18] and 1.07 [95% CI 0.31-3.65])<br>*Life threatening illness (range AORs 1.55 [95% CI 0.75-3.23] and 0.82 [95% CI 0.69-1.95])<br>*Witness death (range AORs 1.50 [95% CI 0.98-2.29] and 1.16 [95% CI 0.69-1.95])<br>- Sexual trauma<br>*Raped [range AORs 4.66 [95% CI 3.05-7.13] and 2.54 [95% CI 1.55-4.18])<br>*Sexual assault (range AORs 5.63 [95% CI 3.68-8.62] and 3.41 [95% CI 2.09-5.57])<br>- Other interpersonal traumas<br>*Child abuse (range AORs 3.98 [95% CI 2.29-6.93] and 2.34 [95% CI 1.15-4.75])<br>*Spouse abused you (range AORs 4.79 [95% CI2.91-7.89] and 3.71 [95% CI 1.97-6.99])<br>*Other person abused you (range AOR 4.91 [95% CI 2.26-10.73] and 3.08 [1.04-9.14])<br>*Witness to domestic violence (range AORs 2.38 [95% CI1.54-3.70] and 1.73 [95% CI 1.00-3.01])<br>*Stalked (range AORs 2.58 [95% CI 1.61-4.12] and 1.86 [95% CI 1.09-3.19])<br>*Mugged (range AORs 2.40 [95% CI 1.38-4.17] and 1.71 [95% CI 0.89-3.29])<br>*Kidnapped (range AORs 3.29 [95% CI 0.88-12.34] and 1.49 [95% CI 0.36-6.11])<br>- Civilian in war zone or refugee<br>*Civilian in war zone (range AORs 1.31 [95% CI 0.39-4.38] and 0.98 [95% CI 0.27-3.56])<br>*Civilian in religious terror (range AORs 0.27 [95% CI 0.08-0.90] and 0.22 [95% CI 0.07-0.71])<br>- Event happened to other (range AORs 2.47 [95% CI 1.60-3.82] and 2.21 [95% CI 1.34-3.62])<br>- Other trauma (range AORs 2.24 [95% CI 0.86-5.86] and 1.10 [95% CI 0.36-3.37]) | |
| **Bergman et al (2019)**<br>*UK* | - Highest risks of non-fatal self-harm in veterans with the shortest service, including | - Veterans *[vs non-veterans]* (HR 1.27 [95% CI 1.21-1.35], *p*<.001) | Good |

| | | | |
|---|---|---|---|
| | those who did not complete training, and in the oldest and youngest birth cohorts<br>- Veterans who served for the longest are at reduced risk of suicide<br>- Association between mental health disorder (especially PTSD) and self-harm in early service leaver veterans | **All Veterans:**<br>- Sex:<br>*Men (HR 1.38 [95% CI 1.30-1.47], *p*<.001)<br>*Women (HR 1.12 [95% CI 0.94-1.34], *p*=0.199)<br>- Length of Service:<br>*Untrained ESL (HR 1.69 [95% CI 1.50-1.91], *p*<.001)<br>*Trained ESL (HR 1.59 [95% CI 1.46-1.74], *p*<.001)<br>*4-6 years (HR 1.33 [95% CI 1.20-1.47], *p*<.001)<br>*7-9 years (HR 1.19 [95% CI 1.04-1.36], *p*=0.008)<br>*10-12 years (HR 0.97 [95% CI 0.82-1.14], *p*=0.690)<br>*13-16 years (HR 0.97 [95% CI 0.79-1.18], *p*=0.740)<br>*17-22 years (HR 0.89 [95% CI 0.70-1.14], *p*=0.362]<br>*≥23 years (HR 0.40 [95% CI 0.28-0.57]. *p*<.001)<br>- Age: Oldest (1945-1949) and youngest (1980-1985) cohorts<br>- Early service leavers (ESLs; ≤2.5 years service), especially those who did not complete initial training<br>- Older veterans with 4-6 years and 7-9 years service<br>Comorbid disorders *[veterans vs non-veterans]*:<br>*Peptic ulcer (OR 1.07 [95% CI 0.88-1.31], *p*=0.482)<br>*COPD (OR 0.97 [95% CI 0.81-1.13], *p*=0.726)<br>*Diabetes (OR 1.24 [95% CI 1.00-1.55], *p*0.049)<br>*Any cancer (OR 1.20 [95% CI 0.99-1.46], p=0.068)<br>*AMI (OR 1.22 [95% CI 0.95-1.57], *p*0.116)<br>*Alcoholic liver disease (OR 0.84 [95% CI 0.67-1.06], *p*0.140)<br>*Hepatitis C (OR 0.49 [95% CI 0.32-0.74], *p*<.001)<br>*Lung cancer (OR 2.13 [95% CI 1.28-3.53], *p*=0.002)<br>*Any mental health disorder (OR 1.08 [95% CI1.02-1.14], *p*=0.013)<br>*Any mental health disorder *excl PTSD* (OR 0.96 [95% CI 0.89-1.04], *p*=0.325)<br>*Mood disorder (OR 1.05 [95% CI 0.98-1.14], *p*=0.179)<br>*Anxiety (OR 1.18 [95% CI 1.06-1.30], *p*=0.002<br>*Stress/PTSD (OR 1.42 [95% CI 1.24-1.62], *p*<.001)<br>*Anxiety *excl stress/PTSD* (OR 0.91 [95% CI 0.76-1.10], *p*=0.351)<br>*Psychosis (OR 0.86 [95% CI 0.71-1.05], *p* 0.135) | |
| **Bergman et al (2022)**<br>*UK* | - No difference in risk of suicide overall between veterans and non-veterans | - No difference in risk of suicide overall between veterans and non-veteran (HR 1.01 [95% CI 0.90-1.13], *p*=0.859) | Good |

| | | | |
|---|---|---|---|
| | - Highest risk for both men and women in middle age, many years after leaving service | - Non-significant increase in overall risk for female veterans (HR 1.57 [95% CI 0.91-2.69], *p*=0.103)<br>- Older women veterans (>40 years) were at significantly increased risk (HR 2.65 [95% CI 1.41-4.99, *p*=0.003), but non-significant for older male veterans (HR 1.04 [95% CI 0.90-1.21, p=0.571)<br>- ESLs at statistically significant increased risk of suicide compared to non-veterans (HR 1.25 [95% CI 1.05-1.48], *p*=0.010) *[non-significant after adjustment (HR 1.14 [95% CI 0.96-1.35], p0.138)]*<br>- Years of service:<br>*Veterans with 17-22 years' service (HR 1.29 [95% CI 0.86-1.91], *p*=0.218)<br>*Veterans with >22 years' service (HR 0.57 [95% CI 0.30-1.06], *p*=0.076)<br>- Antecedent conditions *[veterans vs non-veterans]*:<br>*Mood disorder (OR 1.10 [95% CI 0.87-1.40], *p*=0.423)<br>*PTSD (OR 1.26 [95% CI 0.81-1.97], *p*=0.306)<br>*Type 2 Diabetes (OR 1.01 [95% CI 0.55-1.83], *p*=0.984)<br>*Cancer (OR 1.15 [95% CI 0.61-2.04], *p*=0.724)<br>*Cardiovascular disease (OR 1.57 [95% CI 0.82-3.05], *p*=0.173)<br>*Alcoholic liver disease (OR 0.79 [95% CI 0.30-2.05], *p*=0.625) | |
| **Boulos & Fikretoglu (2018)**<br>*Canada* | - Past-year mental health problems differences were identified between components<br>- Deployment-related experiences were highly associated with mental health problems, but these only partially accounted for mental health problem differences between components<br>- Regular Force higher prevalence of suicidal ideation compared to Reserve Force (in this sample) | - Reserves vs Regulars (OR 1.27 [95% CI 1.01-1.60]) [*non-significant after adjustment (range AORs 1.24 [95% CI 0.94-1.61] to 1.18 [95% CI 0.91-1.54])*<br>- Other non-Afghanistan deployment (AOR 0.85 [95% CI 0.61-1.19])<br>- Time away on deployment in past 3 years *(vs none)*<br>*<6 months (AOR 0.60 [95% CI 0.36-0.99])<br>*7-12 months (AOR 0.51 [95% CI 0.29-0.88])<br>*13-24 months (AOR 0.54 [95% CI 0.32-0.91])<br>*25-36 months (AOR 0.54 [95% CI 0.26-1.13])<br>- Interval from last Afghanistan deployment to survey interview date *(vs ≥7 years)*<br>*<4 years (AOR 1.78 [95% CI 1.01-3.14])<br>*4 years (AOR 1.24 [95% CI 0.68-2.23])<br>*5 years (AOR 1.12 [95% CI 0.65-1.93])<br>*6 years (AOR 1.09 [95% CI 0.59-2.01])<br>- ≥2 total number of Afghanistan deployments *(vs 1)* (AOR 1.43 [95% CI 0.83-2.47])<br>- Afghanistan deployment location *(vs other Afghanistan related)*<br>*Kabul (AOR 0.57 [95% CI 0.26-1.26]) | Fair |

| | | | |
|---|---|---|---|
| | | *Kandahar (AOR 0.64 [95% CI 0.32-1.28])<br>*Multiple (AOR 0.88 [95% CI 0.43-1.83])<br>- Cumulative duration of Afghanistan deployments *(vs ≤120 days)*<br>*121-240 days (AOR 0.74 [95% CI 0.46-1.19])<br>*241-360 days (AOR 0.70 [95% CI 0.39-1.29])<br>*≥361 days (AOR 0.48 [95% CI 0.23-0.98])<br>- Deployment-related experiences *(vs no)*<br>*ever known someone who was seriously injured or killed (AOR 1.60 [95% CI 1.03-2.49])<br>*ever found yourself in a threatening situation where you were unable to respond because of the rules of engagement (AOR 1.81 [95% CI 1.28-2.56])<br>*ever been injured (AOR 1.40 [95% CI 1.03-1.90])<br>*ever seen ill or injured women or children who you were unable to help (AOR 0.85 [95% CI 0.62-1.18])<br>*ever received incoming artillery, rocket or mortar fire (AOR 0.86 [95% CI 0.57-1.32])<br>*ever felt responsible for the death of a Canadian or ally personnel (AOR 2.45 [95% CI 1.61-3.73])<br>*ever had a close call, for example shot or hit but protective gear saved you (AOR 0.82 [95% CI 0.58-1.15])<br>*ever had difficulty distinguishing between combatants and non-combatants (AOR 1.01 [95% CI 0.73-1.41])<br>- Deployment-related lifetime potentially traumatic event *(vs no)*<br>*Combat experience (AOR 1.08 [95% CI 0.74-1.59])<br>*Relief worker in a war zone (AOR 1.23 [95% CI 0.90-1.85])<br>*Saw atrocities (AOR 0.92 [95% CI 0.65-1.32]) | |
| **Harden & Murphy (2018)**<br>*UK* | - Unemployment, being an early service leaver, taking less than 5 years to seek help and experiencing pre-service childhood adversity were associated with suicidal ideation (after adjusting for relevant variables)<br>- No association between health outcomes and suicidal ideation | - Age *(vs <35 years old)*<br>*35-44 years (AOR 0.30 [95% CI 0.05-0.78])<br>*45-54 years (AOR 0.20 [95% CI 0.04-0.97])<br>*55+ years (AOR 0.29 [95% CI 0.06-1.39])<br>- Female sex *(vs male)* (AOR 6.68 [95% CI 0.92-48.28])<br>- Not in a relationship *(vs in a relationship)* (AOR 1.77 [95% CI 0.59-5.34])<br>- Not working *(vs working)* (AOR 8.01 [95% CI 1.79-35.80])<br>- Financial difficulties *(vs no)* (AOR 0.38 [95% CI 0.06-1.87])<br>- Greater than 5 years to seek help *(vs less than 5 years)* (AOR 0.10 [95% CI 0.06-0.87]) | Fair |

| | | | |
|---|---|---|---|
| | | - Early service leave *(vs no)* (AOR 8.46 [95% CI 2.21-32.35])<br>- Combat role *(vs no)* (AOR 1.21 [95% CI 0.38-3.82])<br>- Non-voluntary discharge from service *(vs voluntary)* (AOR 2.16 [95% CI 0.63-7.38])<br>- High childhood adversity *(vs low)* (AOR 6.92 [95% CI 2.10-22.82])<br>- Health outcomes:<br>*PTSD (AOR 1.00 [95% CI 0.95-1.04])<br>*CMD (AOR 1.03 [95% CI 0.91-1.18])<br>*Anger (AOR 0.99 [95% CI 0.90-1.11])<br>*Alcohol (AOR 1.01 [95% CI 0.96-1.06])<br>*Functioning (AOR 1.02 [95% CI 0.96-1.09]) | |
| **Hines et al (2013)**<br>*UK* | - Overall prevalence of self-harm in the UK military was 2.3%, and was significantly lower in Reserves (0.7%) than in Regulars (2.5%)<br>- Factors associated with self-harm in the military reflected those found in the general population<br>- Self-harm was not associated with deployment, but was linked to available social support in childhood and adulthood | - Age (AOR 0.95 [95% CI 0.92-0.98])<br>- Female sex *(vs male)* (AOR 1.79 [95% CI 1.17-2.74])<br>- Reserve status *(vs Regular)* (AOR 0.30 [95% CI 0.17-0.56])<br>- Marital status *(vs married)*<br>*Single (AOR 0.95 [95% CI 0.60-1.50])<br>*Separated/divorced (AOR 2.15 [95% CI 1.23-3.75])<br>- Deployed *(vs non-deployed)* (OR 0.78 [95% CI 0.56-1.09] *AORs not presented*)<br>- Discharged *(vs serving)* (AOR 2.34 [95% CI 1.61-3.40])<br>- Social support: Number of close family/friends *(vs 3-5)*<br>*None (AOR 2.79 [95% CI 1.40-5.55])<br>*1-2 (AOR 1.48 [95% CI 0.95-2.29])<br>*6-10 (AOR 0.71 [95% CI 0.43-1.16])<br>*11-15 (AOR 0.79 [95% CI 0.30-2.07])<br>*15+ (AOR 1.03 [95% CI 0.49-2.16])<br>- Social support: Family relationship adversity *(vs 0)*<br>*1 item (AOR 1.21 [95% CI 0.71-2.08])<br>*2+ items (AOR 2.25 [95% CI 1.50-3.38])<br>- Childhood adversity: Childhood anti-social behaviour *(vs no)* (OR 2.07 [95% CI 1.44-2.99] *AORs not presented*)<br>- Childhood adversity: Time spent in local authority/social services care *(vs no)* (AOR 2.19 [95% CI 1.07-4.50]) | Fair |
| **Jones et al (2019)**<br>*UK* | - Rate of lifetime self-harm increased significantly among UK AF serving and ex-serving personnel from 2004 to 2016 | **Lifetime Self-Harm:**<br>- Veteran *(vs serving)*<br>*Phase 1 (AOR 1.76 [95% CI 1.17-2.62])<br>*Phase 2 (AOR 2.36 [95% CI 1.62-3.45]) | Good |

| | | |
|---|---|---|
| | - A greater number of ex-serving personnel experience lifetime self-harm on each occasion the outcome was measured<br>- Main risk factors for self-harm and suicide attempts (interview study) were symptoms of mental disorder and suicidal ideation<br>- Subjectively higher levels of social support were protective for both self-harm and suicide attempts<br>- Stigmatising beliefs and more negative social attitudes about mental illness and perceived barriers to care were associated with greater frequency of self-harm<br>- Suicide prevention should focus on ameliorating mental disorder by encouraging engagement with health care, reducing negative views of mental illness, and fostering social support | *Phase 3 (AOR 1.95 [95% CI 1.46-2.62])<br>*Telephone Interview Study (AOR 1.26 [95% CI 0.91-1.73])<br><br>**Lifetime Self-Harm (Telephone Interview Study):**<br>- Age *(vs <30 years)*<br>*30-34 years (OR 1.06 [95% CI 0.64-1.75]; AOR1 1.17 [95% CI 0.69-1.98]; AOR2 1.09 [95% CI 0.65-1.81]; AOR3 1.17 [95% CI 0.68-2.01])<br>*35-39 years (OR 0.59 [95% CI 0.35-1.00]; AOR 0.71 [95% CI 0.40-1.26]; AOR2 0.59 [95% CI 0.34-1.00]; AOR3 0.68 [95% CI 0.38-1.23])<br>*40-49 years (OR 0.50 [95% CI 0.31-0.80]; AOR1 0.47 [95% CI 0.29-0.77]; AOR2 0.52 [95% CI 0.32-0.84]; AOR3 0.62 [95% CI 0.36-1.09])<br>*>49 years (OR 0.15 [95% CI 0.14-0.30]; AOR1 0.14 [95% CI 0.06-0.33]; AOR2 0.14 [95% CI 0.06-0.33]; AOR3 0.18 [95% CI 0.73-0.47])<br>- Rank *(vs junior rank)*<br>*Junior NCO to Warrant Officer (OR 0.53 [95% CI 0.35-0.81]; AOR1 0.77 [95% CI 0.49-1.22]; AOR2 0.57 [95% CI 0.37-0.87]; AOR3 0.85 [95% CI 0.53-1.35])<br>*Commissioned Officer (OR 0.32 [95% CI 0.19-0.55]; AOR1 0.52 [95% CI 0.28-0.94]; AOR2 0.37 [95% CI 0.21-0.63]; AOR3 0.60 [95% CI 0.33-1.11])<br>- Female sex *(vs male)* (OR 2.27 [95% CI 1.55-3.34]; AOR1 2.01 [95% CI 1.40-3.14]; AOR2 2.25 [95% CI 1.52-3.33]; AOR3 2.03 [95% CI 1.35-3.05])<br>- Service branch *(vs Army)*<br>*RAF (OR 0.90 [95% CI 0.59-1.38]; AOR1 0.82 [95% CI 0.52-1.27]; AOR2 0.94 [95% CI 0.62-1.44]; AOR3 0.88 [95% CI 0.56-1.37]<br>*Royal Navy/Royal Marines (OR 1.07 [95% CI 0.66-1.74]; AOR1 1.10 [95% CI 0.68-1.81]; AOR2 1.06 [95% CI 0.65-1.73]; AOR3 1.10 [95% CI 0.67-1.81])<br>- Reserve *(vs Regular)* (OR 0.92 [95% CI 0.60-1.42]; AOR1 1.17 [95% CI 0.80-1.73]; AOR2 0.82 [95% CI 0.58-.15]; 3 1.14 [95% CI 0.78-1.69])<br>- Veteran *(vs serving)* (*OR 0.85* [95% CI 0.61-1.20]; AOR1 1.17 [95% CI 0.80-1.73]; AOR2 0.82 [95% CI 0.58-1.15]; AOR3 1.14 [95% CI 0.78-1.69])<br>- Mental health *(vs no case)*<br>*PHQ-9 (OR 3.29 [95% CI 2.07-5.23]; AOR1 3.11 [95% CI 1.91-5.06])<br>*GAD-7 (OR 2.37 [95% CI 1.64-3.43]; AOR1 2.11 [95% CI 1.51-3.24])<br>*PCL-5 (OR 3.31 [95% CI 2.12-5.15]; AOR1 3.37 [95% CI 2.13-5.33])<br>*Lifetime suicidal ideation (OR 1.05 [95% CI 0.75-1.46]; AOR1 1.12 [95% CI 0.79-1.57]; AOR2 0.99 [95% CI 0.71-1.39]; AOR3 1.06 [95% CI 0.75-1.49]) |

| | | |
|---|---|---|
| | | - High score perceived social support *(vs low to moderate score)* (OR 0.56 [95% CI 0.40-0.78]; AOR1 0.51 [95% CI 0.36-0.72]; AOR2 0.64 [95% CI 0.45-0.90]; AOR3 0.58 [95% CI 0.40-0.82])<br>- Mental health stigmatization *(vs lower score)*<br>*Moderate score (OR 1.77 [95% CI 1.14-2.75]; AOR1 0.68 [95% CI 1.07-2.63]; AOR2 1.69 [95% CI 1.08-2.64]; AOR3 1.60 [95% CI 1.02-2.52])<br>*Higher score (OR 2.25 [95% CI 1.47-3.45]; AOR1 2.06 [95% CI 1.34-3.22]; AOR2 1.97 [95% CI 1.27-3.05]; AOR3 1.83 [95% CI 1.16-2.87])<br>- Perceived practical barriers to care *(vs lower score)*<br>*Moderate score (OR 1.14 [95% CI 0.73-1.77]; AOR1 1.22 [95% CI 0.78-1.92]; AOR2 1.07 [95% CI 0.69-1.67]; AOR3 1.17 [95% CI 0.74-1.84])<br>*Higher score (OR 1.83 [95% CI 1.26-2.66]; AOR1 1.29 [95% CI 1.90-2.80]; AOR2 1.61 [95% CI 1.10-2.36]; AOR3 1.70 [95% CI 1.15-2.53])<br>- Negative attitudes to mental illness *(vs lower score)*<br>*Moderate score (OR 1.67 [95% CI 1.07-2.61]; AOR1 1.73 [95% CI 1.10-2.73]; AOR2 1.70 [95% CI 1.08-2.67]; AOR3 1.76 [95% CI 1.11-2.79])<br>*Higher score (OR 1.86 [95% CI 1.28-2.70]; AOR1 1.53 [95% CI 1.03-2.26]; AOR2 1.67 [95% CI 1.14-2.42]; AOR3 1.37 [95% CI 0.91-2.04])<br>- Help-seeking *(vs none)*<br>*Informal (OR 1.74 [95% CI 0.65-3.98]; AOR1 1.50 [95% CI 0.56-3.98]; AOR2 1.86 [95% CI 0.70-4.95]; AOR3 1.59 [95% CI 0.61-4.19]<br>*Formal nonmedical (OR 2.52 [95% CI 0.92-6.90]; AOR1 2.01 [95% CI 0.73-5.59]; AOR2 2.75 [95% CI 1.00-7.53]; AOR3 2.21 [95% CI 0.81-6.08])<br>- *Formal medical (OR 2.94 [95% CI 1.16-7.44]; AOR1 2.48 [95% CI 1.00-6.23]; AOR2 2.80 [95% CI 1.11-7.07]; AOR3 2.41 [95% CI 0.97-6.96])<br><br>**Lifetime Suicide Attempts (Telephone Interview Study):**<br>- Age *(vs <30 years)*<br>*30-34 years (OR 0.81 [95% CI 0.43-1.54]; AOR1 0.97 [95% CI 0.49-1.92]; AOR2 0.83 [95% CI 0.43-1.59]; AOR3 0.95 [95% CI 0.47-1.91])<br>*35-39 years (OR 0.70 [95% CI 0.37-1.31]; AOR1 0.92 [95% CI 0.45-1.86]; AOR2 0.70 [95% CI 0.37-1.31]; AOR3 0.87 [95% CI 0.43-1.77])<br>*40-49 years (OR 0.90 [95% CI 0.53-1.53]; AOR1 1.16 [95% CI 0.59-2.28]; AOR2 0.95 [95% CI 0.56-1.63]; AOR3 1.13 [95% CI 0.57-2.27])<br>*>49 years (OR 0.98 [95% CI 0.54-1.79]; AOR1 1.31 [95% CI 0.60-2.84]; AOR2 0.98 [95% CI 0.53-1.82]; AOR3 1.21 [95% CI 0.54-2.69]) |

| | | |
|---|---|---|
| | | - Rank *(vs junior rank)*<br>*NCO to Warrant Officer (OR 0.71 [95% CI 0.45-1.12]; AOR1 0.66 [95% CI 0.39-1.10]; AOR2 0.79 [95% CI 0.49-1.26]; AOR3 0.75 [95% CI 0.45-1.27])<br>*Commissioned Officer (OR 0.34 [95% CI 0.18-0.62]; AOR1 0.32 [95% CI 0.16-0.64]; AOR2 0.41 [95% CI 0.22-0.76]; AOR3 0.40 [95% CI 0.19-0.82])<br>- Female sex *(vs male)* (OR 1.00 [95% CI 0.61-1.63; AOR1 1.06 [95% CI 0.64-1.77]; AOR2 0.96 [95% CI 0.59-1.57]; AOR3 1.02 [95% CI 0.63-1.69])<br>- Service Branch *(vs Army)*<br>*RAF (OR 0.46 [95% CI 0.27-0.79]; AOR1 0.42 [95% CI 0.24-0.72]; AOR2 0.49 [95% CI 0.28-0.84]; AOR3 0.46 [95% CI 0.27-0.80])<br>*Royal Navy/Royal; Marines (OR 0.98 [95% CI 0.60-1.62]; AOR1 1.07 [95% CI 0.65-1.77]; AOR2 1.04 [95% CI 0.70-1.55]; AOR3 1.04 [95% CI 0.62-1.75])<br>- Reserve status *(vs Regular)* (OR 1.64 [95% CI 1.09-2.46]; AOR1 1.42 [95% CI 0.90-2.22]; AOR2 1.78 [95% CI 1.17-2.70]; AOR3 1.58 [95% CI 1.00-2.49])<br>- Veteran *(vs serving)* (OR 1.39 [95% CI 0.98-1.98]; AOR1 1.39 [95% CI 0.93-2.08]; AOR2 1.32 [95% CI 0.92-1.89]; AOR3 1.14 [95% CI 0.75-1.71])<br>- Mental health *(vs no case)*<br>*PHQ-9 (OR 5.83 [95% CI 3.73-9.13]; AOR1 5.53 [95% CI 3.51-8.71])<br>*GAD-7 (OR 2.76 [95% CI 1.89-4.05; AOR1 2.55 [95% CI 1.74-3.75])<br>*PCL-5 (OR 4.61 [95% CI 2.97-7.17]; AOR1 4.32 [95% CI 2.77-6.75])<br>*AUDIT-C (OR 0.97 [95% CI 0.68-1.38]; AOR1 1.00 [95% CI 0.69-1.43]; AOR2 0.89 [95% CI 0.62-1.28]; AOR3 0.91 [95% CI 0.63-1.32])<br>*Lifetime suicidal ideation (OR 12.22 [95% CI 6.82-21.93]; AOR1 11.73 [95% CI 6.54-21.04]; AOR2 10.82 [95% CI 5.98-19.57]; AOR3 10.45 [95% CI 5.78-18.88])<br>- High score perceived social support *(vs low to moderate score)* (OR 0.53 [95% CI 0.37-0.76]; AOR1 0.55 [95% CI 0.39-0.79]; AOR2 0.65 [95% CI 0.46-0.94]; AOR3 0.67 [95% CI 0.47-0.97])<br>- Mental health stigmatization *(vs lower score)*<br>*Moderate score (OR 1.33 [95% CI 0.85-2.06]; AOR1 1.33 [95% CI 0.85-2.07]; AOR2 1.22 [95% CI 0.78-1.92]; AOR3 1.33 [95% CI 0.85-2.07])<br>*Higher score (OR 1.45 [95% CI 0.94-2.25]; AOR1 1.53 [95% CI 0.98-2.39]; AOR2 1.15 [95% CI 0.74-1.80]; AOR3 1.53 [95% CI 0.98-2.39])<br>- Perceived practical barriers to care *(vs lower score)*<br>*Moderate score (OR 1.12 [95% CI 0.67-1.88]; AOR1 1.10 [95% CI 0.66-1.85]; AOR2 1.14 [95% CI 0.67-1.93]; AOR3 1.11 [95% CI 0.66-1.87]) |

| | | | |
|---|---|---|---|
| | | *Higher score (OR 1.87 [95% CI 1.27-2.75]; AOR1 1.60 [95% CI 1.07-2.38]; AOR2 1.60 [95% CI 1.07-2.38]; AOR3 1.38 [95% CI 0.92-2.09])<br>- Negative attitudes to mental illness *(vs lower score)*<br>*Moderate score (OR 1.69 [95% CI 1.09-2.63]; AOR1 1.79 [95% CI 1.15-2.79]; AOR2 1.56 [95% CI 0.99-2.44]; AOR3 1.64 [95% CI 1.04-2.58])<br>*Higher score (OR 1.84 [95% CI 1.22-2.79]; AOR1 1.84 [95% CI 1.21-2.81]; AOR2 1.51 [95% CI 0.99-2.31]; AOR3 1.52 [95% CI 0.98-2.34])<br>- Help-seeking *(vs none)*<br>*Informal (OR 1.19 [95% CI 0.47-3.01]; AOR1 1.20 [95% CI 0.47-3.05]; AOR2 1.29 [95% CI 0.51-3.27]; AOR3 1.20 [95% CI 0.47-3.05]<br>*Formal nonmedical (OR 0.52 [95% CI 0.16-1.65]; AOR1 0.57 [95% CI 0.18-1.86]; AOR2 0.58 [95% CI 0.18-1.84]; AOR3 0.57 [95% CI 0.18-1.86])<br>*Formal medical (OR 2.55 [95% CI 1.09-5.97]; AOR1 2.67 [95% CI 1.13-6.30]; AOR2 2.39 [95% CI 1.02-5.60]; AOR3 2.67 [95% CI 1.13-6.30]) | |
| **Jones et al (2021)**<br>*UK* | - No significant associations between those who had enlisted as JEs compared to those who joined as SEs in respect of lifetime self-harm<br>- Among the smaller sub-sample of more recent joiners, lifetime self-harm was associated with junior entry | - Junior entrants enlisted between April 2003 and March 2013 (AOR 2.13 [95% CI 1.15-2.93])<br>- Junior entry <17.5 years (AOR 1.21 [95% CI 0.84-1.75]) | Fair |
| **Kapur et al (2009)**<br>*UK* | - Young men who leave UK Armed Forces were at increased risk of suicide<br>- Transition to civilian life is difficult for some individuals<br>- Highest risk of suicide after discharge may have been those with the most adverse experiences whilst serving in the military<br>- High suicide risk may be a consequence of pre-enlistment vulnerability | - Age *(vs 45-59 years)*<br>*16-19 years (HR 2.3 [95% CI 1.1-4.5])<br>*20-24 years (HR 2.0 [95% CI 1.0-3.9])<br>*25-29 years (HR 1.5 [95% CI 0.7-3.1])<br>*30-34 years (HR 1.1 [95% CI 0.5-2.3])<br>*35-39 years (HR 1.1 [95% CI 0.5-2.4])<br>*40-44 years (HR 0.8 [95% CI0.3-2.0])<br>- Female *(vs male)* (HR 0.4 [95% CI 0.2-0.7])<br>- Service branch *(vs British Army)*<br>*Naval Service (HR0.5 [95% CI 0.4-0.7]<br>*RAF (HR 0.4 [95% CI 0.3-0.6])<br>- Officer Rank *(vs Other rank)* (HR 0.5 [95% CI 0.3-0.8])<br>- Marital status *(vs not married)*<br>*Unknown (HR 0.4 [95% CI 0.2-0.7])<br>*Married (HR 0.4 [95% CI 0.3-0.6]) | Good |

| | | | |
|---|---|---|---|
| | | - Untrained *(vs trained)* (HR 1.5 [95% CI 1.1-1.9])<br>- Medical discharge *(vs administrative discharge)* (HR 1.2 [95% CI 0.7-1.9])<br>- Length of service *(vs 15+ years)*<br>*<1 year (HR 2.2 [95% CI 1.4-3.3])<br>*1-<2 years (HR 3.4 [95% CI 2.0-5.8])<br>*2-<3 years (HR 3.2 [95% CI 1.6-6.3])<br>*3-<4 years (HR 3.5 [95% CI 1.9-6.2])<br>*4-<5 years (HR 0.9 [95% CI 0.4-2.3])<br>*5-<10 years (HR 1.5 [95% CI 0.9-2.4])<br>*10-<15 years (HR 0.9 [95% CI 0.5-1.7]) | |
| **Kerr et al (2018)**<br>*Australia* | - Age, employment status and conflicts served significantly differed between veterans with PTSD who had attempted suicide, compared to veterans with PTSD who had not<br>- PTSD symptom severity (measured by the CAPS-IV) and not working/total and permanently incapacitated (TPI) status were significant predictors of suicide attempt history<br>- Those with more severe psychopathology, who served in contemporary conflicts, and who were unemployed or on TPI were significantly more likely to have attempted suicide in the past<br>- Results highlight the importance of early identification of PTSD, therapeutic and social engagement, and the diligent application of effective treatment modalities<br>- Highlights need for tangible employment options, or meaningful and goal-directed activities in those veterans deemed unable to work, to facilitate socialisation and provide veterans with a sense of continued purpose and worth, potentially reducing suicide risk | - Age (OR 0.98 [95% CI 0.94-1.03])<br>- Employment *(vs full-time)*<br>*Part-time (OR 1.82 [95% CI 0.36-9.14])<br>*Retired (OR 2.36 [95% CI 0.75-7.40])<br>*Not working/totally and permanently incapacitated (OR 3.26 [95% CI 1.33-7.97])<br>- Conflicts served *(vs no deployment)*<br>*Pre-East Timor (OR 0.81 [95% CI 0.22-2.91])<br>*From East Timor onwards (OR 0.61 [95% CI 0.17-2.15])<br>*Both periods (OR 2.25 [95% CI 0.39-13.1])<br>- Mental health<br>*CAPS-IV PTSD scale (OR 1.02 [95% CI1.00-1.05])<br>*DAR (OR 0.99 [95% CI 0.96-1.03])<br>*HADS Anxiety score (OR 1.05 [95% CI 0.91-1.22])<br>*HADS Depression score (OR 1.04 [95% CI 0.92-1.18]) | Fair |
| **Ketcheson et al (2018)**<br>*Canada* | - Veterans with low or medium social support had more frequent suicidal ideation than CAF | - Perceived social support *(vs high)*<br>*Low (Regression coefficient 0.73, SE 0.10, *p*<.001) | Good |

| | | | |
|---|---|---|---|
| | members reporting that same level of social support<br>- Compared to CAF members with high social support, those with low social support had more frequent suicidal ideation<br>- CAF members with medium social support had lower suicidal ideation frequency than CAF members with high social support | *Medium (Regression coefficient 0.31, SE 0.09, *p<.01*)<br>- Female sex *(vs male)* (Regression coefficient 0.05, SE 0.12)<br>- Age (Regression coefficient 0.01, SE 0.00, *p*<.01)<br>- Service duration (Regression coefficient -0.01, SE 0.00, *p*<.01)<br>- Currently service *(vs veteran)* (Regression coefficient 0.18, SE 0.17)<br>- Currently serving AND low perceived social support (Regression coefficient -0.50, SE -0.23, *p<.05)*<br>- Currently serving AND medium perceived social support (regression coefficient -0.50, SE -0.23, *p<.05)* | |
| **Nelson et al (2011)**<br>*Canada* | - Social support and number of lifetime traumatic events experienced (including combat exposure) were significant predictors of past year suicidal ideation<br>- Depression mediated the relationship between number of traumatic events and suicidal ideation<br>- Past year diagnosis of PTSD or major depressive disorder is a significant predictor of suicidal ideation<br>- Dose response reaction relating to number of traumatic events experienced and likelihood of adverse mental health outcomes in military personnel | - Higher levels of perceived social support (β = 0.11, *p*<.05)<br>- Higher number of lifetime traumatic experiences (including exposure to combat)<br>- Health: Past year diagnosis of PTSD or major depressive disorder (MDD)<br>- Self-perceived religiosity (β = 0.22, *p*>.05)<br>- Past-year MDD mediates relationship between number of lifetime traumatic events and past-year SI (β = 0.04, *B* = 0.07 [SE = 0.02], *p*<.001)<br>- Past-year PTSD did not mediate the relationship between either perceived social support and pat-year suicidal ideation [β = 0.01, *B* = 0.02 (SE = 0.06), *p*>.05] or between number of lifetime traumatic events experienced and past-year suicidal ideation [β = -0.01, *B* = 0.02 (SE = 0.06), *p*≥.05] | Fair |
| **O'Toole et al (2015)**<br>*Australia* | - Both Australian Vietnam veterans and their partners had significantly higher rates of suicidal ideation, suicide planning, and attempting suicide than the age- and sex-matched general<br>- In veterans, PTSD, alcohol disorders, phobia, and depression predicted ideation, and alcohol disorders, phobia and depression predicted planning but PTSD, phobia, agoraphobia and depression predicted attempts<br>- Robust associations between depression and suicidality in both sexes, and that PTSD was | **Suicidal ideation:**<br>- PTSD (OR 3.95 [95% CI 2.51-6.21], *p*<.001)<br>- Alcohol dependence (OR 5.38 [95% CI 3.28-8.82], *p*<.001)<br>- Alcohol abuse (OR 0.71 [95% CI 0.44-1.15])<br>- Cannabis abuse (OR 3.81 [95% CI 1.35-10.75], *p*<.01)<br>- Depression single mild (OR 1.43 [95% CI 0.60-3.39])<br>- Depression single moderate (OR 5.03 [95% CI 2.01-12.13], *p*<.001)<br>- Depression single severe (OR 15.23 [95% CI 5.56-41.72], *p*<.001)<br>- Depression recurrent mild (OR 2.58 [95% CI 1.32-5.03], *p*<.01)<br>- Depression recurrent moderate (OR 4.07 [95% CI 1.82-9.10], *p*<.001)<br>- Depression recurrent severe (OR 14.03 [95% CI 5.20-39.37], *p*<.001)<br>- Depression (moderate + severe) lifetime (OR 15.66 [95% CI 9.18-26.74], *p* <.001) | Fair |

| | | |
|---|---|---|
| | associated with increasing severity of suicidality in veterans and in partners<br>- A higher education reduced the risk in veterans but not in partners | - Dysthymia (OR 1.42 [95% CI 0.43-4.69])<br>- Generalised anxiety disorder (OR 2.84 [95% CI 1.59-5.11], *p*<.001)<br>- Obsessive compulsive disorder (OR 4.94 [95% CI 1.37-17.85], *p*<.05)<br>- Agoraphobia without panic (OR 2.39 [95% CI 0.94-6.11])<br>- Panic without agoraphobia (OR 3.94 [95% CI 1.18-13.18], *p*<.05)<br>- Panic with agoraphobia (OR 9.69 [95% CI 1.00-94.10])<br>- Social phobia (OR 4.07 [95% CI 1.82-9.10], *p*<.001)<br>- Situational phobia (OR 2.97 [95% CI 1.36-6.45], *p*<.01)<br><br>**Suicide plan:**<br>- PTSD (OR 3.48 [95% CI 2.09-5.81], *p*<.001)<br>- Alcohol dependence (OR 4.50 [95% CI 2.64-7.68], *p*<.001)<br>- Alcohol abuse (OR 0.78 [95% CI 0.45-1.35])<br>- Cannabis abuse (OR 3.52 [95% CI 1.21-10.20], *p*<.05)<br>- Depression single mild (OR 1.53 [95% CI 0.60-3.96])<br>- Depression single moderate (OR 3.06 [95% CI 1.24-7.58], *p*<.05)<br>- Depression single severe (OR 16.51 [95% CI 6.60-41.29], *p*<.001)<br>- Depression recurrent mild (OR 1.27 [95% CI 0.56-2.88])<br>- Depression recurrent moderate (OR 2.86 [95% CI 1.22-6.68], *p*<.05)<br>- Depression recurrent severe (OR 6.90 [95% CI 2.96-16.10], *p*<.001)<br>- Depression (moderate + severe) lifetime (OR 11.76 [95% CI 6.72-20.56], *p* <.001)<br>- Dysthymia (OR 0.90 [95% CI 0.20-4.15])<br>- Generalised anxiety disorder (OR 2.58 [95% CI 1.37-4.88], *p*<.01)<br>- Obsessive compulsive disorder (OR 3.44 [95% CI 0.95-12.52])<br>- Agoraphobia without panic (OR 1.35 [95% CI 0.43-4.17])<br>- Panic without agoraphobia (OR 4.37 [95% CI 1.30-14.71], *p*<.05)<br>- Panic with agoraphobia (OR 5.08 [95% CI 0.71-36.66])<br>- Social phobia (OR 2.36 [95% CI 0.98-5.64])<br>- Situational phobia (OR 4.25 [95% CI 1.92-9.41], *p*<.001)<br><br>**Suicide attempt:**<br>- PTSD (OR 4.39 [95% CI 2.07-9.31], *p*<.001)<br>- Alcohol dependence (OR 7.32 [95% CI 3.49-15.41], *p*<.001)<br>- Alcohol abuse (OR 0.54 [95% CI 0.23-1.28])<br>- Cannabis abuse (OR 0.90 [95% CI 0.14-7.03]) |

| | | | |
|---|---|---|---|
| | | - Depression single mild (OR 1.05 [95% CI 0.24-4.66])<br>- Depression single moderate (OR 2.08 [95% CI 0.58-7.44])<br>- Depression single severe (OR 2.59 [95% CI 0.83-8.04])<br>- Depression recurrent mild (OR 3.80 [95% CI 1.60-9.18], *p*<.01)<br>- Depression recurrent moderate (OR 1.70 [95% CI 0.48-6.00])<br>- Depression recurrent severe (OR 6.30 [95% CI 2.40-16.55], *p*<.001)<br>- Depression (moderate + severe) lifetime (OR 4.49 [95% CI 2.16-9.19], *p* <.001)<br>- Dysthymia (OR 2.37 [95% CI 0.50-11.17])<br>- Generalised anxiety disorder (OR 1.66 [95% CI 0.65-4.22])<br>- Obsessive compulsive disorder (OR 1.41 [95% CI 0.17-11.48])<br>- Agoraphobia without panic (OR 3.68 [95% CI 1.15-11.80], *p*<.05)<br>- Panic with agoraphobia (OR 13.32 [95% CI 1.81-97.82], *p*<.05)<br>- Social phobia (OR 4.39 [95% CI 1.63-11.84], *p*<.01)<br>- Situational phobia (OR 3.97 [95% CI 1.49-10.61], *p*<.01) | |
| **Pinder et al (2012)**<br>*UK* | - Lifetime prevalence of 5.6% for attempted suicide and self-harm is higher than previous research has suggested<br>- Early service leavers, those who experienced childhood adversity, those with other psychological morbidity, and ex-service personnel are more likely to report self-harm behaviours | - Age (AOR 0.94 [95% CI 0.87-1.00], *p*=0.042)<br>- Female sex *(vs male)* (OR 0.90 [95% CI 0.42-1.93] *AORs not presented*)<br>- Marital status *(vs married/in long-term relationship)*<br>*Single/not in a relationship (OR 1.93 [95% CI 0.76-4.91] *AORs not presented*)<br>*Divorced/separated/widowed (OR 1.95 [95% CI 0.70-5.46 *AORs not presented*)<br>- Educational status *(vs O-levels)*<br>*No qualifications (AOR 1.29 [95% CI 0.25-6.56], *p*=0.758)<br>*A-levels (AOR 0.66 [95% CI 0.19-2.26], *p*=0.504)<br>*Degree (AOR 0.73 [95% CI 0.25-2.16], *p*=0.566 *significant at unadjusted level*)<br>- Reserve *(vs Regular)* (AOR 0.61 [95% CI 0.23-1.66], *p*=0.334 *significant at unadjusted level*)<br>- Officer rank *(vs other ranks)* (AOR 0.56 [95% CI 0.17-1.89], *p*=0.350 *significant at unadjusted level*)<br>- Service branch *(vs Army)*<br>*Navy (including Marines) (AOR 0.39 [95% CI 0.13-1.16], *p*=0.091 *significant at unadjusted level*)<br>*RAF (AOR 1.24 [95% CI 0.32-4.88], *p*=0.756)<br>- Length of service (OR 0.93 [95% CI 0.87-0.98] *AORs not presented*)<br>- Left service *(vs serving)* (AOR 2.82 [95% CI 1.08-7.34], *p*=0.034)<br>- Deployed to Iraq *(vs not)* (OR 1.34 [95% CI 0.56-3.22] *AORs not presented*)<br>- Number of childhood adversity factors *(vs 6-16 factors)* | Good |

| | | | |
|---|---|---|---|
| | | *0-1 factors (AOR 0.11 [95% CI 0.04-0.33], *p*<.001)<br>*2-3 factors (AOR 0.19 [95% CI 0.07-0.52], *p*=0.001)<br>*4-5 factors (AOR 0.77 [95% CI 0.24-2.49], *p*=0.664)<br>- Health outcomes *(vs no case)*<br>*PHQ or PTSD diagnosis (AOR 4.65 [95% CI 1.91-11.33], *p*=0.001)<br>*Any PHQ diagnosis (AOR 4.14 [95% CI 1.75-9.81], *p*=0.001)<br>*Any depressive syndrome (AOR 3.08 [95% CI 1.08-8.78], *p*=0.036)<br>*Any anxiety syndrome (AOR 1.69 [95% CI 0.48-5.89], *p*=0.411)<br>*Alcohol abuse (AOR 2.23 [95% CI 0.89-5.60], *p* 0.088)<br>*Somatization disorder (AOR 3.65 [95% CI 1.20-11.03], *p*=0.022)<br>*PTSD (AOR 8.48 [95% CI 2.73-26.33], *p*<.0001) | |
| **Richardson et al (2017)**<br>*Canada* | - Mental health status moderated the relationship between sleep disturbances and suicidal ideation, such that insomnia was associated with suicidal ideation among Canadian Forces personnel without mental health diagnoses, or only one mental health disorder, but not among those with comorbid mental health disorders<br>- Findings highlight the potential benefits of assessing sleep disturbances in Canadian Forces personnel in order to further suicide prevention efforts, and inform preventative interventions targeting sleep in both clinical and non-clinical populations | - Health outcomes *(vs no case)*<br>*PTSD (OR 13.06 [95% CI 9.47-18.00], *p*<.001)<br>*Major depressive disorder (OR 16.44 [95% CI 12.18-22.21], *p*<.001)<br>*Generalised anxiety disorder (OR 11.65 [95% CI 8.35-16.26], *p*<.001)<br>*Panic disorder (OR 8.98 [95% CI 6.21-12.98], *p*<.001)<br>*Alcohol abuse and dependency (OR 4.55 [95% CI 3.04-6.81], *p*<.001)<br>*Insomnia incremental continuous change (OR 2.05 [95% CI 1.85-2.28], *p*<.001)<br>- Past-year mental health status *(vs no mental health disorder)*<br>*1 disorder (OR 12.23 [95% CI 8.33-17.96], *p*<.001)<br>*2+ disorders (OR 33.71 [95% CI 22.82-49.79], *p*<.001)<br>- Female sex *(vs male)* (OR 0.98 [95% CI 0.65-1.48)<br>- Age *(vs 17-29 years)*<br>*30-39 years (OR 1.22 [95% CI 0.86-1.74])<br>*40-49 years (OR 0.87 [95% CI 0.59-1.28])<br>*50-60 years (OR 0.77 [95% CI 0.47-1.25])<br>- Non-white race *(vs white race)* (OR 0.91 [95% CI 0.54-1.53])<br>- Marital status *(vs married)*<br>*Divorced/separated/widowed (OR 1.94 [95% CI 1.31-2.89])<br>*Single, never married (OR 1.34 [95% CI 0.96-1.86])<br>- French language *(vs English)* (OR 0.70 [95% CI 0.48-1.02])<br>- No deployment to Afghanistan *(vs deployment to Afghanistan)* (OR 1.16 [95% CI 0.89-1.51]) | Fair |
| **Richardson et al (2018)**<br>*Canada* | - Sleep disturbances and trauma-related nightmares are associated with suicidal ideation as a function of depressive | **Correlations:**<br>- PTSD symptom severity (correlation coefficient 0.406, *p*<.001) | Fair |

| | | | |
|---|---|---|---|
| | symptoms in treatment-seeking Canadian Armed Forces personnel and veterans<br>- Treating depression in patients who present with sleep difficulties may subsequently help mitigate suicide risk | - Depressive symptom severity (correlation coefficient 0.529, *p*<.001) – only variable emerged as significantly associated with suicidal ideation after accounting for all other health related<br>- Trauma-related nightmares (correlation coefficient 0.172, *p*<.001)<br>- Sleep disturbances (correlation coefficient 0.272, *p*<.001)<br>- Anxiety symptom severity (correlation coefficient 0.382, *p*<.001)<br>- Severity of alcohol use (correlation coefficient 0.073)<br>- Age (correlation coefficient 0.039)<br>- Sex (correlation coefficient 0.052)<br>- Marital status (correlation coefficient (0.025)<br>- Level of education (correlation coefficient (-0.080)<br><br>**Regression models:**<br>- Model 1<br>*Sleep disturbances (β = 0.215, SE 0.035, *p*<.001)<br>*Depressive symptom severity significantly mediated relationship between sleep disturbances and suicidal ideation (bootstrapped unstandardized indirect effect = 0.221, 95% CI 0.175-0.279)<br>- Model 2:<br>*Nightmares (β 0.109, SE 0.031, *p*<.001)<br>*Depressive symptom severity significantly mediated relationship between trauma-related nightmares and suicidal ideation (bootstrapped unstandardized indirect effect = 0.136, 95% CI 0.101-0.177) | |
| **Sareen et al (2007)**<br>*Canada* | - Evidence of an association between combat exposure and witnessing atrocities and negative mental health outcomes<br>- Highlight importance of improving services, education, and outreach to military personnel with emotional problems or perceiving a need for treatment<br>- May be important to move towards providing mental health treatment in primary-care settings to reduce the barriers and stigma related to seeking mental health services | - Deployment experiences<br>*Exposure to peacekeeping operations *(vs no peacekeeping)* (AOR 0.90 [95% CI 0.63-1.27])<br>*Exposure to combat *(vs no combat)* (AOR 1.17 [95% CI 0.80-1.71])<br>*Witnessing atrocities or massacres *(vs not witnessing atrocities)* (AOR 1.98 [95% CI 1.37-2.86])<br>- Perceived need and no help-seeking or DSM diagnosis (OR 3.04 [95% CI 1.83-5.05]) | Fair |
| **Sareen et al (2017)**<br>*Canada* | - Lifetime history of deployment not significantly associated with past-year | **Past-year suicidal ideation:** | Fair |

| | | |
|---|---|---|
| | suicidal ideation, suicide plan or suicide attempts<br>- Exposure to multiple individual deployment related traumatic events (DRTEs) and number of types of DRTEs associated with suicidal ideation, suicide plans and suicide attempts among active military personnel<br>- Relationships between DRTEs and suicidality persisted even after adjustment for child abuse<br>- Most of the relationship between individual DRTEs and suicidal behaviour was accounted for by mental disorders | - Lifetime deployment (AOR1 1.02 [95% CI 0.74-1.40]; AOR2 1.02 [95% CI 0.74-1.41]; AOR3 0.84 [95% CI 0.60-1.16])<br>- Deployment-related traumatic events<br>*Combat exposure (lifetime) (AOR1 1.50 [95% CI 1.15-1.95]; AOR2 1.40 [95% CI 1.07-1.82]; AOR3 1.08 [95% CI 0.82-1.43])<br>*Peacekeeping (lifetime) (AOR1 1.33 [95% CI 1.00-1.76]; AOR2 1.29 [95% CI 0.98-1.71]; AOR3 0.93 [95% CI 0.69-1.25])<br>*Witnessed atrocities (lifetime) (AOR1 1.48 [95% CI 1.08-2.05]; AOR2 1.33 [95% CI 0.95-1.85]; AOR3 1.01 [95% CI 0.72-1.43])<br>*Known someone injured or killed (CF deployment) (AOR1 1.55 [95% CI 1.18-2.03]; AOR2 1.46 [95% CI 1.11-1.92]; AOR3 1.08 [95% CI 0.80-1.44])<br>*Ever been in life-threatening situation and unable to respond because of rules of engagement (CF deployment) (AOR1 2.05 [95% CI 1.53-2.73]; AOR2 1.90 [95% CI 1.42-2.56]; AOR3 1.30 [95% CI 0.95-1.78])<br>*Ever been injured (CF deployment) (AOR1 1.84 [95% CI 1.36-2.49]; AOR2 1.65 [95% CI 1.20-2.25]; AOR3 1.08 [95% CI 0.79-1.49])<br>*Seen ill or injured women or children (CF deployment) (AOR1 1.38 [95% CI 1.07-1.79]; AOR2 1.25 [95% CI 0.96-1.63]; AOR3 0.86 [95% CI 0.65-1.13])<br>*Received incoming artillery, rocket, or mortar fire (CF deployment) (AOR1 1.31 [95% CI 1.00-1.72]; AOR2 1.27 [95% CI 0.96-1.66]; AOR3 0.97 [95% CI 0.73-1.29])<br>*Felt responsible for death of Canadian or ally personnel (CF deployment) (AOR1 2.92 [95% CI 1.99-4.28]; AOR2 2.51 [95% CI 1.68-3.75]; AOR3 1.64 [95% CI 1.08-2.51])<br>*Had a close call but protective gear saved you (CF deployment) (AOR1 1.50 [95% CI 1.09-2.06]; AOR2 1.34 [95% CI 0.97-1.85]; AOR3 0.95 [95% CI 0.68-1.31])<br>*Had difficulty distinguishing combatants from noncombatants (CF deployment) (AOR1 1.51 [95% CI 1.16-1.97]; AOR2 1.42 [95% CI 1.08-1.86]; AOR3 0.98 [95% CI 0.73-1.31])<br>*Military sexual trauma during CF deployment (AOR1 3.01 [95% CI 0.91-9.94]; AOR2 2.57 [95% CI 0.76-8.75]; AOR3 1.96 [95% CI 0.57-6.73])<br>*Total number of deployment-related traumatic events (AOR1 1.10 [95% CI 1.05-1.15]; AOR2 1.08 [95% CI 1.03-1.13]; AOR3 1.01 [95% CI 0.96-1.06])<br><br>**Past-year suicide plans:**<br>- Lifetime deployment (AOR1 1.07 [95% CI 0.68-1.69]; AOR2 1.09 [95% CI 0.69-1.73]; AOR3 0.87 [95% CI 0.54-1.39])<br>- Deployment-related traumatic events |

| | | |
|---|---|---|
| | | *Combat exposure (lifetime) (AOR1 1.50 [95% CI 0.996-2.27]; AOR2 1.36 [95% CI 0.88-2.08]; AOR3 0.98 [95% CI 0.63-1.52])<br>*Peacekeeping (lifetime) (AOR1 2.08 [95% CI 1.34-3.22]; AOR2 1.99 [95% CI 1.28-3.10]; AOR3 1.41 [95% CI 0.89-2.25])<br>*Witnessed atrocities (lifetime) (AOR1 2.04 [95% CI 1.27-3.30]; AOR2 1.74 [95% CI 1.03-2.92]; AOR3 1.39 [95% CI 0.83-2.30])<br>*Known someone injured or killed (CF deployment) (AOR1 1.80 [95% CI 1.20-2.71]; AOR2 1.67 [95% CI 1.10-2.53]; AOR3 1.15 [95% CI 0.74-1.77])<br>*Ever been in life-threatening situation and unable to respond because of rules of engagement (CF deployment) (AOR1 2.49 [95% CI 1.60-3.87]; AOR2 2.21 [95% CI 1.39-3.50]; AOR3 1.42 [95% CI 0.87-2.30])<br>*Ever been injured (CF deployment) (AOR1 1.79 [95% CI 1.07-2.97]; AOR2 1.47 [95% CI 0.85-2.53]; AOR3 0.96 [95% CI 0.56-1.64])<br>*Seen ill or injured women or children (CF deployment) (AOR1 1.76 [95% CI 1.19-2.60]; AOR2 1.54 [95% CI 1.02-2.33]; AOR3 1.01 [95% CI 0.66-1.53])<br>*Received incoming artillery, rocket, or mortar fire (CF deployment) (AOR1 1.78 [95% CI 1.21-2.62]; AOR2 1.71 [95% CI 1.15-2.54]; AOR3 1.26 [95% CI 0.84-1.90])<br>*Felt responsible for death of Canadian or ally personnel (CF deployment) (AOR1 3.59 [95% CI 1.97-6.54]; AOR2 2.81 [95% CI 1.44-5.46]; AOR3 1.78 [95% CI 0.92-3.44])<br>*Had a close call but protective gear saved you (CF deployment) (AOR1 2.17 [95% CI 1.34-3.51]; AOR2 1.83 [95% CI 1.11-3.04]; AOR3 1.33 [95% CI 0.81-2.19])<br>*Had difficulty distinguishing combatants from noncombatants (CF deployment) (AOR1 1.94 [95% CI 1.31-2.89]; AOR2 1.80 [95% CI 1.20-2.71]; AOR3 1.18 [95% CI 0.77-1.80])<br>*Total number of deployment-related traumatic events (AOR1 1.15 [95% CI 1.07-1.23]; AOR2 1.12 [95% CI 1.04-1.20]; AOR3 1.04 [95% CI0.97-1.11])<br><br>**Past-year suicide attempts:**<br>- Lifetime deployment (AOR1 1.09 [95% CI 0.40-2.97]; AOR2 1.10 [95% CI 0.41-3.00]; AOR3 0.89 [95% CI 0.33-2.40])<br>- Deployment-related traumatic events<br>*Combat exposure (lifetime) (AOR1 3.16 [95% CI 1.22-8.12]; AOR2 3.02 [95% CI 1.14-7.96]; AOR3 2.29 [95% CI 0.85-6.18])<br>*Peacekeeping (lifetime) (AOR1 2.92 [95% CI 1.21-7.08]; AOR2 2.81 [95% CI 1.16-6.81]; AOR3 2.03 [95% CI 0.81-5.07]) |

| | | | |
|---|---|---|---|
| | | *Witnessed atrocities (lifetime) (AOR1 3.12 [95% CI 1.12-8.71]; AOR2 2.92 [95% CI 0.99-8.59]; AOR3 2.13 [95% CI 0.76-5.98])<br>*Known someone injured or killed (CF deployment) (AOR1 2.25 [95% CI 0.92-5.52]; AOR2 2.18 [95% CI 0.87-5.47]; AOR3 1.58 [95% CI 0.62-4.02])<br>*Ever been in life-threatening situation and unable to respond because of rules of engagement (CF deployment) (AOR1 5.32 [95% CI 2.13-13.32]; AOR2 5.07 [95% CI 1.96-13.12]; AOR3 3.34 [95% CI 1.27-8.78])<br>*Ever been injured (CF deployment) (AOR1 3.14 [95% CI 1.04-9.46]; AOR2 2.89 [95% CI 0.92-9.02]; AOR3 1.78 [95% CI 0.56-5.64])<br>*Seen ill or injured women or children (CF deployment) (AOR1 2.07 [95% CI 0.78-5.46]; AOR2 1.95 [95% CI 0.71-5.39]; AOR3 1.29 [95% CI 0.47-3.50])<br>*Received incoming artillery, rocket, or mortar fire (CF deployment) (AOR1 1.95 [95% CI 0.84-4.572]; AOR2 1.93 [95% CI 0.82-4.53]; AOR3 1.43 [95% CI 0.62-3.29])<br>*Had a close call but protective gear saved you (CF deployment) (AOR1 4.48 [95% CI 1.58-12.72]; AOR2 4.17 [95% CI 1.40-12.39]; AOR3 2.77 [95% CI 0.95-8.06])<br>*Had difficulty distinguishing combatants from noncombatants (CF deployment) (AOR1 1.92 [95% CI 1.30-2.83]; AOR2 2.15 [95% CI 0.73-6.36]; AOR3 1.41 [95% CI 0.48-4.11])<br>*Total number of deployment-related traumatic events (AOR1 1.25 [95% CI 1.08-1.45]; AOR2 1.23 [95% CI 1.05-1.44]; AOR3 1.14 [95% CI 0.99-1.32]) | |
| **Syed Sheriff et al (2019)**<br>*Australia* | - Lack of association between childhood trauma that was not interpersonal in nature (such as accidents) and adult suicidality<br>- The higher proportion of those who had three or more types of childhood trauma in the military compared with civilians highlights how military service may provide a refuge for a sub-group of recruits who then carry this vulnerability into their service<br>- In the ADF, childhood anxiety was not only independently associated with suicidality, but also and in contrast to civilians, fully mediated the association between childhood trauma and suicidality | - Age *(vs <25 years)*<br>*25-34 years (AOR 1.19 [95% CI 1.09-1.30])<br>*35-44 years (AOR 1.87 [95% CI 1.71-2.05])<br>*45+ years (AOR 1.90 [95% CI 1.73-2.09])<br>- Highest education *(vs Year 10)*<br>*Certificate or diploma (AOR 0.84 [95% CI 0.77-0.91])<br>*Year 11/12 (AOR 0.75 [95% CI 0.69-0.82])<br>*University degree (AOR 0.76 [95% CI 0.70-0.83])<br>- Current significant relationship *(vs no current significant relationship)* (AOR 0.45 [95% CI 0.42-0.47])<br>- Childhood trauma – number of types *(vs none)*<br>*Low (1 or 2) (AOR 1.89 [95% CI 0.97-3.67])<br>*High (3+) (AOR 3.15 [95% CI 1.53-6.49])<br>- Childhood trauma category *(vs none)*<br>*Unclassified (AOR 1.58 [95% CI0.72-3.48])<br>*Non-interpersonal, without interpersonal (AOR 1.53 [95% CI 0.62-3.74]) | Fair |

| | | | |
|---|---|---|---|
| | - Adult-onset depression was also associated with suicidality in the military but not civilian population<br>- Highlight need for whole life approach to understanding suicidality, and importance of categorising the nature of childhood trauma exposure<br>- Potential to inform the development of strategies for suicide prevention | *Interpersonal, without non-interpersonal (AOR 3.42 [95% CI1.65-7.09])<br>*Both interpersonal and non-interpersonal (AOR 3.38 [95% CI 1.45-7.88])<br>- Childhood disorder<br>*Anxiety (AOR 3.72 [95% CI 2.01-6.91])<br>*Depression (AOR 1.90 [95% CI 0.51-7.08])<br>*Alcohol (AOR 1.79 [95% CI 0.76-4.18])<br>- Trauma types first experienced as adult *(vs none)*<br>*Low (1 or 2) (AOR 1.48 [95% CI 0.58-3.75])<br>*High (3+) (AOR 3.54 [95% CI1.43-8.77])<br>- Adult trauma class<br>*Unclassified (AOR 0.98 [95% CI 0.51-1.90])<br>*Non-interpersonal (AOR 1.78 [95% CI 1.01-3.14])<br>*Non-intimate interpersonal (AOR 1.54 [95% CI 0.84-2.81])<br>*Intimate interpersonal (AOR 0.78 [95% CI 0.31-1.95])<br>- Adult onset disorder<br>*Anxiety (AOR 1.19 [95% CI 0.59-2.40])<br>*Depression (AOR 4.16 [95% CI 2.23-7.78])<br>*Alcohol (AOR 1.32 [95% CI 0.77-2.26])<br>- Service branch *(vs Navy)*<br>*Army (AOR 0.94 [95% CI 0.89-1.01])<br>*RAF (AOR 0.84 [95% CI 0.79-0.90])<br>- Rank *(vs officers)*<br>*Non-commissioned officers (AOR 1.17 [95% CI 1.07-1.28])<br>*Other ranks (AOR 1.35 [95% CI 1.22-1.50])<br>- Military experiences<br>*Combat exposure (AOR 1.07 [95% CI 0.61-1.87])<br>*Deployment (AOR 0.97[95% CI 0.92-1.02]) | |
| **Syed Sherriff et al (2020)**<br>*Australia* | - Concerningly high rates of suicidal ideation and attempts in recently transitioned ADF males<br>- Suicidality may be precipitated by the period of relative social instability associated with transitioning<br>- Several factors which are associated with suicidality are common in this population and are potentially identifiable early, such as | - Age *(vs <25 years)*<br>*26-35 years (AOR 1.01 [95% CI 0.65-1.56])<br>*36-45 years (AOR 1.26 [95% CI 0.78-2.02])<br>*46-55 years (AOR 1.20 [95% CI 0.73-1.95])<br>*56+ years (AOR 0.97 [95% CI 0.58-1.62])<br>- Current significant relationship (AOR 0.51 [95% CI 0.40-0.66])<br>- Highest educational attainment *(vs Year 10)*<br>*Year 11/12 (AOR 0.65 [95% CI 0.43-0.98])<br>*Certificate or diploma (AOR 0.78 [95% CI 0.55-1.11]) | Good |

| | | |
|---|---|---|
| | childhood interpersonal trauma and childhood-onset anxiety<br>- Findings have potential to aid early intervention and prevention strategies in identifying those at risk prior to transition<br>- Imply early interventions targeting anxiety and interventions to reduce social instability during transition may be useful to reduce suicidality in the transition period | *University degree (AOR 0.61 [95% CI 0.39-0.96])<br>- Service *(vs Army)*<br>*Navy (AOR 0.96 [95% CI 0.73-1.26])<br>*Air Force (AOR 0.78 [95% CI 0.60-1.01])<br>- Rank *(vs CO)*<br>*NCO (AOR 1.63 [95% CI 1.23-2.15])<br>*Other (AOR 1.82 [95% CI 1.29-2.56])<br>- Previously deployed (AOR 1.42 [95% CI 1.04-1.95])<br>- Childhood-onset trauma *(vs nil)*<br>*Other (AOR 0.79 [95% CI 0.36-1.71])<br>*Non-interpersonal (AOR 1.28 [95% CI 0.65-2.52])<br>*Interpersonal (AOR 2.37 [95% CI 1.29-4.34])<br>*1 or 2 types (AOR 1.40 [95% CI 0.82-2.39])<br>*3+ types (AOR 2.10 [95% CI 0.96-4.60])<br>*Mean number of types (AOR 1.18 [95% CI 0.99-1.41])<br>- Adult-onset trauma<br>*Mean number of types (AOR 1.16 [95% CI 1.05-1.29])<br>*Combat (AOR 1.45 [95% CI 0.83-2.55])<br>- Childhood-onset disorder<br>*Anxiety (AOR 1.93 [95% CI 1.08-3.44])<br>*Depression (AOR 1.82 [95% CI 0.39-8.50])<br>*Alcohol (AOR 1.56 [95% CI 0.46-5.27])<br>*Any disorder (AOR 2.13 [95% CI 1.26-3.59])<br>- Adult-onset disorder<br>*Anxiety (AOR 2.01 [95% CI 1.17-3.46])<br>*Depression (AOR 2.20 [95% CI 1.31-3.67])<br>*Alcohol (AOR 1.65 [95% CI 1.31-3.67])<br>- Adult-onset disorder – number of types *(vs nil)*<br>*Single (AOR 2.77 [95% CI 1.23])<br>*Multiple (AOR 5.46 [95% CI 2.48-12.03])<br><br>**Previously deployed:**<br>- Age *(vs <25 years)*<br>*26-35 years (AOR 0.72 [95% CI 0.41-1.26])<br>*36-45 years (AOR 0.91 [95% CI 0.50-1.65])<br>*46-55 years (AOR 0.86 [95% CI 0.47-1.59]) |

| | | |
|---|---|---|
| | | *56+ years (AOR 0.74 [95% CI 0.40-1.38])<br>- Current significant relationship (AOR 0.49 [95% CI 0.38-0.65])<br>- Highest educational attainment *(vs Year 10)*<br>*Year 11/12 (AOR 0.70 [95% CI 0.45-1.10])<br>*Certificate or diploma (AOR 0.79 [95% CI 0.54-1.16])<br>*University degree (AOR 0.76 [95% CI 0.47-1.23])<br>- Service *(vs Army)*<br>*Navy (AOR 0.92 [95% CI 0.70-1.22])<br>*Air Force (AOR 0.83 [95% CI 0.62-1.12])<br>- Rank *(vs CO)*<br>*NCO (AOR 1.79 [95% CI 1.32-2.43])<br>*Other (AOR 2.05 [95% CI 3.04])<br>- Previously deployed (AOR 1.42 [95% CI 1.04-1.95])<br>- Childhood-onset trauma *(vs nil)*<br>*Other (AOR 0.96 [95% CI 0.42-2.16])<br>*Non-interpersonal (AOR 1.10 [95% CI 0.54-2.23])<br>*Interpersonal (AOR 2.23 [95% CI 1.17-4.25])<br>*1 or 2 types (AOR 1.41 [95% CI 0.80-2.49])<br>*3+ types (AOR 2.10 [95% CI 0.92-4.76])<br>- Childhood-onset disorder<br>*Anxiety (AOR 2.02 [95% CI 1.11-3.68])<br>*Depression (AOR 0.54 [95% CI 0.11-2.53])<br>*Alcohol (AOR 1.16 [95% CI 0.25-5.29])<br>*Any disorder (AOR 2.05 [95% CI 1.18-3.57])<br>- Adult-onset disorder<br>*Anxiety (AOR 1.90 [95% CI 1.08-3.33])<br>*Depression (AOR .61 [95% CI 1.51-4.50])<br>*Alcohol (AOR 1.87 [95% CI 1.06-3.28])<br>- Adult-onset disorder – number of types *(vs nil)*<br>*Single (AOR 3.08 [95% CI 1.31-7.23])<br>*Multiple (AOR 5.34 [95% CI 2.31-12.32])<br>- Combat (AOR 1.29 [95% CI 0.73-2.26])<br>- Mean deployment trauma exposure types (AOR 1.09 [95% CI 1.00-1.18])<br>- Non-deployment adult trauma<br>*Non-interpersonal (AOR 1.53 [95% CI 0.87-2.71])<br>*Interpersonal (AOR 1.19 [95% CI 0.61-2.31]) |

| | | | |
|---|---|---|---|
| | | - Non-deployment adult trauma – number of types *(vs nil)*<br>*Single (AOR 0.86 [95% CI 0.45-1.63])<br>*Multiple (AOR 1.93 [95% CI 1.01-3.70])<br>*Mean (AOR 1.35 [95% CI 1.08-1.68]) | |
| **Taillieu et al (2022)**<br>*Canada* | - Associations between child abuse history and DRTEs on suicide outcomes were largely indirect via their impact on mental disorders<br>- Information regarding the role of specific types of pre-deployment trauma on mental disorders and suicidal behaviour can be used to develop more targeted prevention and intervention strategies aimed at improving the mental health of military personnel | **Suicidal ideation:**<br>*Model 1:*<br>- Any DTRE (AOR 1.26 [95% CI 0.94-1.70])<br>- Any child abuse (AOR 2.15 [95% CI 1.61-2.88], *p*<.001)<br>*Model 2:*<br>- Any DRTE (AOR 0.72 [95% CI 0.50-1.04]<br>- Any child abuse (AOR 1.42 [95% CI 1.02-1.98], *p*<.05)<br>- Major depressive episode (AOR 6.69 [95% CI 4.36-10.26], *p*<.001)<br>- Generalised anxiety disorder (AOR 2.16 [95% CI 1.29-3.63], *p*<.01)<br>- Panic disorder (AOR 0.76 [95% CI 0.41-1.42])<br>- Panic attacks (AOR 2.72 [95% CI 1.83-4.03], *p*<.001)<br>- PTSD (AOR 3.79 [95% CI 2.36-6.08], *p*<.001)<br>- Alcohol abuse or dependence (AOR 1.58 [95% CI 0.92-2.72])<br><br>**Suicide plans:**<br>*Model 1:*<br>- Any DTRE (AOR 1.55 [95% CI 0.1.02-2.37], *p*<.05)<br>- Any child abuse (AOR 2.63 [95% CI 1.65-4.22], *p*<.001)<br>*Model 2:*<br>- Any DRTE (AOR 0.80 [95% CI 0.46-1.38]<br>- Any child abuse (AOR 1.54 [95% CI 0.89-2.66])<br>- Major depressive episode (AOR 15.15 [95% CI 7.49-30.62], *p*<.001)<br>- Generalised anxiety disorder (AOR 1.57 [95% CI 0.77-3.20])<br>- Panic disorder (AOR 1.57 [95% CI 0.77-3.20])<br>- Panic attacks (AOR 1.18 [95% CI 0.60-2.32])<br>- PTSD (AOR 3.80 [95% CI 2.05-7.04], *p*<.001)<br>- Alcohol abuse or dependence (AOR 2.30 [95% CI 1.10-4.81], *p*<.05) | Fair |
| **Thompson et al (2014)**<br>*Canada* | - Mental health had a strong, independent association with suicidal ideation and that physical health also was independently, though less strongly, associated with ideation | - Physical health conditions<br>*Gastrointestinal conditions (OR 3.59 [95% CI2.49-5.18], *p*<.001; AOR 1.66 [95% CI 1.03-2.65], *p*<.05)<br>*Chronic pain or discomfort (OR 2.91 [95% CI 1.76-4.83], *p*<.001; AOR 1.42 [95% CI 0.77-2.61]) | Fair |

| | | |
|---|---|---|
| | - Found no direct association between suicidal ideation and deployment<br>- The current singular focus on psychosocial problems in contemporary veterans should not obscure the potential impact of physical health problems<br>- Highlight importance of considering physical health in population-based suicide prevention efforts and mitigating suicide risk in individual veterans | *Musculoskeletal condition (OR 2.05 [95% CI 1.41-2.97], *p*<.001; AOR 0.80 [95% CI 0.51-1.24])<br>*Diabetes (OR 2.10 [95% CI 1.29-3.43], *p*<.01; AOR 1.57 [95% CI 0.87-2.84])<br>*Respiratory condition (OR 2.10 [95% CI 1.35-3.26], *p*<.001; AOR 1.06 [95% CI 0.65-1.71])<br>*Cardiovascular condition (OR 1.86 [95% CI 1.32-2.64], *p*<.001; AOR 1.46 [95% CI 0.96-2.22])<br>*Obesity (OR 1.73 [95% CI 1.24-2.42], *p*<.001; AOR 1.29 [95% CI 0.89-1.86])<br>*Hearing problem (OR 1.65 [95% CI 1.18-2.30], *p*<.01; AOR 1.03 [95% CI 0.70-1.51])<br>- Mental health condition<br>*Depression or anxiety (OR 11.59 [95% CI 7.79-17.25], *p*<.001; AOR 5.06 2.97-8.62], *p*<.001)<br>*Mood disorders (OR 10.59 [95% CI 6.51-17.22], *p*<.001; AOR 2.91 [95% CI 1.67-5.07, *p*<.001)<br>*Anxiety disorder (OR 8.22 [95% CI 5.79-11.66], *p*<.001; AOR 1.54 [95% CI 0.99-2.41])<br>*PTSD (OR 7.03 [95% CI 5.03-9.81], *p*<.001; AOR 1.31 [95% CI 0.82-2.07])<br>- Number of physical health conditions (OR 1.46 [95% CI 1.31-1.63], *p*<.001; AOR 1.22 [95% CI 1.05-1.42], *p*<.01)<br>- Number of mental health conditions (OR 2.65 [95% CI 2.34-3.01], *p*<.001 AOR [95% CI 1.05-1.42], p<.01)<br>- SF-12 physical health (OR 0.95 [95% CI 0.94-0.97], *p*<.001; AOR 0.98 [95% CI 0.96-0.99], *p*<.05)<br>- SF-12 mental health (OR 0.87 [95% CI 0.86-0.89], *p*<.001; AOR 0.88 [95% CI 0.87-0.89], *p*<.001)<br>- Age *(vs 50-67 years)*<br>*20-34 years (OR 1.03 [95% CI 0.58-3.36]; AOR1 1.39 [95% CI 0.583.36]; AOR2 1.30 [95% CI 0.54-3.13]; AOR3 0.84 [95% CI 0.34-2.08])<br>*35-49 years (OR 1.03 [95% CI 0.58-1.82]; AOR1 1.59 [95% CI 0.97-2.60]; AOR2 1.49 [95% CI 0.94-2.37]; AOR3 0.98 [95% CI 0.61-1.61])<br>- Female sex *(vs male)* (OR 1.69 [95% CI 1.08-2.64], *p*<.05; AOR1 1.14 [95% CI 0.65-2.00]; AOR2 1.35 [95% CI 0.79-2.33]; AOR3 1.01 [95% CI 0.56-1.84])<br>- Marital status *(vs married/common-law)*<br>*Single, never married (OR 1.60 [95% CI 1.00-2.60]; AOR1 1.92 [95% CI 1.05-3.49], *p*<.05; AOR2 1.97 [95% CI 1.10-3.52], *p*<.05; AOR3 2.22 [95% CI 1.14-4.30], *p*<.05) |

| | | | |
|---|---|---|---|
| | | *Separated/divorced/widowed (OR 3.44 [95% CI 2.25-5.26], *p*<.001; AOR1 2.50 [95% CI 1.53-4.07], *p*<.001; AOR2 2.51 [95% CI 1.51-4.17], *p*<.01; AOR3 1.60 [95% CI 0.96-2.68])<br>- Education *(vs university graduate)*<br>*Less than high school graduation (OR 1.34 [95% CI 0.60-3.00]; AOR1 0.59 [95% CI 0.23-1.54]; AOR2 0.59 [95% CI 0.24-1.48]; AOR3 0.50 [95% CI 0.19-1.33]<br>*High school graduation (OR 2.00 [95% CI 1.04-3.72], *p*<.05; AOR1 1.14 [95% CI 0.57-2.30]; AOR2 1.00 [95% CI 0.50-1.99]; AOR3 0.92 [95% CI 0.44-1.93])<br>*Some post-secondary education (OR 2.31 [95% CI 1.21-4.40], *p*<.01; AOR1 1.15 [95% CI 0.57-2.32]; AOR2 1.08 [95% CI 0.54-2.16]; AOR3 1.06 [95% CI 0.50-2.25])<br>- Below low income measure *(vs above)* (OR 2.19 [95% CI 1.27-3.77], *p*<.001; AOR1 1.58 [95% CI 0.84-2.98]; AOR2 1.58 [95% CI 0.84-3.00]; AOR3 1.06 [95% CI 0.56-2.01])<br>- Rank *(vs Officers and cadets)*<br>*Recruits (OR 1.60 [95% CI 0.69-3.71]; AOR1 1.11 [95% CI 0.37-3.35]; AOR2 1.42 [95% CI 0.48-4.27]; AOR3 1.61 [95% CI 0.55-4.75])<br>*Junior NCMs (OR 3.37 [95% CI 1.80-6.29], *p*<.05; AOR1 0.96 [95% CI 0.30-3.13]; AOR2 1.40 [95% CI 0.66-2.99]; AOR3 1.50 [95% CI 0.74-3.04])<br>*Senior NCMs (OR 2.07 [95% CI 1.08-3.96], *p*<.05; AOR1 0.72 [95% CI 0.26-3.03]; AOR2 1.33 [95% CI 0.64-2.78]; AOR3 1.37 [95% CI 0.65-2.91]) | |
| **Thompson et al (2019)**<br>*Canada* | - Weak group identity was associated with difficult adjustment to civilian life and suicidal ideation in CAF Regular Force Veterans within 3.6 years after military release<br>- Highlight importance in suicide prevention of attending to social identity during transition to post-military life | - Perceived adjustment (*vs easy)*<br>*Neither (OR 4.1 [95% CI 1.5-11.4])<br>*Difficult (OR 17.1 [95% CI 8.1-36.3])<br>- Group identity *(vs strong sense of local community belonging, part of a group)*<br>*Strong sense of local community belonging, not part of a group (AOR 6.0 [95% CI 2.0-18.4])<br>*Weak sense of local community belonging, part of a group (AOR 3.8 [95% CI 1.7-8.9])<br>*Weak sense of local community belonging, not part of a group (AOR 9.0 [95% CI 3.7-22.1])<br>- Age *(vs 60+ years)*<br>*<30 years (OR 0.9 [95% CI 0.4-2.6]; AOR 0.7 [95% CI 0.2-3.6])<br>*30-39 years (OR 3.9 [95% CI 1.6-9.4]; AOR 1.3 [95% CI 0.3-5.2])<br>*40-49 years (OR 2.5 [95% CI 1.0-6.2]; AOR 1.2 [95% CI 0.3-4.7])<br>*50-59 years (OR 1.8 [95% CI 0.7-4.4]; AOR 1.2 [95% CI 0.3-3.9])<br>- Female sex *(vs male)* (AOR 1.0 [95% CI 0.5-2.1]) | Fair |

| | | | |
|---|---|---|---|
| | | - Marital status *(vs married/common-law)*<br>*Widowed/separated/divorced (OR 2.1 [95% CI 1.2-3.7]; AOR 1.1 [95% CI 0.6-2.3]<br>*Single/never married (OR 1.4 [95% CI 0.9-2.3]; AOR 1.6 [95% CI 0.8-3.0])<br>- Main employment activity year prior *(vs working or in Reserves)*<br>*Looked for work (OR 3.4 [95% CI 1.4-8.0]; AOR 1.6 [95% CI 0.5-5.6])<br>*School or training (OR 3.5 [95% CI 1.9-6.5]; AOR 1.9 [95% CI 0.9-4.2])<br>*Retired/not looking (OR 1.9 [95% CI 0.9-3.7]; AOR 1.4 [95% CI 0.6-3.2])<br>*Disabled (OR 11.7 [95% CI 6.8-20.1]; AOR 2.0 [95% CI 1.0-4.2])<br>- Household income *(vs Quintile 5 highest)*<br>*Quintile 1 (OR 5.8 [95% CI 2.5-13.4]; AOR 2.1 [95% CI 0.8-5.6])<br>*Quintile 2 (OR 3.7 [95% CI 1.5-8.9]; AOR 1.9 [95% CI 0.7-5.0])<br>*Quintile 3 (OR 3.0 [95% CI 1.2-7.4]; AOR 1.7 [95% CI 0.6-4.8])<br>*Quintile 4 (OR 1.8 [95% CI 0.7-4.9]; AOR 4.6 [95% CI 0.5-4.9])<br>- Rank NCM *(vs commissioned officer)* (OR 2.1 [95% CI 1.3-3.3; AOR 0.7 [95% CI 0.4-1.4])<br>- Mental health problem *(vs no/little)*<br>*Mild/moderate (OR 13.4 [95% CI 4.1-44.2]; AOR 6.6 [95% CI 2.1-21.1])<br>*Severe (OR 79.3 [95% CI 25.9-243.2]; AOR 20.2 [95% CI 6.5-63.1])<br>- Physical conditions *(vs 0 conditions)*<br>*1 conditions (OR 2.3 [95% CI 1.0-5.2]; AOR 1.1 [95% CI 0.4-3.1])<br>*2 conditions (OR 4.7 [95% CI 2.1-10.3]; AOR 1.5 [95% CI 0.5-4.1])<br>*3+ conditions (OR 7.8 [95% CI 3.7-16.7]; AOR 1.7 [95% CI 0.6-4.8]) | |
| **VanTil et al (2021)**<br>*Canada* | - Risk of suicide varies across sub-groups of the veteran population and extends beyond the first few years after release from the CAF<br>- Findings provide evidence to ensure prevention and treatment efforts take into account different risk profiles for male and female veterans | **Male:**<br>- Age at release *(vs ≥45 years)*<br>*<25 years (adjusted HR 2.82 [95% CI 2.17-3.67], *p*<.001)<br>*25-34 years (adjusted HR 2.13 [95% CI 1.61-2.81], *p*<.001)<br>*35-44 years (adjusted HR 1.45 [95% CI 1.12-1.87], *p*=0.005)<br>- Rank at release *(vs junior officer)*<br>*Junior NCM (adjusted HR 1.90 [95% CI 1.56-2.31], *p*<.001)<br>*Senior NCM (adjusted HR 1.32 [95% CI 0.98-1.77], *p*=0.069)<br>*Senior officer (adjusted HR 0.91 [95% CI 0.55-1.51], *p*=0.703<br>- Reserve *(vs Regular)* (adjusted HR 0.95 [95% CI 0.76-1.18], *p*=0.634)<br><br>**Female:**<br>- Age at release <25 years *(vs ≥25 years)* (adjusted HR 1.44 [95% CI 0.82-2.50], *p*=0.201) | Fair |

| | | | |
|---|---|---|---|
| | | - Rank at release NCM *(vs Officer)* (adjusted HR 3.05 [95% CI 1.10-8.46], *p*=0.032)<br>Reserve *(vs Regular)* (adjusted HR 1.01 [95% CI 0.46-2.23], *p*=0.973) | |
| **Varker et al (2022)**<br>*Australia* | - Problem anger made a significant independent contribution to risk of suicidality and violence when controlling for the effects of demographic factors and key mental disorders such as PTSD and depression<br>- Problem anger was also correlated with any suicidality (i.e. ideation, plans or attempts)<br>- When breaking down suicidality into ideation or plans vs attempts, problem anger was significantly correlated with both ideation and plans compared to no suicidality, and a significant correlate of attempts compared to ideation or plans<br>- Path model findings suggested that problem anger has an influence on suicide attempts which occurs primarily through suicidal ideation, and these effects are closely intertwined with other co-occurring mental health problems | - DAR-5 total anger score (OR1 1.26, [95% CI 1.24-1.28], *p*<.001; OR2 1.25 [95% CI 1.23-1.28], *p*<.001; OR3 1.04 [95% CI 1.01-1.07], *p*<.01)<br>- Age (OR2 1.01 [95% CI 1.01-1.02, *p*<.001; OR3 1.01 [95% CI 1.00-1.01])<br>- Male sex *(vs female)* (OR2 0.87 [95% CI 0.69-1.09]; OR3 0.86 [95% CI 0.66-1.13])<br>- Relationship status *(vs in a relationship and living together)*<br>*Not in a relationship (OR2 2.49 [95% CI 2.03-3.05], *p*<.001; OR3 1.93 [95% CI 1.52-2.45], *p*<.001)<br>*In a relationship, not living together (OR2 2.13 [95% CI 1.57-2.88], *p*<.001; OR3 1.94 [95% CI 1.36-2.77], *p*<.001)<br>- Service branch *(vs Army)*<br>*Navy (OR2 1.23 [95% CI 0.99-1.52]; OR3 1.16 [95% CI 0.91-1.49])<br>*Air Force (OR2 1.13 [95% CI 0.91-1.39]; OR3 1.09 [95% CI 0.86-1.40])<br>- Ex-serving status *(vs serving)* (OR2 1.18 [95% CI 0.95-1.46]; OR3 0.95 [95% CI 0.74-1.22])<br>- Medical discharge from service *(vs no)* (OR2 2.36 [95% CI 1.86-2.99], *p*<.001; OR3 1.40 [95% CI 1.05-1.86], *p*<.05)<br>- PCL-5 PTSD total score (OR3 1.03 [95% CI 1.02-1.04], *p*<.001)<br>- PHQ-9 depression total score (OR3 1.16 [95% CI 1.13-1.18], *p*<.001)<br>- AUDIT alcohol use total score (OR3 1.02 [95% CI 1.01-1.04], *p*<.01) | Fair |
| **Williamson et al (2021)**<br>*UK* | - PMIEs were associated with adverse mental health outcomes, including suicidal ideation (compared to those who reported no event exposure)<br>- Likelihood of meeting case criteria for suicidal ideation was greatest in the 'mixed' group compared to those who reported moral injurious or non-morally injurious traumatic events | - Exposure to moral injury (AOR 2.60 [95% CI 1.20-5.66])<br>- Exposure to trauma (AOR 3.33 [95% CI 1.47-7.58])<br>- Exposure to mixed events (AOR 3.89 [95% CI 1.48-10.21]) | Fair |
| **Woodhead et al (2011)**<br>*UK* | - Results do not suggest that the post-national service veteran population in England fares any worse than a comparable population of non-veterans in terms of psychological ill health, marital stability, social disadvantage or treatment-seeking behaviour | **Self-Harm (ever);**<br>- Female veterans *(vs non-veterans)* (OR 1.98 Female veterans *(vs non-veterans)*<br>- 0.54-7.29])<br>- Male veterans *(vs non-veterans)* (OR 0.96 [95% CI 0.43-2.15])<br>- ESLs (AOR 12.36 [95% CI 1.61-94.68]) | Fair |

| | | |
|---|---|---|
| | - Some evidence that early service leavers may experience more mental health problems than those who served for longer | **Suicidal thoughts (ever):**<br>- Female veterans *(vs non-veterans)* (OR 2.59 [95% CI 1.11-6.05])<br>- Male veterans *(vs non-veterans)* (OR 1.17 [95% CI 0.72-1.88])<br>- ESLs (AOR 2.37 [95% CI 1.21-4.66])<br><br>**Suicide attempts (ever):**<br>- Female veterans *(vs non-veterans)* (OR 1.98 [95% CI 0.54-7.29])<br>- Male veterans *(vs non-veterans)* (OR 1.46 [95% CI 0.69-3.09]) |

**Note.** Quality score: 0-5 = Poor; 6-9 = Fair; 10+ = Good
OR: odds ratio; AOR: adjusted odds ratio; HR: hazard ratio
