## Supplementary figures and images for "Risk and Protective Factors for Self-Harm and Suicide Behaviours among Serving and Ex-Serving Personnel of the UK Armed Forces, Canadian Armed Forces, Australian Defence Force and New Zealand Defence Force: A Systematic Review"

### Supplementary 3

**Supplementary 3. PRISMA Flow Diagram**

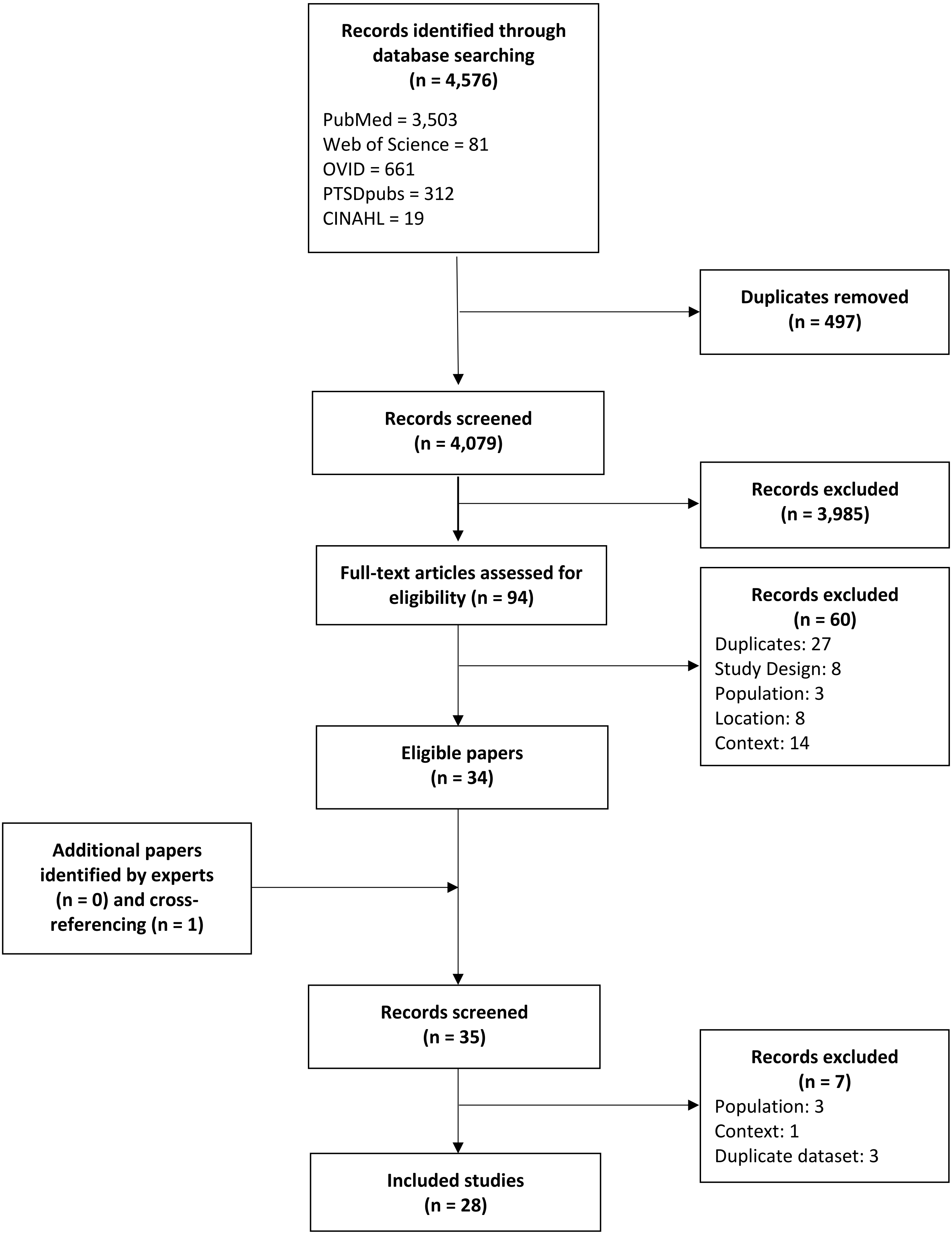
