## Supplementary 4 for "Risk and Protective Factors for Self-Harm and Suicide Behaviours among Serving and Ex-Serving Personnel of the UK Armed Forces, Canadian Armed Forces, Australian Defence Force and New Zealand Defence Force: A Systematic Review"

**Supplementary 4. List of Included Papers**

1. **Afifi et al (2016)** Association of Child Abuse Exposure With Suicidal Ideation, Suicide Plans, and Suicide Attempts in Military Personnel and the General Population in Canada
2. **Belik et al (2009)** Relation between traumatic events and suicide attempts in Canadian military personnel
3. **Bergman et al (2019)** Non-fatal self-harm in Scottish military veterans: a retrospective cohort study of 57,000 veterans and 173,000 matched non-veterans
4. **Bergman et al (2022)** Suicide among Scottish military veterans: follow-up and trends
5. **Boulos and Fikretoglu (2018)** Influence of military component and deployment-related experiences on mental disorders among Canadian military personnel who deployed to Afghanistan: A cross-sectional survey
6. **Harden and Murphy (2018)** Risk factors of suicidal ideation in a population of UK military veterans seeking support for mental health difficulties
7. **Hines et al (2013)** Self-harm in the UK military
8. **Jones et al (2019)** Suicidal ideation, suicidal attempts, and self-harm in the UK Armed Forces
9. **Jones et al (2021)** Do Junior Entrants to the UK Armed Forces have worse outcomes than Standard Entrants?
10. **Kapur et al (2009)** Suicide after leaving the UK armed forces - a cohort study
11. **Kerr et al (2018)** Increased risk of attempted suicide in Australian veterans is associated with total and permanent incapacitation, unemployment and posttraumatic stress disorder severity
12. **Ketcheson et al (2018)** Association between social support and mental health conditions in treatment-seeking Veterans and Canadian Forces personnel
13. **Nelson et al (2011)** Predictors of posttraumatic stress disorder, depression, and suicidal ideation among Canadian Forces personnel in a National Canadian Military Health Survey
14. **O'Toole et al (2015)** Suicidality in Australian Vietnam veterans and their partners
15. **Pinder et al (2012)** Self-harm and attempted suicide among UK armed forces personnel: results of a cross-sectional survey
